# Disentangling osteoarthritis-specific genetic effects from obesity to identify novel therapeutic targets

**DOI:** 10.1101/2025.09.23.25336398

**Authors:** Chen-Yang Su, Masashi Hasebe, Dandan Tan, Bangli Cao, Kevin Liang, Takayoshi Sasako, Vincent Mooser, Wenmin Zhang, Sirui Zhou, Satoshi Yoshiji, Tianyuan Lu, Guillaume Butler-Laporte

## Abstract

Osteoarthritis (OA) significantly impairs mobility and quality of life for hundreds of millions of individuals. Given the limited non-surgical treatment options for OA, genetics may help identify new strategies for treatment. However, many genetic associations with OA arise from its genetic correlation with obesity (measured by body mass index [BMI]), which makes it difficult to find OA-specific genetic associations. This study used a genome-wide association study (GWAS)-by-subtraction approach to separate genetic effects specific to OA from those shared with BMI, using GWAS of 12 OA traits from the Genetics of Osteoarthritis Consortium. Subsequent proteome-wide Mendelian randomization and colocalization analyses across four large proteomics cohorts identified 27 candidate causal proteins influencing OA via pathways not fully mediated by BMI. Among these, extracellular matrix and bone remodeling mediators such as COL6A2, SMAD3, SPP1, and TNFSF11 (RANKL) were highlighted as promising therapeutic targets. Colocalization with expression quantitative trait loci in osteoclasts and other relevant tissues provided additional biological support. Further, actionability assessments identified several proteins already targeted by existing therapies, such as the approved TNFSF11 (RANKL) inhibitor, denosumab, suggesting repurposing opportunities to modulate subchondral bone turnover in OA. This integrative proteogenomic framework clarifies biological mechanisms of OA beyond BMI-related pathways and offers potential targets for intervention.

## Introduction

Osteoarthritis (OA) is the most prevalent form of arthritis, affecting approximately 595 million people worldwide^1^. Progressive loss of articular cartilage, subchondral bone remodeling, synovial inflammation, and osteophyte formation drive chronic pain, disability, and diminished quality of life^2^. As global life expectancy increases, both the prevalence and public health burden of OA are expected to rise. Although any synovial joint can be involved, the disease most often affects weight-bearing joints such as the knees and hips^3^. Current treatment options for OA remain limited, with symptom management primarily relying on pain relief, physical therapy, weight loss, and, in severe cases, joint replacement surgery. Thus, OA is a leading cause of mobility limitations and healthcare utilization in aging populations, highlighting the need for effective prevention and treatment strategies^4^.

The etiology of OA is multifactorial, involving genetic, environmental, and mechanical factors^5^. Among modifiable risk factors, obesity is one of the most significant contributors to OA development and progression^6^. Conventionally measured by body mass index (BMI), obesity exerts both biomechanical and systemic effects on joint tissues. Excess body weight increases mechanical loading on joints, accelerating cartilage wear, while adipose tissue produces pro-inflammatory cytokines that exacerbate OA pathogenesis^7^. The genetic architecture of OA has been extensively explored through genome-wide association studies (GWAS), which have identified numerous loci associated with OA risk^8^. However, many of these loci overlap with BMI-associated variants^9,10^, complicating the identification of OA-specific genetic associations. This overlap raises the possibility that many OA-associated variants act through pathways shared with obesity, limiting their utility as targets for OA-specific interventions.

One way to better disentangle the genetic effects on OA that are not fully mediated by obesity-related pathways is through GWAS-by-subtraction^11^. This framework involves comparing GWAS for OA and obesity-related traits such as BMI to identify genetic signals that are unique to OA after accounting for shared genetic architecture. Combining this framework with Mendelian randomization (MR) can facilitate the identification of novel therapeutic targets specific to OA. MR uses genetic variants to infer causal effects of exposures (e.g., protein levels) on outcomes (e.g., OA), effectively reducing the risk of confounding and reverse causation when its assumptions are met^12,13^. Circulating proteins offer a unique opportunity to advance therapeutic discovery because they are not only frequently implicated in disease pathways^14–18^ but are also druggable as many existing drugs already target extracellular proteins^19^. Recent advancements in large-scale proteomics have identified protein quantitative trait loci (pQTLs)^20–23^, which can be used as genetic instruments in MR to assess how circulating proteins may causally influence diseases^24–29^. Proteogenomics-based MR can therefore prioritize potential therapeutic targets, increasing the likelihood of clinical success compared to conventional drug discovery approaches^30^.

In this study, we applied the GWAS-by-subtraction approach^11^ to disentangle the genetic influences on OA from those potentially mediated through BMI. We assessed 12 OA-related traits curated from the Genetics of Osteoarthritis Consortium^8^. We then conducted proteome-wide MR and colocalization analyses across four large-scale proteomics cohorts to identify proteins implicated in non-BMI-mediated OA traits. Colocalization was further performed with expression quantitative trait loci (eQTLs) data from relevant tissues in the GTEx project^31^ and from osteoclasts^32–34^. Finally, we evaluated the actionability of these candidate proteins to highlight novel therapeutic targets for OA. By focusing on genetic associations of OA not fully mediated by BMI-related pathways, our approach uncovered proteins with potential as targets for interventions in OA, thereby improving our understanding of OA pathogenesis beyond BMI-related mechanisms.

## Results

The study design is presented in **Figure 1**.

**Figure 1.**
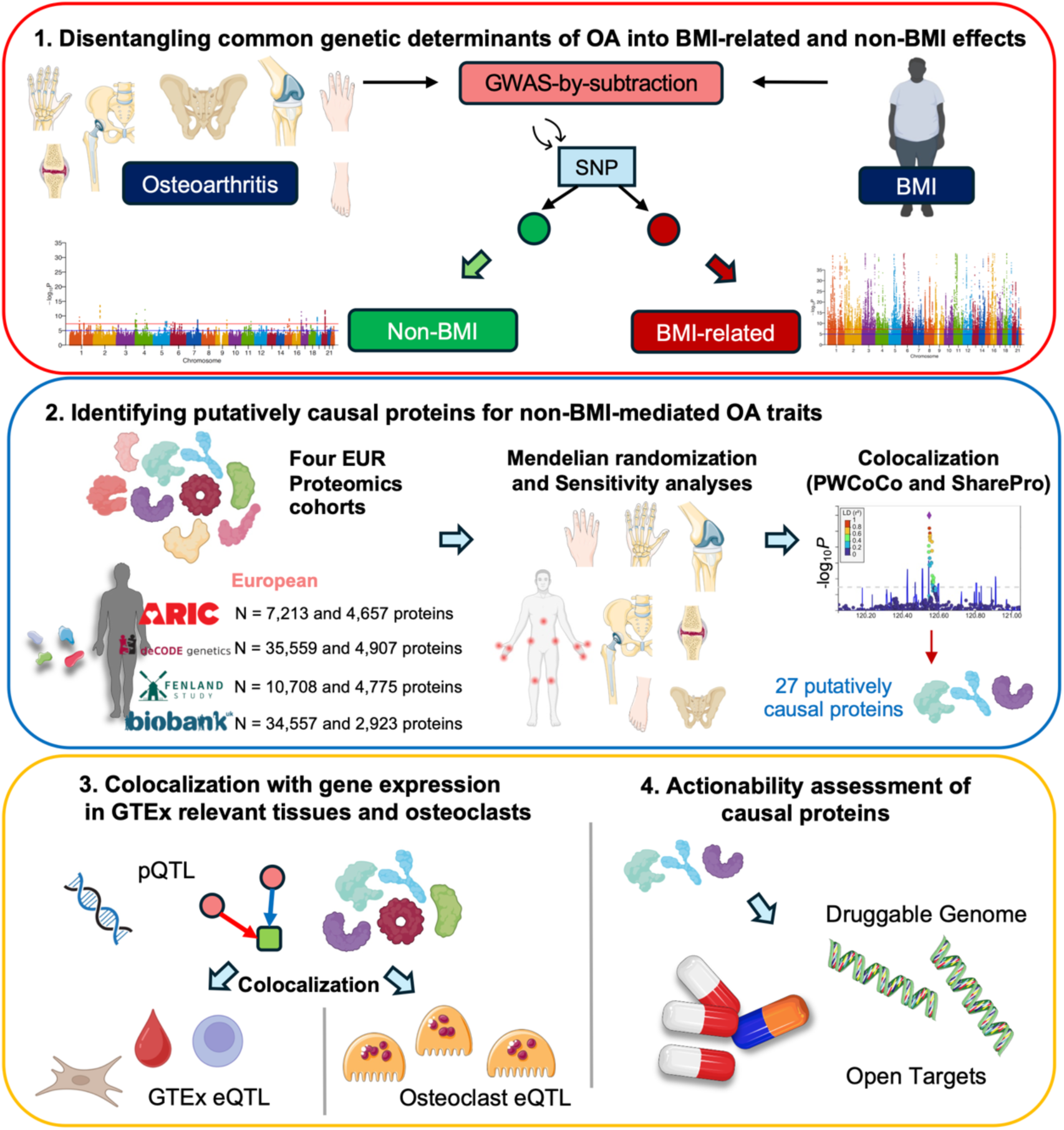
Overview of study design. GWAS-by-subtraction model was applied to partition the genetic effects on osteoarthritis traits onto BMI-related effects potentially associated with OA (BMI-related) and effects not fully mediated by BMI (non-BMI effects). Genetic correlation analysis was performed to identify OA phenotypes that may have a shared genetic basis with BMI prior to and upon subtraction of BMI. Proteome-wide Mendelian randomization and colocalization analyses were conducted using four large-scale European ancestry cohorts to assess putatively causal proteins influencing non-BMI-mediated OA traits. Gene expression colocalization analyses with relevant tissues in GTEx and osteoclasts were performed to assess whether genetic variants associated with the trait of interest influence gene regulation in biologically relevant cell types. Finally, we assessed the actionability of the identified target proteins using the druggable genome and Open Targets Platform to evaluate their potential as therapeutic targets and to identify any existing drugs or compounds that may modulate their activity.

### GWAS-by-subtraction identifies genetic signals for OA not mediated through BMI

We used a GWAS-by-subtraction approach to disentangle common genetic variants associated with OA from those shared with BMI (**Supplementary Figure 1**). We considered 12 OA-related traits (**Supplementary Table 1**) and applied this GWAS-by-subtraction model for each OA trait with BMI. For OA at any site (*n* = 826,690; composed of 177,517 cases, 649,173 controls), as expected, we found that genetic effects not mediated through BMI were less polygenic (**Figure 2A**) compared to those associated with BMI (**Figure 2B**). For non-BMI-mediated OA at any site, the linkage disequilibrium score regression (LDSC) intercept was 0.99 (SE = 0.01), suggesting no evidence of confounding. Similarly, for the BMI-related GWAS, the intercept was 1.01 (SE = 0.03), likewise indicating no evidence of confounding. Notably, we found more BMI-related (38,798 genome-wide significant) than non-BMI-mediated (306 genome-wide significant) OA genetic loci.

**Figure 2.**
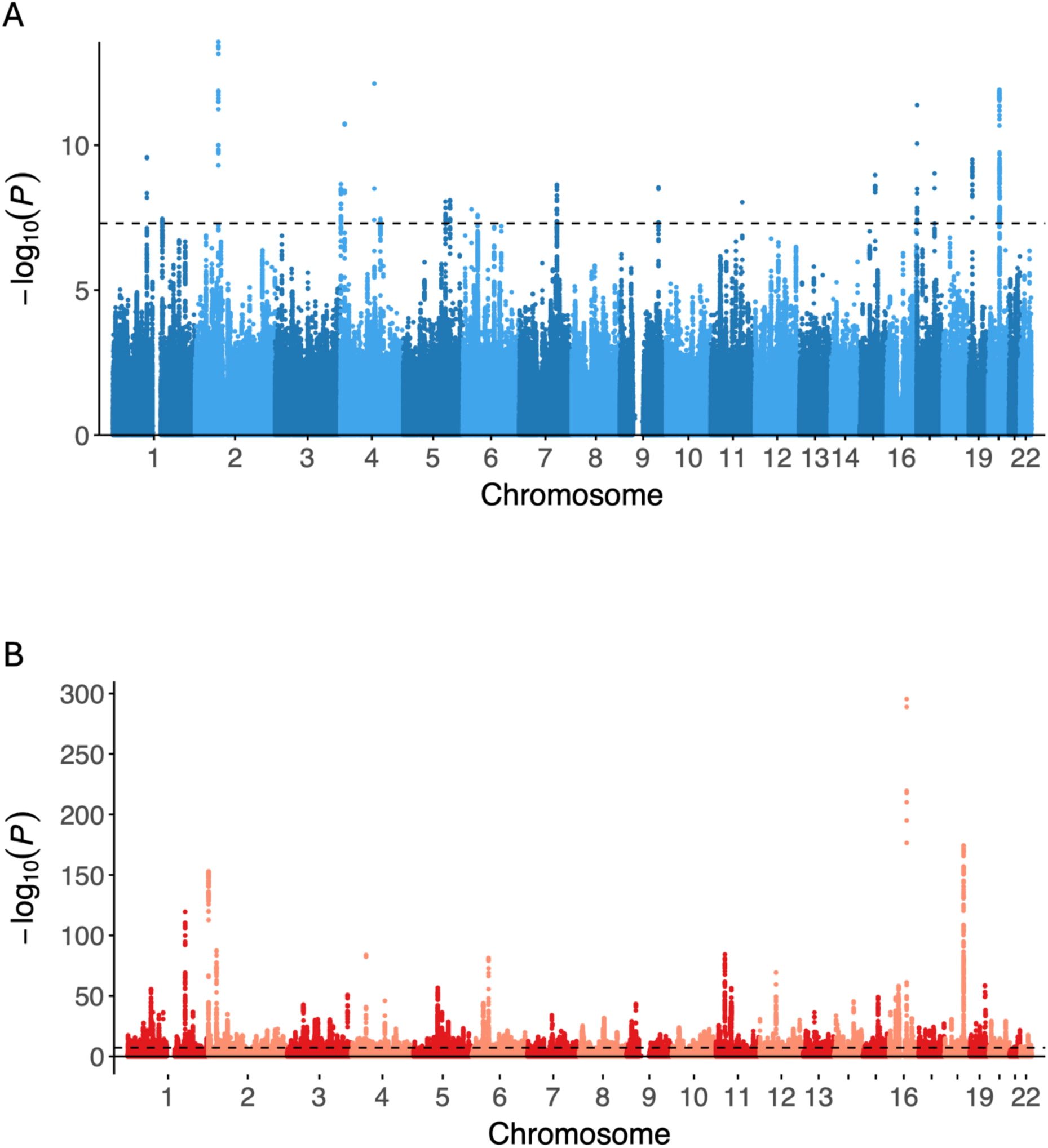
Investigation of the shared genetic architecture between OA at any site and BMI. (A) Non-BMI and (B) BMI-related genetic effects on OA at any site are illustrated. Manhattan plot displays the −log_10_*P* values of association between single nucleotide polymorphisms (SNPs) and the phenotype of interest across the genome. Each dot represents a SNP, plotted according to its chromosomal position along the x-axis and its −log_10_*P* value on the y-axis. Alternating colors are used to distinguish between chromosomes. The black dashed horizontal line indicates the genome-wide significance threshold (*P* = 5 × 10^-8^).

Of non-BMI-mediated SNPs significantly associated with OA at any site, most loci showed attenuated associations after removal of BMI-related genetic effects, while others exhibited strong residual effects, suggesting OA-specific genetic contributions. We observed similar findings for the other 11 OA traits (**Supplementary Figure 2 - 12**). Interestingly, in contrast to OA at any site (*n* = 826,690), upon subtraction of BMI, early-onset OA (*n* = 48,287)—characterized by an onset before the age of 45 years—had no remaining genome-wide significant signals. While these findings could potentially be due to smaller sample size, they may also suggest that its genetic architecture may be more strongly driven by BMI-related pathways or that non-BMI-mediated genetic contributions are weaker in this subgroup.

To validate the GWAS-by-subtraction approach, we estimated the genetic correlation between BMI and each of the 12 OA traits both before and after removing BMI-related effects. As expected, the genetic correlations between BMI and the non-BMI-mediated OA traits were consistently weaker than that observed with the original OA traits across all anatomical sites (**Supplementary Figure 13**), supporting the effectiveness of the GWAS-by-subtraction model in isolating non-BMI-mediated genetic associations. Prior to the removal of BMI-associated genetic associations, we observed moderate to strong positive genetic correlations between BMI and several OA traits. Notably, OA at any site exhibited a correlation of *r_g_* = 0.424 (standard error, SE = 0.023), with even higher estimates observed for early-onset OA (*r_g_* = 0.446, SE = 0.053), knee OA (*r_g_* = 0.469, SE = 0.025), and total knee replacement (*r_g_* = 0.458, SE = 0.026). These findings support a substantial shared genetic basis between BMI and OA, particularly in weight-bearing joints. In contrast, traits such as finger OA (*r_g_* = 0.149, SE = 0.039) and hand OA (*r_g_* = 0.151, SE = 0.027) displayed relatively weaker correlations, consistent with a more limited mechanical contribution of adiposity to disease risk at these sites. Following the removal of BMI-associated genetic associations using GWAS-by-subtraction, the magnitudes of all genetic correlation estimates were noticeably attenuated. For example, the correlation between BMI and non-BMI-mediated OA at any site was 0.174 (SE = 0.021). Similarly, BMI and non-BMI-mediated early-onset OA was 0.195 (SE = 0.056), and the correlation between BMI and knee OA was 0.223 (SE = 0.022). These findings demonstrate the extent to which BMI contributes to the genetic architecture of OA and highlight the utility of GWAS-by-subtraction in isolating non-BMI-mediated genetic associations.

### Proteome-wide Mendelian randomization and colocalization identify putatively causal proteins

Next, to gain insight into the biological mechanisms through which non-BMI-mediated OA occurs, and to pinpoint potential therapeutic targets acting through OA pathways not fully mediated by BMI, which could be intervened on, we performed MR and colocalization analyses. We considered proteins measured in four large-scale proteomics cohorts of European ancestry as exposures. Their circulating levels were quantified using the SomaScan v4 assay (ARIC^23^, deCODE^21^, and Fenland^22^) or Olink Explore 3072 assay (UKB-PPP^20^). These exposures, excluding proteins mapping to the major histocompatibility complex (MHC), were assessed for their causal effects on the non-BMI-mediated OA traits. The minimum *F*-statistics of these genetic instruments were > 30 in all cohorts, demonstrating low risk of weak instrument bias (**Supplementary Table 2**).

After data harmonization (**Supplementary Table 3 - 6)**, we performed a total of 37,327 tests for associations between proteins and the 12 non-BMI-mediated OA traits using MR, including 9,558 associations in ARIC, 9,200 in deCODE, 9,198 in Fenland, and 9,371 in UKB-PPP (**Supplementary Table 7**). Upon applying Benjamini-Hochberg false discovery rate (FDR) correction^35^, sensitivity analyses, and colocalization with PWCoCo and SharePro (see **Methods**), we found 112 robust associations between 27 unique proteins and the 12 non-BMI-mediated OA traits (**Supplementary Figure 14**, **Supplementary Figure 15 - 26**, and **Supplementary Table 8**). Hereafter, we refer to these 27 proteins as the “target” proteins. These targets had strong colocalization evidence (posterior probability that the protein and the OA trait share the same causal variants, PP.H4 ≥ 0.8) provided by at least one colocalization method. Each protein also had consistent direction for all OA-related traits that it was putatively causal for, as expected (**Supplementary Table 9**).

Of these 27 targets, 18 colocalized with PP.H4 ≥ 0.8 using both PWCoCo and SharePro. We further stratified these 18 prioritized targets based on the number of non-BMI-mediated OA traits a protein was implicated in, the number of cohorts in which the target appeared, and whether evidence was present in both SomaScan and Olink platforms (**Supplementary Note 1**). We identified USP8, ITIH1, ITIH3, and TNFSF12 as the most compelling OA protein targets whose effects were not fully mediated by BMI, supported by evidence from at least two cohorts and both proteomic platforms. Additionally, HHIP, COL6A2, NOG, SEMA3G, SMAD3, FKBP5, and ASIP also showed strong evidence, with support from at least one cohort.

Notably, a one standard deviation increase in genetically predicted ubiquitin carboxyl-terminal hydrolase 8 (USP8) levels was significantly associated with increased risk of OA at any site, knee OA, knee and/or hip OA, and total knee replacement outcomes in ARIC, deCODE, and UKB-PPP cohorts (**Figure 3A**), while ITIH1 was linked to decreased risk of knee and/or hip OA, total joint replacement (TJR), hip OA, and total hip replacement (THR) (**Figure 3B**). HHIP was linked to knee and/or hip OA, TJR, hip OA, and THR (**Figure 3C**). Similarly, COL6A2 increased the risk of OA at any site, knee and/or hip OA, and TJR (**Figure 3D**). These findings suggest that increased levels of USP8, HHIP, and COL6A2 may contribute to the development and progression of OA, whereas elevated ITIH1 levels appear to confer a protective effect against joint degeneration and the need for joint replacement surgery.

**Figure 3.**
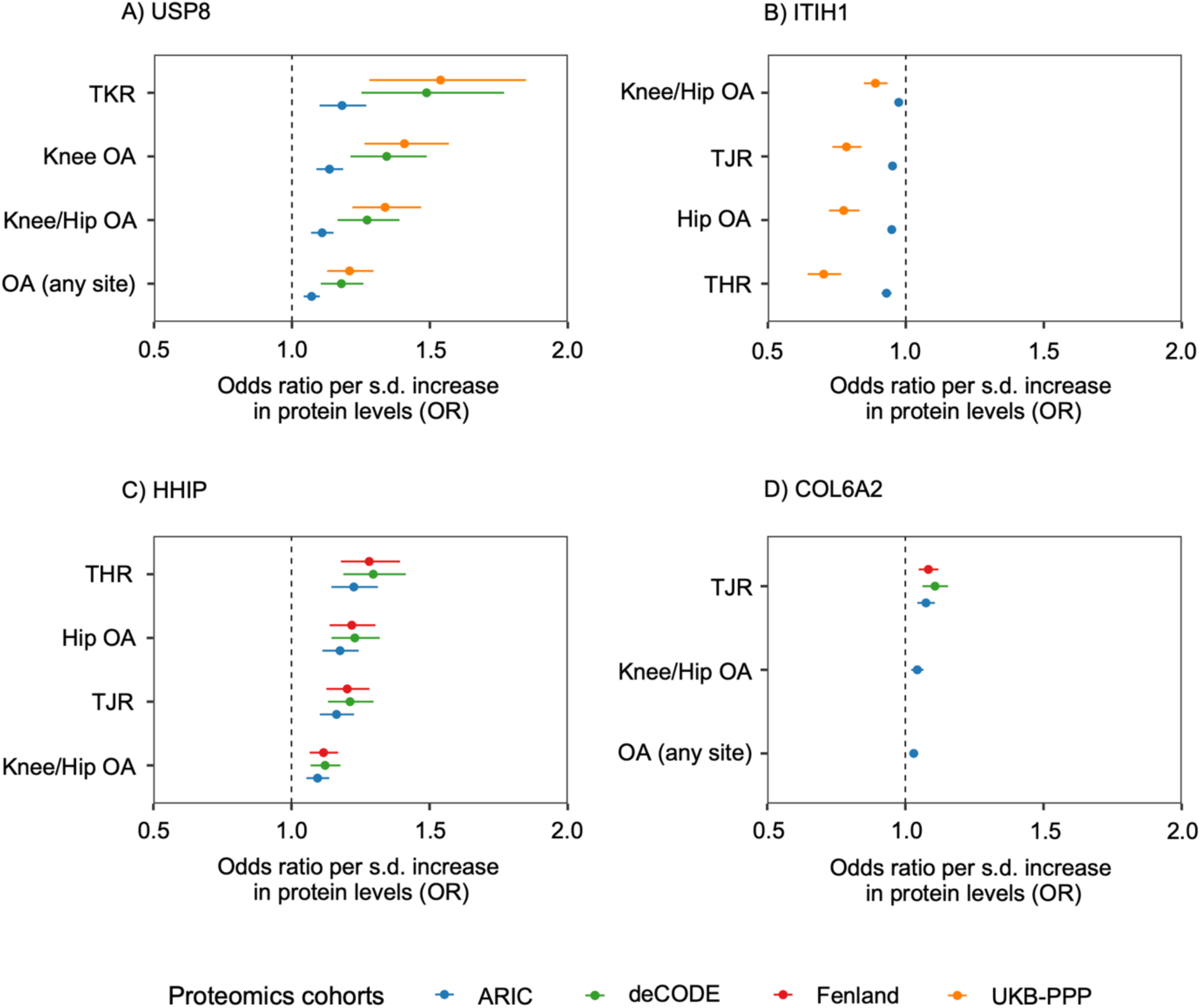
Associations between potential targets and non-BMI-mediated OA traits. Significant MR associations between circulating USP8 (a), ITIH1 (b), HHIP (c), and COL6A2 (d) levels and non-BMI-mediated OA traits. Error bars indicate 95% confidence intervals. Colors indicate the proteomics cohort in which the circulating proteins were measured.

Next, we compared our findings with prior proteome-wide MR studies on OA^36,37^ to assess overlap with previously reported targets and to evaluate whether those associations persisted after subtraction of BMI-related genetic associations. Notably, 12 of the 27 targets (44.4%) were identified by prior studies, but the remaining 15 targets (55.6%) emerged only after removing BMI-related effects, suggesting increased specificity by removing BMI effects (**Supplementary Table 10**).

### Mechanistic classification and protein-protein interaction

By disentangling BMI from the polygenic basis of OA, we uncovered 27 targets with putative causal roles in OA mechanisms acting through non-BMI-related pathways (**Figure 4A** and **Supplementary Note 2**). A wide range of physiological mechanisms have been reported to be active during cartilage degeneration including extracellular matrix organization, skeletal system development, and various signaling pathways^38^. Our findings are consistent with these previously described mechanisms, and the identified targets in our study can be grouped into four broad axes including (1) inflammation and immune modulation; (2) extracellular matrix, cartilage, and bone remodeling; (3) vesicular transport, protein trafficking, and autophagy; and (4) metabolism and cellular stress.

**Figure 4.**
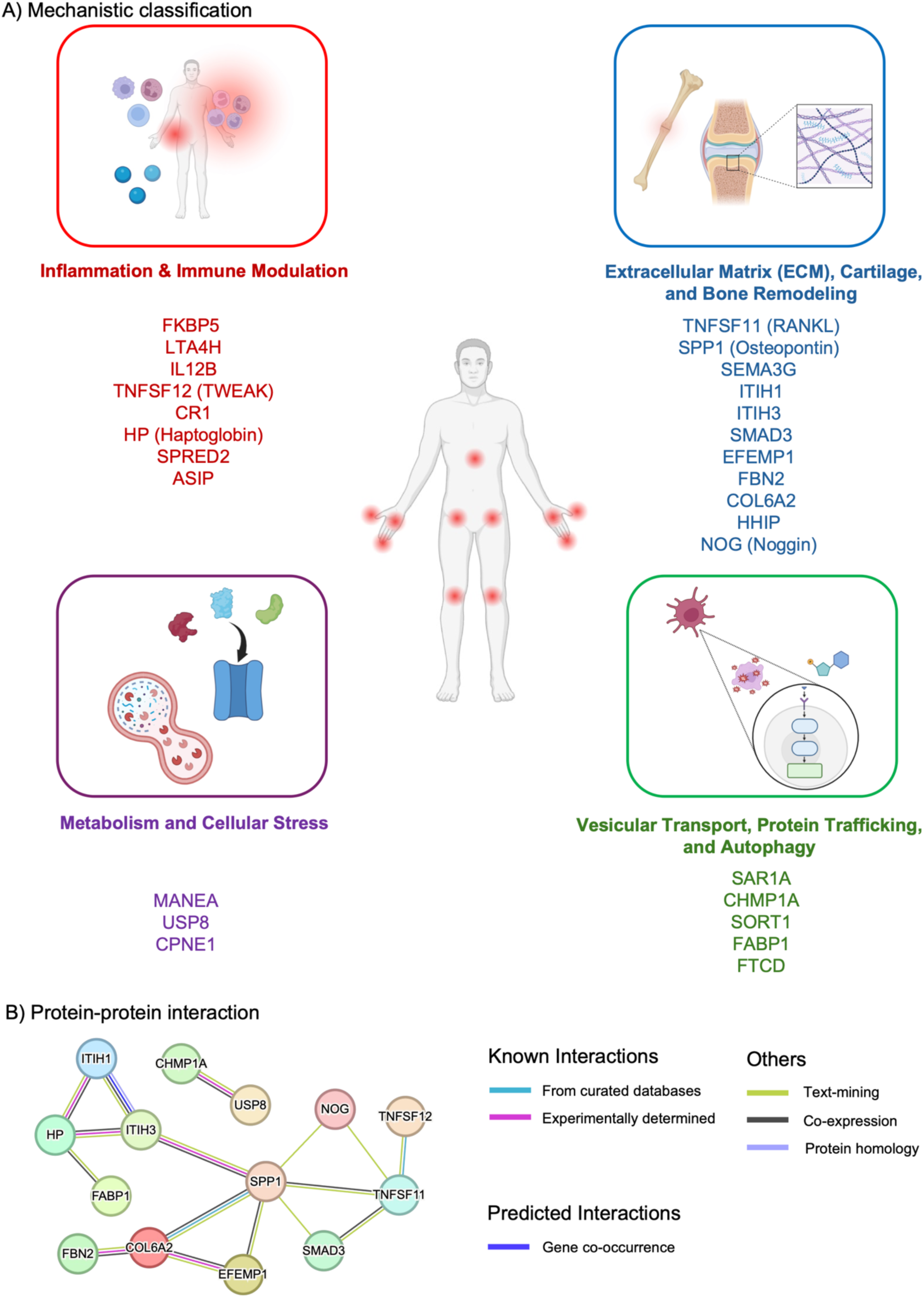
Mechanistic classification of candidate proteins and their interaction network. (A) Mechanistic classification and potential mechanisms through which identified target proteins may act in osteoarthritis. Proteins are grouped into four functional modules illustrated with icons: Inflammation & Immune Modulation (red; FKBP5, LTA4H, IL12B, TNFSF12/TWEAK, CR1, HP, SPRED2, ASIP); Extracellular Matrix (ECM), Cartilage, and Bone Remodeling (blue; TNFSF11 (RANKL), SPP1 (Osteopontin), SEMA3G, ITIH1, ITIH3, SMAD3, EFEMP1, FBN2, COL6A2, HHIP, NOG); Metabolism and Cellular Stress (purple; MANEA, USP8, CPNE1); and Vesicular Transport, Protein Trafficking, and Autophagy (green; SAR1A, CHMP1A, SORT1, FABP1, FTCD). (B) Protein-protein interaction network. Network generated from human proteins shows nodes connected by edges that denote functional associations. Multiple edges between two nodes indicate support from several evidence channels. Edge colors correspond to evidence types as shown at right: Known interactions—curated databases (cyan) and experimentally determined (magenta); Predicted interactions—computational predictions (dark blue); Others—text mining (green), protein homology (grey), and co-expression (black). The line color of edges indicates the type of interaction evidence using the following active interaction sources: text-mining, experiments, databases, co-expression, neighborhood, gene fusion, co-occurrence. We set the minimum required interaction score at medium confidence (0.4; default setting). Disconnected nodes in the network are hidden.

To explore the biological connectivity and functional pathways involving the 27 targets with putative causal roles in OA, we constructed a protein-protein interaction (PPI) network using STRING^39^. Several formed highly interconnected subnetworks, suggesting potential shared biological mechanisms (**Figure 4B**). One prominent network module was centered on SPP1, which exhibited direct interactions with NOG, TNFSF11 (RANKL), SMAD3, EFEMP1, ITIH3, and COL6A2. These proteins collectively map to processes relevant to bone morphogenesis, and extracellular matrix organization^40–46^. Secreted phosphoprotein 1 (SPP1), also known as osteopontin, has active roles in bone remodeling and serves as a positive control, having been previously identified in erosive hand osteoarthritis^47^. Another subnetwork emerged around HP and ITIH3, connected to FABP1 and ITIH1, indicating potential roles in acute phase response and matrix stabilization. Together, these results highlight a coordinated network of pathways contributing to joint degeneration and remodeling in OA.

### Colocalization with relevant tissues in GTEx

To determine the tissue-specific relevance, we performed colocalization analyses with three relevant tissues in GTEx v8^31^ in order to prioritize proteins acting in anatomically and immunologically relevant contexts. We used cultured fibroblasts to explore connective-tissue regulation as well as whole blood and EBV-transformed lymphocytes which could implicate immune cells. In cultured fibroblasts, three unique targets, MANEA, TNFSF12, and USP8, had strong colocalization evidence (PP.H4 ≥ 0.8) (**Supplementary Table 11** and **Figure 5**). In whole blood, three unique targets, SPP1, TNFSF12, and USP8 had strong colocalization evidence (**Supplementary Table 11** and **Figure 5**). None of the target proteins had colocalization evidence with EBV-transformed lymphocytes, likely due to the limited sample size of the eQTL study (**Supplementary Table 11** and **Figure 5**).

**Figure 5.**
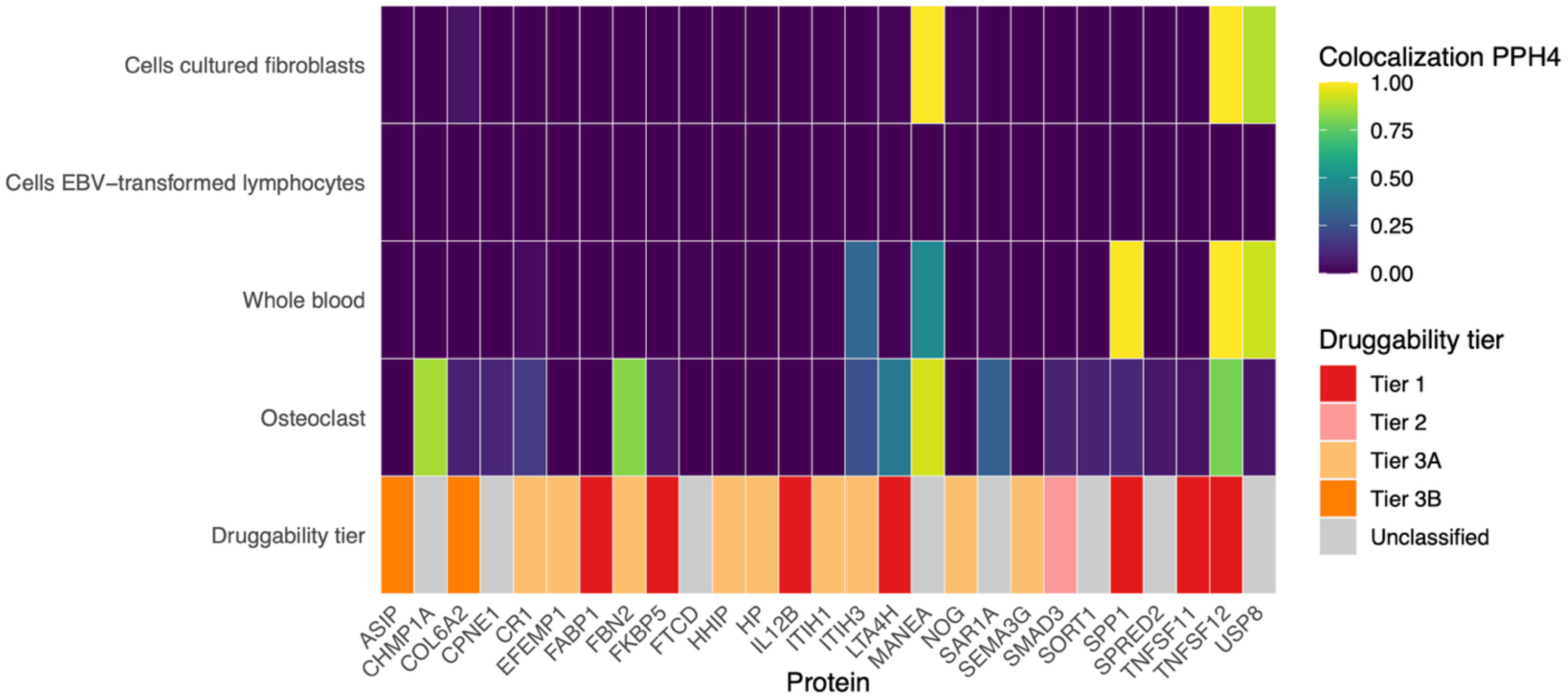
Colocalization across relevant GTEx tissues, osteoclasts, and druggability heatmap or MR-prioritized proteins. Each column is a protein displayed with its protein-coding gene name. Rows display (from top to bottom) PPH4 colocalization in three GTEx tissues: Cells cultured fibroblasts, Cells EBV-transformed lymphocytes, and Whole blood, and PPH4 colocalization in an osteoclast dataset, followed by a categorical Druggability tier row. Tile color encodes PPH4 on a 0-1 scale. The druggability row uses fixed colors to display the druggability tier: Tier 1 = red, Tier 2 = pink, Tier 3A = light orange, Tier 3B = orange, and Unclassified = gray.

### Colocalization with human osteoclast gene expression

To further elucidate cell-specific regulatory mechanisms, we performed colocalization analysis between OA-associated pQTLs and eQTLs derived from human osteoclasts (*n* = 158 individuals)^32–34^. Among the 27 targets, 17 were present in the osteoclast data and were able to be assessed; however, 10 targets, including HHIP, were not available (see **Methods**). We identified strong colocalization (PP.H4 ≥ 0.8) for four unique OA-implicated proteins including MANEA, TNFSF12, CHMP1A, and FBN2 (**Supplementary Table 12** and **Figure 5**). These findings suggest that regulatory variants active in osteoclasts may influence both protein abundance and OA susceptibility, highlighting the relevance of bone-specific transcriptional regulation in OA pathophysiology.

### Actionability assessment of target proteins

To assess therapeutic potential and identify potential drug repositioning opportunities for proteins involved in OA, we incorporated databases from the druggable genome^19^ and Open Targets Platform^48^. Of the 27 targets, 19 (70.4%) were present in Tiers 1 to 3 of the druggable genome which stratifies targets across druggability tiers based on their current or potential interaction with drug-like compounds, while 8 protein targets were unclassified (**Supplementary Table 13** and **Figure 5**).

Tier 1 (approved or clinical-stage targets) included 7 genes: *FKBP5, LTA4H, IL12B, SPP1, TNFSF11, FABP1,* and *TNFSF12*, indicating strong therapeutic precedence and existing pharmacological engagement. Tier 2 (targets closely related to known drug targets) included a single gene, *SMAD3*, which may represent a viable candidate for drug development based on structural or functional similarity to established targets. Tier 3A (secreted or extracellular proteins with biotherapeutic potential) comprised 9 targets, including *SEMA3G, ITIH1, EFEMP1, FBN2, ITIH3, HHIP, NOG, CR1,* and *HP*. Tier 3B (other extracellular proteins) included *ASIP* and *COL6A2*. The remaining 8 targets (*MANEA, USP8, CPNE1, FTCD, SAR1A, CHMP1A, SORT1,* and *SPRED2*) were currently unclassified, reflecting limited evidence for druggability to date. These findings highlight a subset of OA-relevant proteins with high translational potential, particularly among Tier 1 targets, while also pointing to novel candidates in lower tiers that may merit further exploration as emerging therapeutic opportunities.

To further evaluate the translational potential of the 27 OA-implicated targets, we examined evidence from the Open Targets platform, focusing on protein–drug interactions and clinical trial status. Of the six targets previously categorized as Tier 1 in the druggable genome^19^, five (*COL6A2*, *IL12B*, *LTA4H*, *SPP1*, *TNFSF11*, *TNFSF12*) were supported by at least one compound interaction (**Supplementary Table 14** and **Supplementary Table 15**). Notably, *COL6A2*, *IL12B*, and *TNFSF11* each had compounds that reached Phase 4 clinical trials, with one completed trial per target, further supporting their druggability and clinical maturity. In particular, denosumab, a monoclonal antibody against RANKL widely used in osteoporosis^49^, emerged as a concrete repurposing candidate. Our MR results showed directionally concordant results with higher

TNFSF11 (RANKL) associated with greater OA risk, implying that pharmacologic inhibition with denosumab could act in a protective manner. That said, for COL6A2 the most proximate drugs are collagen-degrading enzymes (e.g., collagenase clostridium histolyticum), which are approved for Dupuytren’s disease^50^ and are also used to induce OA-like pathology in animal models^51^. Such catabolic mechanisms run counter to cartilage preservation, underscoring that some seemingly actionable targets may be directionally opposite to what is desirable for OA disease modification. In contrast, the remaining 21 proteins, including several from lower Finan tiers or unclassified genes, had no current compound associations or trial evidence in Open Targets, suggesting these may represent novel or underexplored therapeutic opportunities.

## Discussion

In this study, we demonstrated the utility of GWAS-by-subtraction for disentangling genetic variants associated with OA from those shared with BMI. By isolating non-BMI-mediated genetic effects, we were able to identify loci associated with OA, which were previously masked in conventional GWAS due to their overlap with obesity-related pathways. We performed proteome-wide MR with four large proteomics cohorts and evaluated the putative causal roles of proteins on these non-BMI-mediated OA traits to identify potentially novel OA risk factors and potential drug targets. Our results highlighted extracellular matrix and bone remodeling proteins such as SPP1, TNFSF11 (RANKL), SMAD3, and COL6A2. Notably, TNFSF11 (RANKL) is a secreted ligand with an approved antagonist, denosumab. Our MR results showed that higher TNFSF11 (RANKL) levels were associated with greater OA risk, suggesting that pharmacologic RANKL inhibition by denosumab may be protective, thus supporting its evaluation as a repurposing candidate for OA. Taken together, these findings provide insights into the underlying biological mechanisms of OA that are not fully mediated by BMI-related pathways.

Prior to GWAS-by-subtraction, BMI showed moderate-to-strong genetic correlations with most OA traits, especially knee OA, hip OA, and early-onset OA, consistent with well-established mechanical and metabolic pathways that link excess adiposity to joint degeneration. Accounting for BMI-related effects on OA not only attenuated these correlations but, in some cases, reversed their direction. This pattern suggests that a sizeable fraction of OA susceptibility is independent of mechanical load, systemic adipokine signaling, or other BMI-related factors. Notably, proteins previously considered linked to obesity (e.g., leptin, adiponectin) were absent, supporting the notion that the residual genetic architecture reflects mechanisms independent of adiposity and strengthening the biological validity of the subtraction-based approach.

Several of the identified targets served as positive controls or already had strong prior evidence. TNFSF11 (RANKL) is a key mediator of bone remodeling^52^ and drives subchondral bone turnover^53^, a defining feature of OA progression^54^. Notably, denosumab, a monoclonal antibody to RANKL, is widely used for osteoporosis with a large, well-characterized safety record. Further, preclinical studies in animal models have supported its potential for OA treatment. Moreover, RANKL inhibition is being explored for other skeletal disorders, indicating potential for repurposing in OA^55,56^. SPP1 links bone resorption and immune activation, suggesting potential involvement in OA development^10^. Most OA-associated genetic loci have been identified for knee and hip, with only about 10 identified for hand OA^57^. Notably, the first genetic locus for erosive hand OA recently implicated *SPP1* as a novel susceptibility gene for OA^47^. Our study further supports its role in finger OA, highlighting its relevance in this distinct anatomical and clinical context. *NOG* has previously been identified as an OA-associated gene^8^, with mutations of varying severity known to cause abnormalities in bone and cartilage development^58^.

One specific aspect of our proteomic MR approach warrants emphasis. We instrumented circulating plasma proteins whose most relevant biology in OA likely occurs within the joint microenvironment such as cartilage, synovium, subchondral bone, so the relevance of systemic protein levels is uncertain^59^. This caveat is particularly important for the TGF-β signaling axis linking SMAD3 and COL6A2: SMAD3 functions as a downstream intracellular effector of TGF-β, while COL6A2 is an extracellular matrix component influenced by this pathway. For both SMAD3 and COL6A2, the MR effect directions were discordant with a priori biological expectations, which may reflect compensatory or stage-specific pathology captured in plasma rather than causal benefit from higher circulating levels^30,60^. These nuances are clinically meaningful, as pharmacologic inhibition of TGF-β/SMAD3 signaling such as with losartan has shown beneficial effects in animal models of OA^61–63^, demonstrating that directions inferred from plasma instruments may not translate straightforwardly to tissue-level mechanisms in articulations.

We found potential targets that may have been identified because of residual disease misclassification or BMI-related confounding rather than genuine OA-specific biology. While our analyses provide compelling evidence for the involvement of TNFSF11 (RANKL) in OA pathophysiology, the signal observed for the related TNF-superfamily ligand TNFSF12 (TWEAK) warrants caution. Elevated TWEAK is a well-established biomarker of rheumatoid arthritis activity and joint damage, raising the possibility that its appearance in our dataset reflects residual misclassification of rheumatoid arthritis cases rather than OA-specific biology^64^. Thus, although intriguing, this finding should be interpreted conservatively. Our study also found associations of ASIP in knee and/or hip OA. ASIP has been implicated in monogenic forms of obesity^65^ and functions through melanocortin receptor signaling pathways. Hence, this suggests that residual genetic influences related to obesity, particularly those affecting BMI-regulated pathways, may persist in joints bearing the greatest mechanical and metabolic load despite subtracting out genetic associations with BMI from OA. However, BMI is an imperfect proxy for obesity, as it fails to capture the full complexity of obesity-related metabolic and inflammatory processes that may differentially impact joint health and OA susceptibility.

Lastly, we wish to emphasize our USP8 findings. USP8 is a deubiquitinating enzyme that regulates receptor trafficking, including stabilization of epidermal growth factor receptor, thereby prolonging catabolic signaling in stressed chondrocytes^66^. It also interacts with RNF41^67^, a protein associated with arthropathies, supports its involvement in OA-related pathways. In our study, USP8 showed strong colocalization in cultured fibroblasts and blood, suggesting roles in connective tissue and immune processes. By contrast, no colocalization was observed in EBV-transformed lymphocytes, despite USP8 being expressed in lymphocytes, which may reflect limitations of this transformed cell model or regulation in other immune subsets. Notably, we also found no colocalization with osteoclasts. This absence could reflect limited statistical power (*n* = 158), osteoclast-independent mechanisms consistent with USP8’s role osteogenesis^68^, or the general limitation that many GWAS loci lack matching eQTLs^69^ and effects are tissue-specific. Although direct links to OA remain sparse, USP8’s involvement in key signaling pathways suggests potential relevance and highlights the need for further functional studies in cartilage biology and therapeutic exploration.

This study has additional limitations. First, the OA traits we focused our analyses on were limited to populations of primarily European ancestry. Further studies in more diverse cohorts are needed to validate and extend our findings in non-European ancestries. Second, although we employed GWAS-by-subtraction to eliminate genetic effects of OA potentially mediated through BMI, residual effects by other adiposity traits (e.g., waist-to-hip ratio) cannot be fully excluded. Nevertheless, our genetic correlation analyses revealed a substantially weaker correlation between BMI-related and non-BMI-mediated OA traits compared to the correlation between BMI and OA prior to GWAS-by-subtraction analyses. This suggests that any remaining BMI-related genetic effects were likely greatly reduced. Third, while MR and colocalization analyses provide strong evidence for causality, experimental validation is required to confirm the functional relevance of the identified proteins. Fourth, our study does not address secondary OA, which arises from joint-specific injuries, trauma, or underlying conditions that damage cartilage. Instead, our analysis primarily reflects genetic effects on primary OA, the more common form that develops gradually due to age-related wear and tear. As a result, the findings may not generalize to cases of secondary OA. Last, our analysis included approximately 5,000 proteins from the SomaScan platform and 2,900 proteins from the Olink platform. While these datasets are substantial, they cover only a fraction of the plasma proteome. As a result, other potentially biologically meaningful associations may have been missed, such as intracellular or low-abundance proteins. This highlights the need for more comprehensive and sensitive proteomic assays to better capture the proteome and reveal additional insights.

In summary, this study provides a novel framework for identifying genetic influences of OA that are not fully mediated by BMI-related pathways. Our integrative proteogenomic approach reveals that a substantial amount of OA risk is governed by intrinsic joint and bone biology rather than by excess weight. These findings provide insights into OA pathophysiology, promising avenues for targeted OA therapies, and represent an important advancement toward addressing the significant clinical burden of OA.

## Methods

### GWAS-by-subtraction

To address the genetic correlation between OA and BMI, we applied GWAS-by-subtraction^11^, a genomic structural equation modeling approach that partitions the genetic effects of OA into BMI-related signals potentially associated with OA (*g_BMI_*, BMI-related component) and those not mediated by BMI (*g_nonBMI_*, non-BMI-mediated components). This method enables the identification of genetic variants that contribute specifically to OA. Thus, an important strength of the GWAS-by-subtraction approach is its requirement only for summary statistics. Details on this method have been described elsewhere^11,70,71^. A visualization of the model can be found in **Supplementary Figure 1**. All analyses were performed using GenomicSEM version 0.0.5 and R version 4.1.2.

### Osteoarthritis and BMI cohorts

For OA, we used publicly available GWAS summary statistics from the Genetics of Osteoarthritis Consortium, a large-scale meta-analysis on OA traits which included up to 826,690 individuals from nine populations^8^ of predominantly European ancestry. We considered 12 OA-related traits pertaining to OA at any site, knee and/or hip OA, knee OA, hip OA, TJR, THR, TKR, spine OA, finger OA, thumb OA, hand OA, and early-onset OA (**Supplementary Table 1**).

For BMI, we used the largest available GWAS from the GIANT consortium and UK Biobank meta-analysis which included 681,275 individuals of European ancestry^9^.

### Genetic correlation analyses

To ensure that non-BMI-mediated genetic effects on OA were not correlated with those mediated through BMI, we performed genetic correlation analyses. We used LDSC^72^ to estimate genetic correlations between BMI and each OA trait, using the 1000 Genomes Project LD reference panel^73^ for European ancestry. Upon GWAS-by-subtraction analysis, we also estimated the genetic correlation between BMI and non-BMI-mediated OA traits to assess whether the genetic correlation attenuated.

In genomic structural equation modeling as implemented in GenomicSEM, the LDSC-estimated heritability for each trait and the genetic covariance between the two traits were used as input parameters (𝑆). Specifically, for two traits (𝑖 and 𝑗),

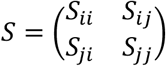

Here, the

- *diagonal* entries are estimated heritability of the two traits (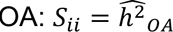 and 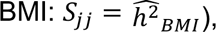),
- the *off-diagonal* entries *(S_ij_* = 𝑆*_ji_*) are estimated genetic covariance.

The proportion of variance in SNP effect sizes shared between OA and BMI was calculated as the squared genetic correlation estimate, with the remaining variance reflecting non-BMI-mediated genetic effects.

### Proteome-wide Mendelian randomization

Following GWAS-by-subtraction, we conducted proteome-wide MR and colocalization analyses to identify putatively causal proteins influencing OA. We leveraged large-scale proteomics datasets from four European ancestry cohorts: ARIC (*n* = 7,213 individuals; 4,657 proteins)^23^, deCODE (*n* = 35,559 individuals; 4,719 proteins)^21^, Fenland (*n* = 10,708 individuals; 4,775 proteins)^22^, and UKB-PPP (*n* = 34,557 individuals; 2,923 proteins)^20^. ARIC, deCODE, and Fenland utilized the SomaScan v4 platform from SomaLogic (https://menu.somalogic.com/) for proteomic profiling, while UKB-PPP employed the Olink Explore 3072 platform. We integrated data from multiple platforms to enhance the robustness and credibility of our findings.

We performed two-sample MR using protein levels as exposures and the 12 non-BMI-mediated OA traits as outcomes. Since ARIC, deCODE, Fenland, and UKB-PPP all had different *cis*-pQTL definitions, we performed LD clumping (*P* value < 5 x 10^-8^; LD *r*^2^ < 0.001) to identify independent pQTLs. Proteins with protein-coding genes in the MHC were excluded owing to its complex LD structure, substantial allelic diversity, and pronounced pleiotropic effects^74^. We denoted pQTLs as *cis* if they were located within 500 kb on either side of the transcription start site of the protein-coding gene, while all other pQTLs were denoted as *trans*. Our analyses only assessed *cis*-pQTLs as genetic instruments for MR since *trans*-pQTLs are more likely to violate the exclusion-restriction assumption due to horizontal pleiotropy. As an extra precautionary measure to mitigate the risk of horizontal pleiotropy, we filtered *cis*-pQTLs by using only those which were associated with a single protein-coding gene and had the highest Open Targets Genetics^75^ V2G score for the corresponding protein-coding gene as we performed previously^24^. Briefly, V2G maps variants to genes by integrating multiple lines of evidence using a machine learning model to provide a quantitative score. When a genetic instrument was not present in the OA outcome, we identified proxies through Plink v.1.9 (parameters: --ld-window = 5000, --ld-window-kb = 5000, --ld-window-r2 = 0.8)^76^. As the LD reference panel, we used 50,000 randomly sampled unrelated European ancestry individuals from the UK Biobank^77^. To avoid allele mismatches, we discarded palindromic proxies with MAF > 0.42.

MR analyses were conducted using either the Wald ratio for proteins with only a single genetic instrument or the inverse-variance weighting method for proteins with two or more independent instruments. *F*-statistics were computed for all genetic instruments to assess risk of weak instrument bias, which can be deemed substantial when *F*-statistics ≤ 10^78^. To account for multiple testing within each cohort, we controlled the false discovery rate at 5% using the Benjamini-Hochberg (BH) procedure^35^, reporting BH-adjusted *P* values, consistent with prior studies^26^.

Sensitivity analyses of MR were conducted to increase the robustness of findings and to account for potential violations of MR assumptions. We performed heterogeneity tests for proteins with two or more genetic instruments. For proteins with three or more genetic instruments, we used alternative MR approaches such as weighted median, weighted mode, and MR-Egger as well as Steiger directionality tests^79^. Consistent effect direction from all MR methods was required and directional pleiotropy was assessed using the MR Egger intercept test, where a nominal *P* value < 0.05 may indicate presence of directional pleiotropy.

We used the R package TwoSampleMR v.0.5.7 to conduct analyses.

### Colocalization analysis

Colocalization analyses were performed using two complementary methods with different model specifications: PWCoCo^25,80^ and SharePro^81^ to determine whether shared genetic variants underlie both protein levels and OA risk and to reduce the risk of confounding due to LD^82^. Briefly, both PWCoCo and SharePro are algorithmic advances of the conventional coloc method, and both were designed to allow for more than one causal variant to exist within a genomic locus. For both methods, we used default parameter settings and performed colocalization for associations which had nominally significant (*P* < 0.05) effect estimates from MR. Colocalization analyses were performed in a 1 Mb region surrounding the lead (lowest *P* value) *cis*-pQTL. We used an LD reference panel composed of 50,000 randomly sampled unrelated European ancestry individuals from the UK Biobank^77^. We considered a colocalization probability (PP.H4) ≥ 0.8 obtained using at least one method as strong evidence of colocalization. Proteins with robust evidence from both MR and colocalization were prioritized for further investigation.

### Comparison with prior MR studies

We compared protein-OA associations identified in this work with two prior studies by Zhang et al.^36^, which identified 26 unique proteins causal for OA, and by Lin et al.^37^, which identified 11 unique causal proteins, respectively.

### Protein-protein interaction

Protein-protein interaction analysis was performed using STRING v12.0 (https://string-db.org/), focusing on the 27 unique proteins identified as putatively causal for OA not fully mediated by BMI-related pathways through proteome-wide MR and colocalization. The analysis included experimentally validated interactions, curated database annotations, and text-mining evidence. Disconnected nodes were hidden for visualization purposes. Network edges reflect known or predicted associations. Functional clusters were visually grouped by interaction strength and pathway proximity.

Protein-protein associations were defined as specific and meaningful interactions in which proteins contribute jointly to a shared biological process or function. Importantly, these associations are not restricted to direct physical binding events; instead, they encompass functional cooperation, such as participation in the same pathway, complex, or regulatory network.

### eQTL colocalization analysis

For colocalization analyses, we used a window size of 1 Mb surrounding the *cis*-pQTL. As the LD reference panel, we used 50,000 randomly sampled unrelated European ancestry individuals from the UK Biobank^77^. For each protein, we reported the highest PP.H4 across all *cis*-pQTL.

#### GTEx

We integrated expression quantitative trait loci (eQTLs) identified in bulk tissues from GTEx v8^31^ to add orthogonal evidence supporting the causal roles of identified proteins by performing colocalization analyses with implicated target proteins. We selected relevant tissues for OA including cultured fibroblasts, whole blood, and EBV-transformed lymphocytes for colocalization analyses. Colocalization was performed using PWCoCo with default parameter settings and strong colocalization evidence was set at PP.H4 ≥ 0.8.

#### Osteoclast

Given that osteoclasts are central mediators of bone resorption and joint remodeling in OA, shared genetic signals between pQTLs and osteoclast gene expression could offer mechanistic support for the bone-active roles of these proteins. We performed colocalization of protein levels with human osteoclast eQTL data^32–34^ (http://www.gefos.org/?q=content/human-osteoclast-eqtl-2018-2020) to add orthogonal evidence for the role of causal proteins in OA-specific pathways. This dataset originates from an eQTL study analyzing gene expression in osteoclast-like cells derived from 158 women of European ancestry, aged 30-70 years, who underwent bone mineral density assessment in Western Australia. Gene expression was quantified using RNA-Seq, while genotyping and imputation yielded data on over 5.3 million variants, with eQTL associations tested using FastQTL within a 2 Mb window of each variant. Of the 27 targets implicated in OA-specific pathways, 10 were not available in the osteoclast eQTL data including EFEMP1, FABP1, HHIP, HP, IL12B, ITIH1, NOG, SEMA3G, ASIP, FTCD. Colocalization was performed using PWCoCo with default parameter settings and strong colocalization evidence was set at PP.H4 ≥ 0.8.

### Actionability assessment

Actionability assessments were conducted using multiple databases, including the Druggable Genome by Finan et al.^19^ and Open Targets platform^48^ (v.24.03) to explore therapeutic potential. In the Druggable Genome, Tier 1 includes genes targeted by approved drugs or those currently in clinical development. Tier 2 comprises genes encoding proteins closely related to known drug targets or with associated drug-like compounds. Tier 3 consists of genes encoding secreted or extracellular proteins that are not currently targeted by drugs but may be tractable based on their characteristics and biological roles. We used the knownDrugsAggregated dataset from Open Targets which provides information on the current clinical trial phase and status of targets for different indications.

## Acknowledgements

C.-Y.S. is supported by a CIHR Canada Graduate Scholarship Doctoral Award (Funding Reference Number: 187673), an FRQS doctoral training scholarship, and a Lady Davis Institute/TD-Bank Scholarship. M.H. is supported by a Graduate Scholarship for Degree-Seeking Study Abroad from the Japan Student Services Organization (JASSO) under the special priority framework for doctoral studies at top-tier global universities in STEM fields, and by the 6th Toshizo Watanabe International Scholarship from The Watanabe Foundation. T.L. has been supported by start-up funding from the Office of the Vice Chancellor for Research and Graduate Education, School of Medicine and Public Health, and Department of Population Health Sciences at the University of Wisconsin-Madison. G.B.L. receives salary support by the FRQS.

The funders had no role in the study design, data collection and analysis, decision to publish, or preparation of the manuscript. We acknowledge Servier Medical Art (https://smart.servier.com/) and BioRender for providing images that were used to create diagrams in this study.

## Author contributions

**Chen-Yang Su:** Data curation; formal analysis; methodology; software; validation; visualization; writing – original draft; writing – review and editing.

**Masashi Hasebe:** Formal analysis; writing – review and editing.

**Tianyuan Lu:** Conceptualization; software; methodology; funding acquisition; supervision; writing – review and editing.

**Guillaume Butler-Laporte**: Conceptualization; methodology; funding acquisition; supervision; writing – review and editing.

All other authors provided critical feedback and reviewed and edited the manuscript.

## Conflict of Interest

The authors have no relevant disclosures.

## Data Availability

The GWAS for BMI-related signals potentially associated with OA and the GWAS for OA genetic signals not fully mediated by BMI will be deposited in the GWAS Catalog upon publication.

All MR analyses adhered to STROBE-MR guidelines (**Supplementary Note 3**).

## Supplementary Figure legends

**Supplementary Figure 1.**
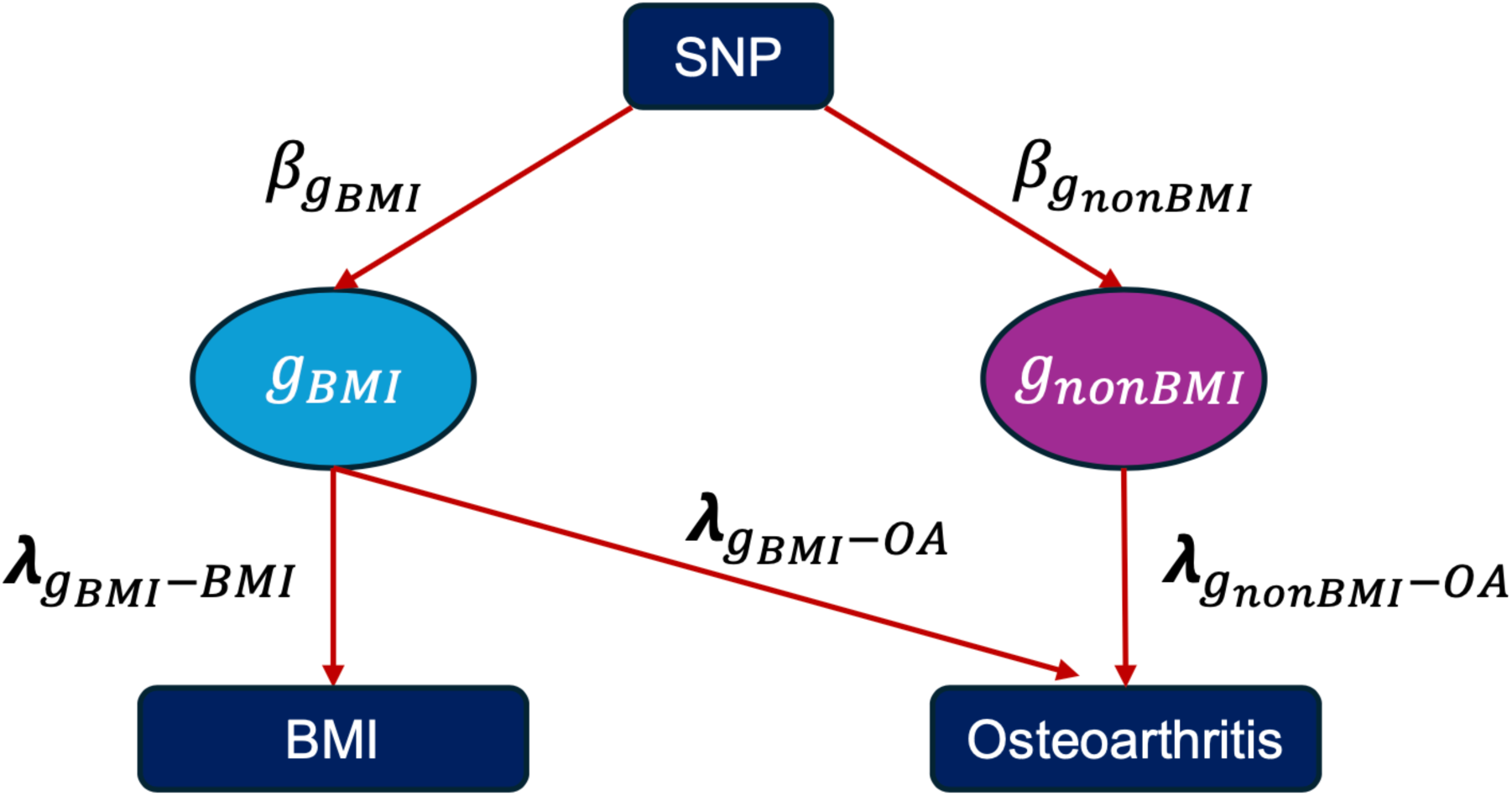
Illustration of GWAS-by-subtraction model for BMI-derived effects on osteoarthritis (OA). *g_BMI_*: BMI genetic latent factors *g_nonBMI_*: non-BMI genetic latent factors 𝛽*_gBMI_*: SNP effect on BMI genetic latent factor 𝛽*_gnonBMI_*: SNP effect on non-BMI genetic latent factor 𝜆*_gBMI-BMI_*_s_: effect of BMI genetic latent factors on genetic components of BMI 𝜆*_gBMI-OA_*: effect of BMI genetic latent factors on genetic components of OA 𝜆*_gnonBMI-OA_*: effect of non-BMI genetic latent factors on genetic components of OA

**Supplementary Figure 2.**
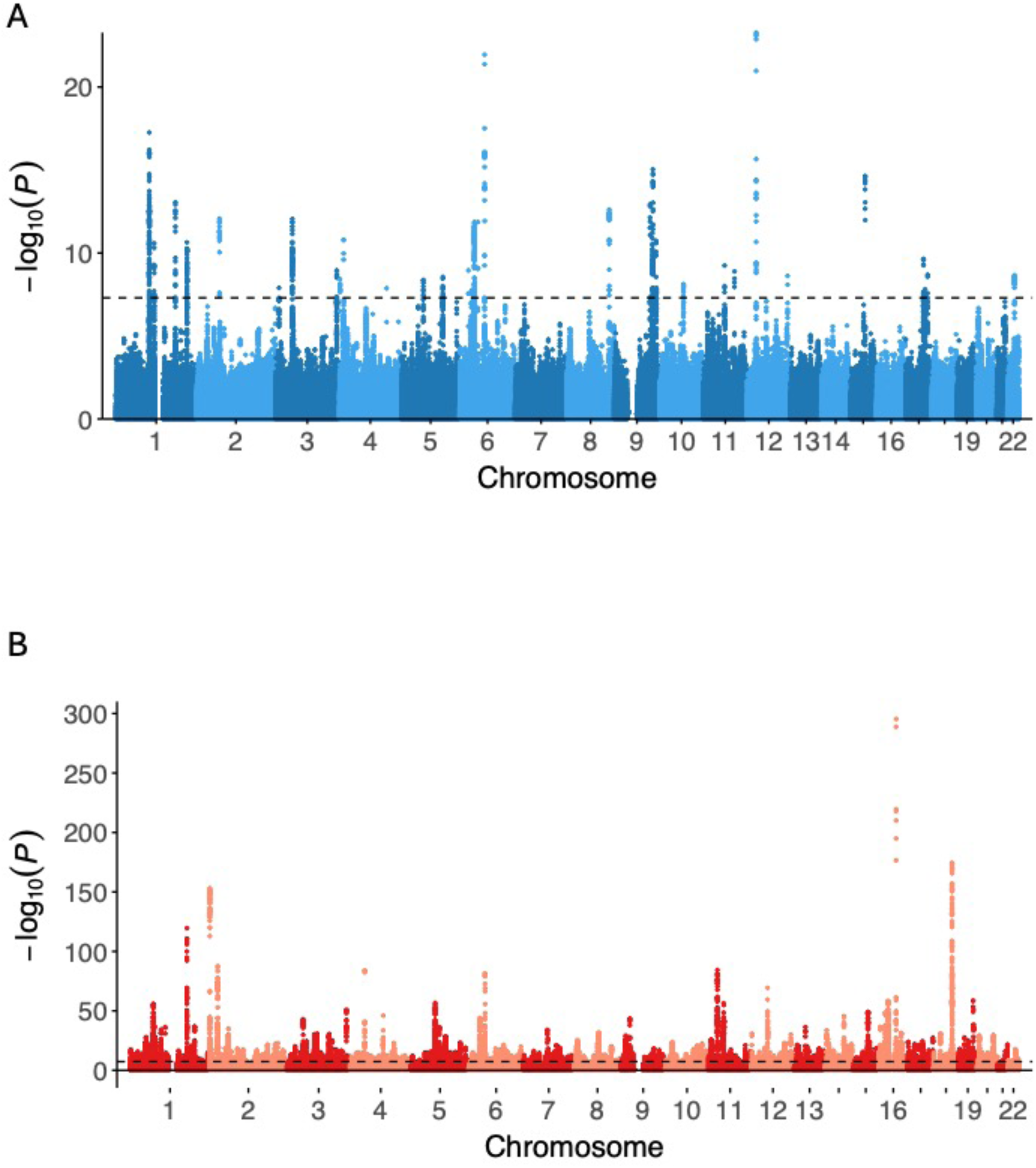
Manhattan plots of non-BMI-mediated and BMI-related GWAS for Hip OA. The non-BMI-mediated panels (A) depict associations not fully mediated by BMI while the BMI-related panels (B) depict associations attributable to BMI-related effects. Each point represents a tested variant. Chromosomes are ordered along the x-axis. Variants are plotted by genomic position (x-axis) and −log_10_*P* (y-axis). Genome-wide significance is indicated by the black horizontal line at *P* = 5 × 10^-8^.

**Supplementary Figure 3.**
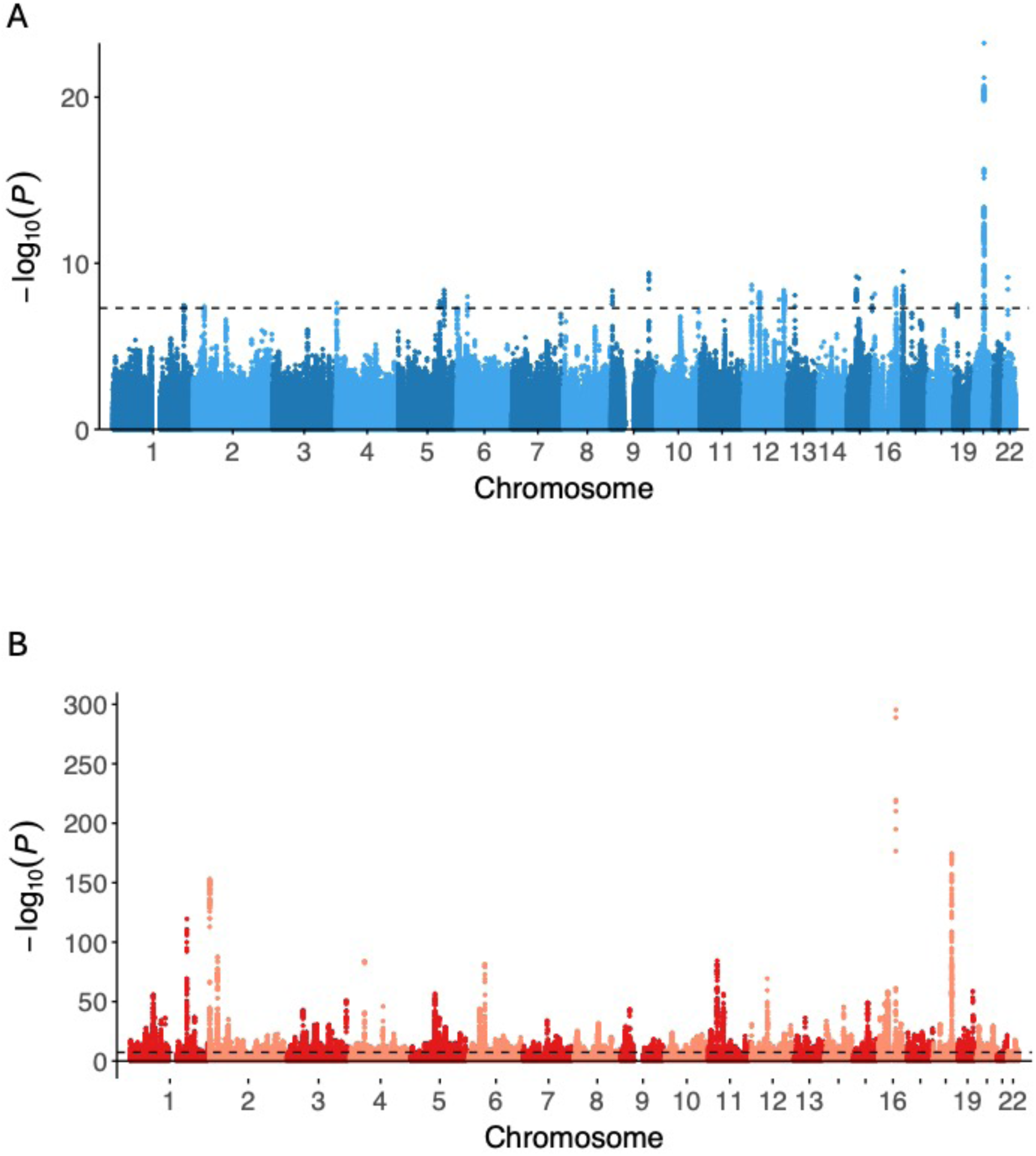
Manhattan plots of non-BMI-mediated and BMI-related GWAS for Knee OA. The non-BMI-mediated panels (A) depict associations not fully mediated by BMI while the BMI-related panels (B) depict associations attributable to BMI-related effects. Each point represents a tested variant. Chromosomes are ordered along the x-axis. Variants are plotted by genomic position (x-axis) and −log_10_*P* (y-axis). Genome-wide significance is indicated by the black horizontal line at *P* = 5 × 10^-8^.

**Supplementary Figure 4.**
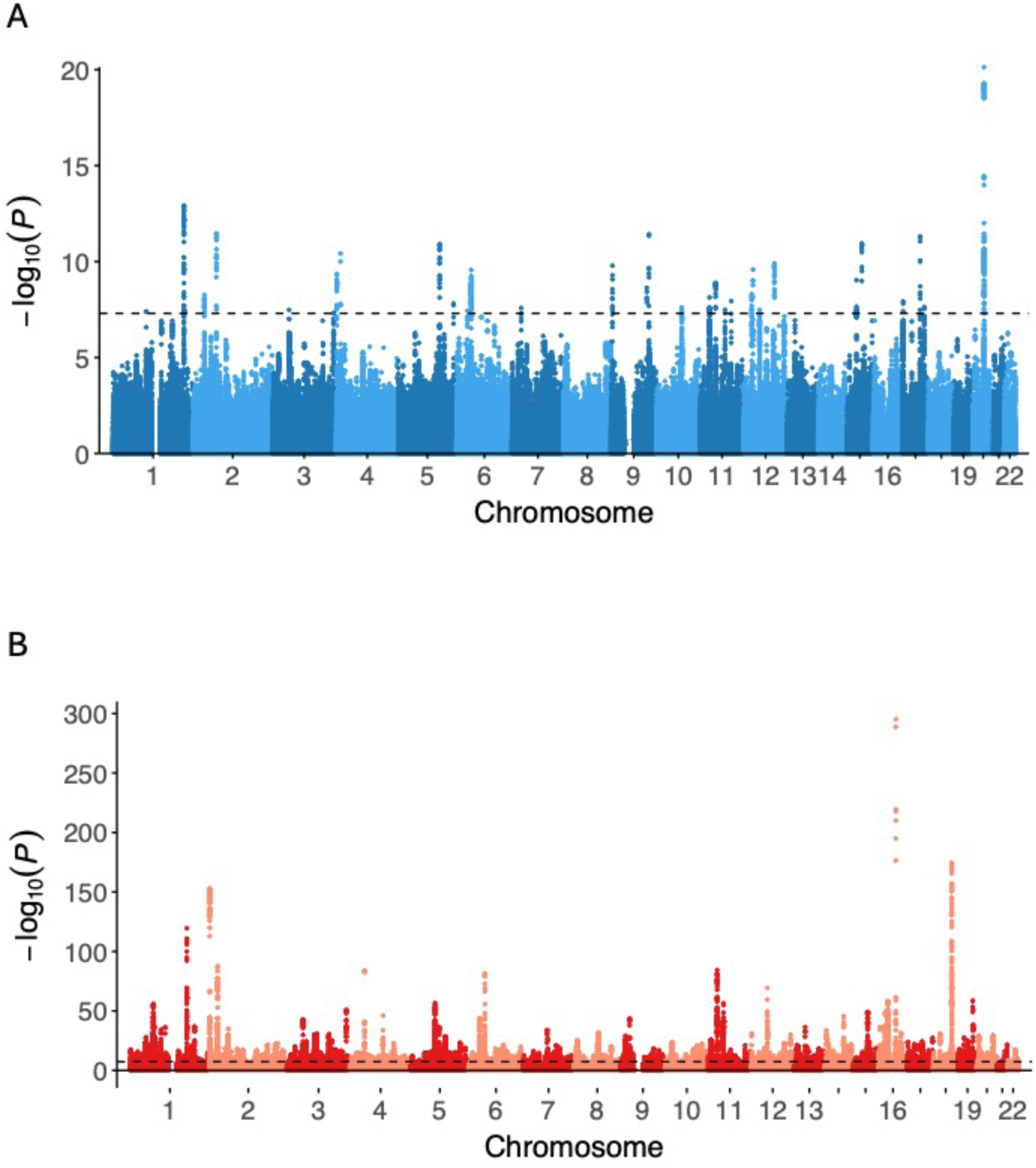
Manhattan plots of non-BMI-mediated and BMI-related GWAS for Knee and/or Hip OA. The non-BMI-mediated panels (A) depict associations not fully mediated by BMI while the BMI-related panels (B) depict associations attributable to BMI-related effects. Each point represents a tested variant. Chromosomes are ordered along the x-axis. Variants are plotted by genomic position (x-axis) and −log_10_*P* (y-axis). Genome-wide significance is indicated by the black horizontal line at *P* = 5 × 10^-8^.

**Supplementary Figure 5.**
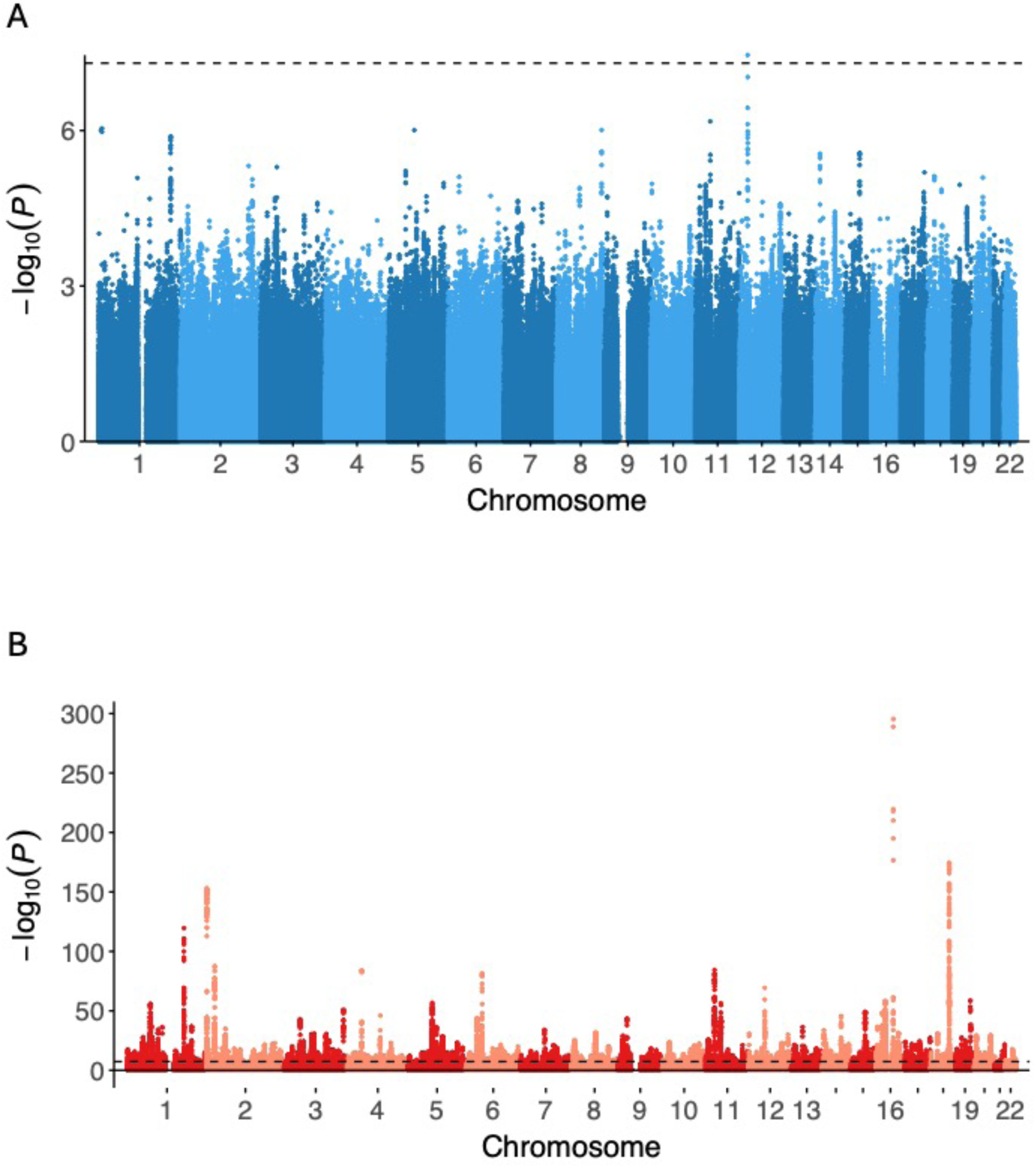
Manhattan plots of non-BMI-mediated and BMI-related GWAS for Spine OA. The non-BMI-mediated panels (A) depict associations not fully mediated by BMI while the BMI-related panels (B) depict associations attributable to BMI-related effects. Each point represents a tested variant. Chromosomes are ordered along the x-axis. Variants are plotted by genomic position (x-axis) and −log_10_*P* (y-axis). Genome-wide significance is indicated by the black horizontal line at *P* = 5 × 10^-8^.

**Supplementary Figure 6.**
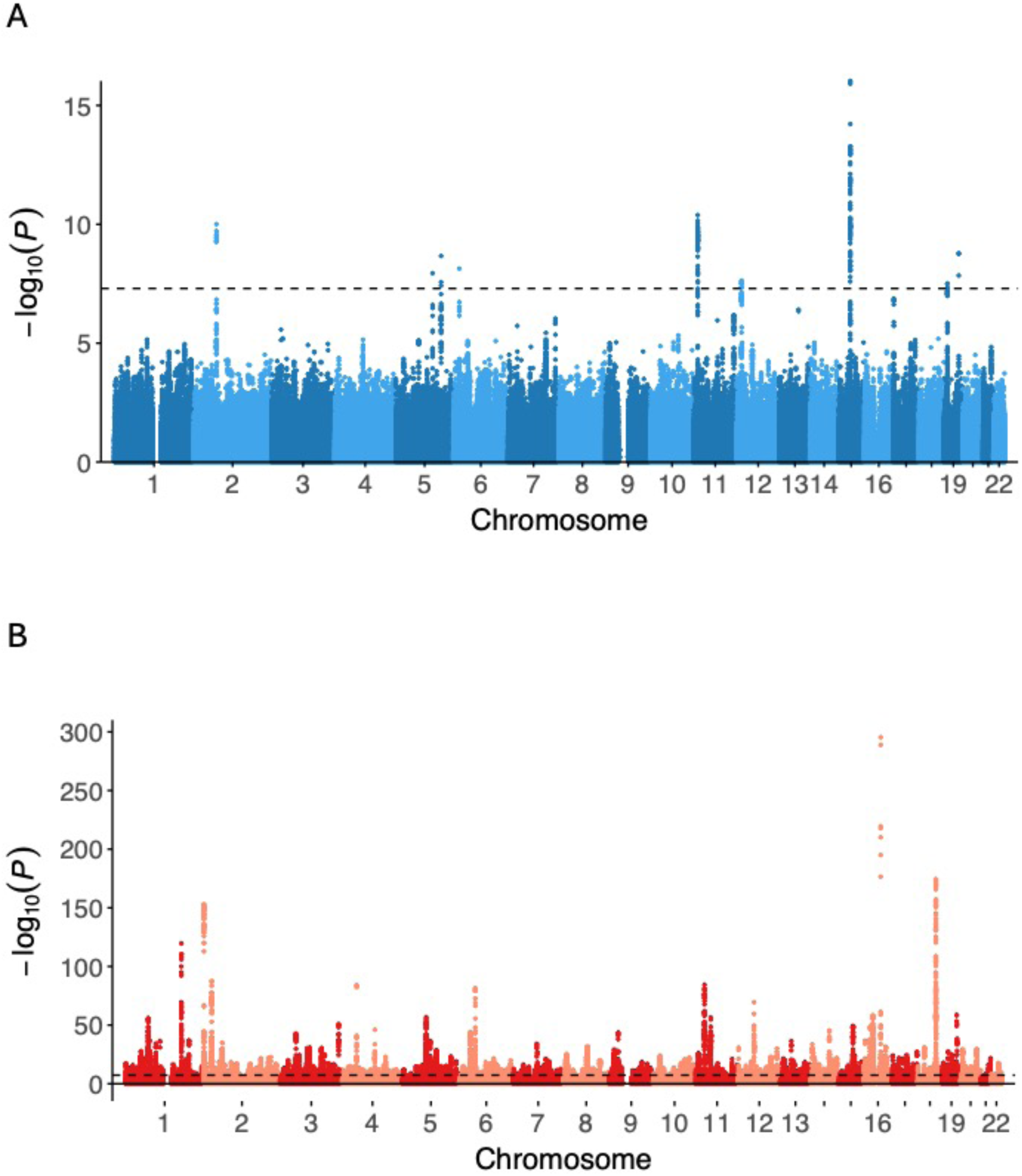
Manhattan plots of non-BMI-mediated and BMI-related GWAS for Hand OA. The non-BMI-mediated panels (A) depict associations not fully mediated by BMI while the BMI-related panels (B) depict associations attributable to BMI-related effects. Each point represents a tested variant. Chromosomes are ordered along the x-axis. Variants are plotted by genomic position (x-axis) and −log_10_*P* (y-axis). Genome-wide significance is indicated by the black horizontal line at *P* = 5 × 10^-8^.

**Supplementary Figure 7.**
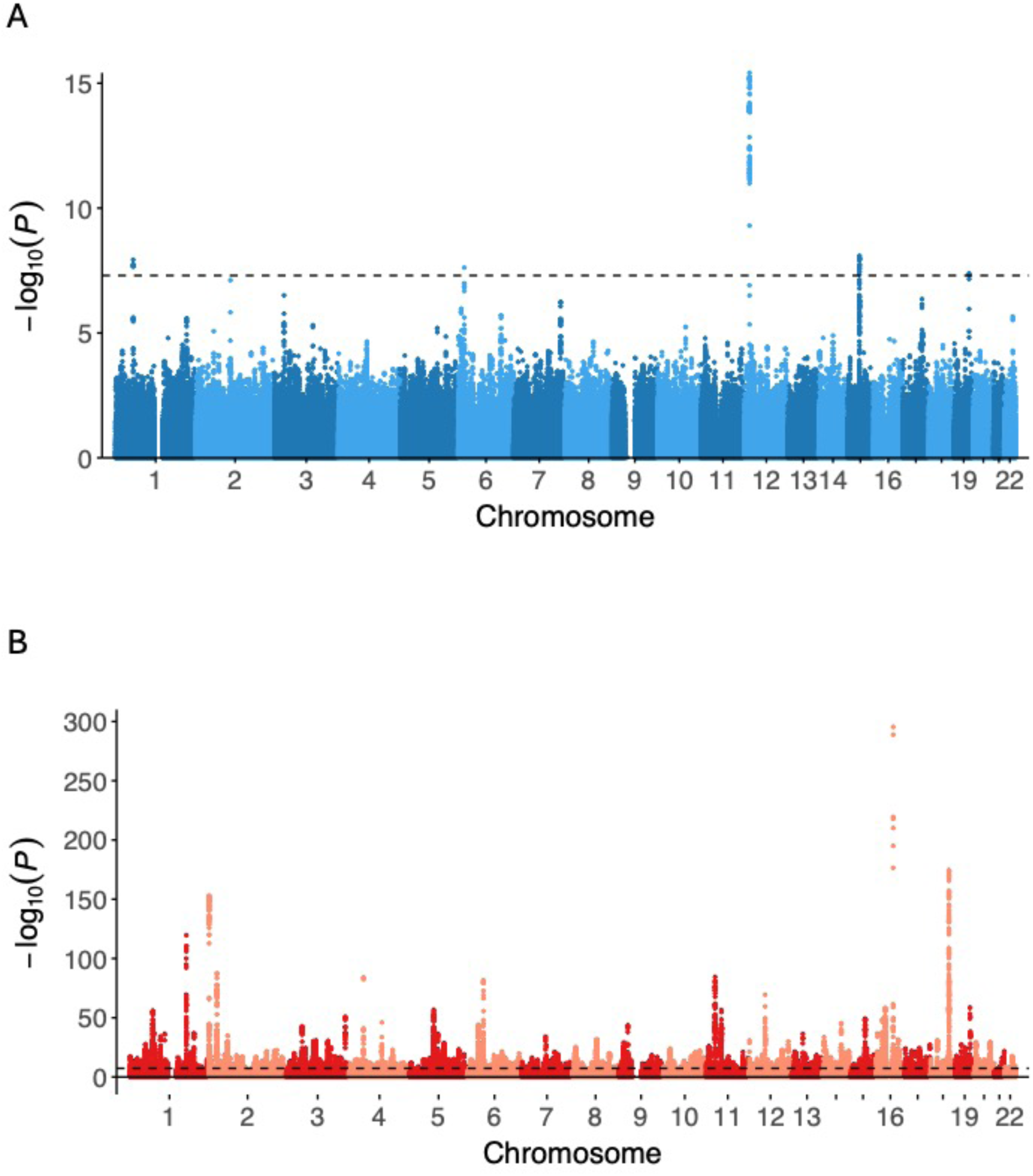
Manhattan plots of non-BMI-mediated and BMI-related GWAS for Finger OA. The non-BMI-mediated panels (A) depict associations not fully mediated by BMI while the BMI-related panels (B) depict associations attributable to BMI-related effects. Each point represents a tested variant. Chromosomes are ordered along the x-axis. Variants are plotted by genomic position (x-axis) and −log_10_*P* (y-axis). Genome-wide significance is indicated by the black horizontal line at *P* = 5 × 10^-8^.

**Supplementary Figure 8.**
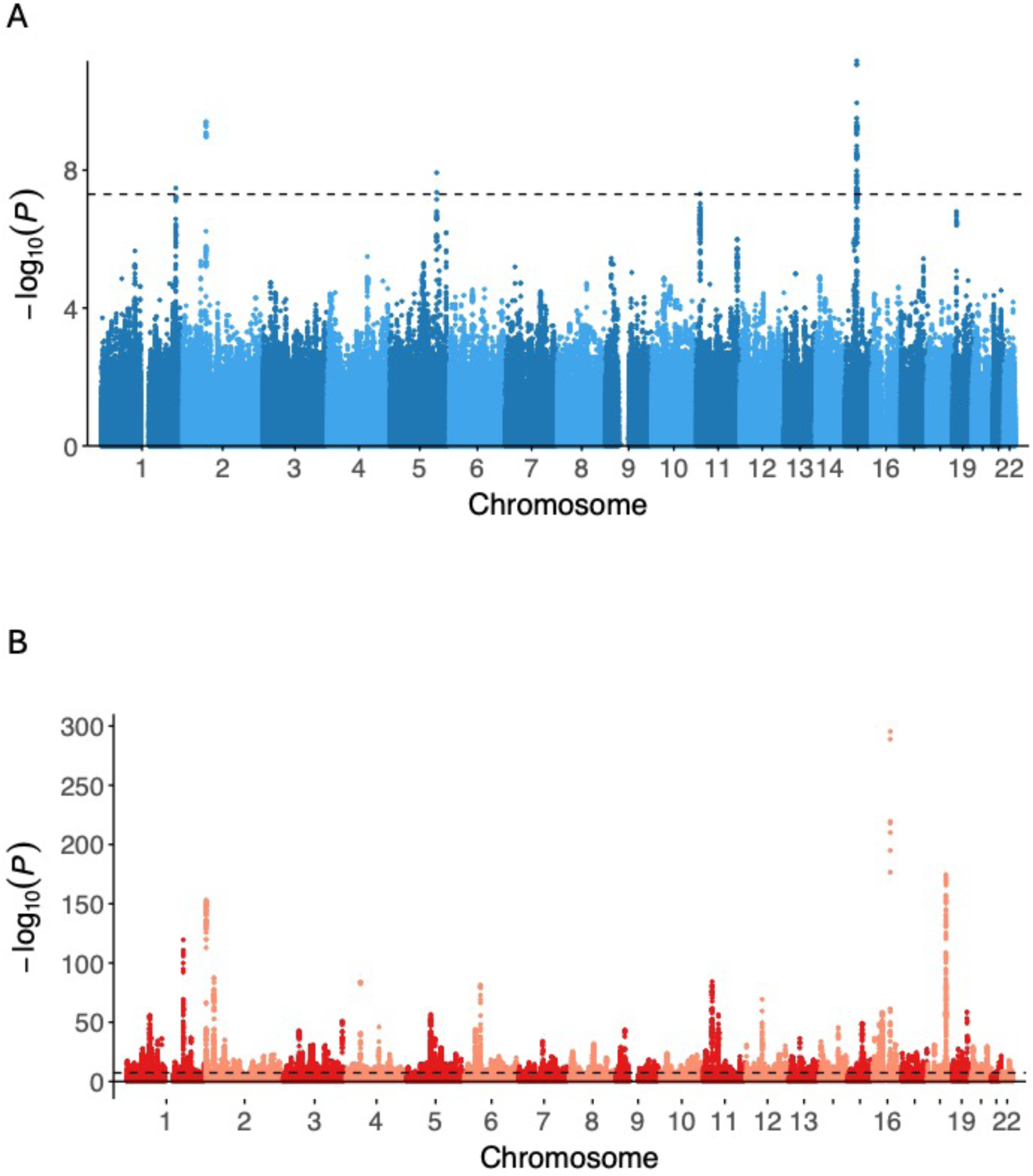
Manhattan plots of non-BMI-mediated and BMI-related GWAS for Thumb OA. The non-BMI-mediated panels (A) depict associations not fully mediated by BMI while the BMI-related panels (B) depict associations attributable to BMI-related effects. Each point represents a tested variant. Chromosomes are ordered along the x-axis. Variants are plotted by genomic position (x-axis) and −log_10_*P* (y-axis). Genome-wide significance is indicated by the black horizontal line at *P* = 5 × 10^-8^.

**Supplementary Figure 9.**
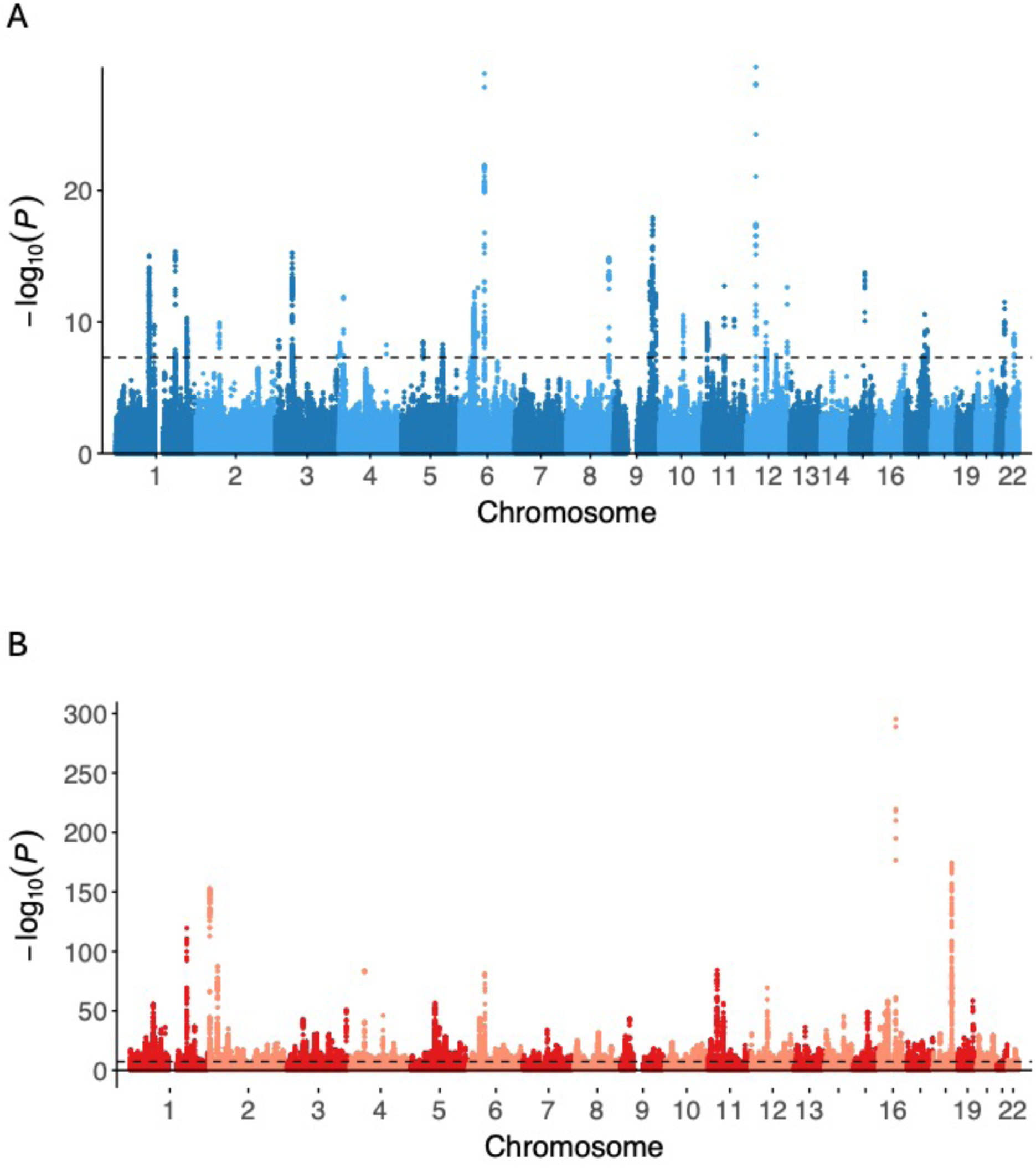
Manhattan plots of non-BMI-mediated and BMI-related GWAS for Total Hip Replacement (TJR). The non-BMI-mediated panels (A) depict associations not fully mediated by BMI while the BMI-related panels (B) depict associations attributable to BMI-related effects. Each point represents a tested variant. Chromosomes are ordered along the x-axis. Variants are plotted by genomic position (x-axis) and −log_10_*P* (y-axis). Genome-wide significance is indicated by the black horizontal line at *P* = 5 × 10^-8^.

**Supplementary Figure 10.**
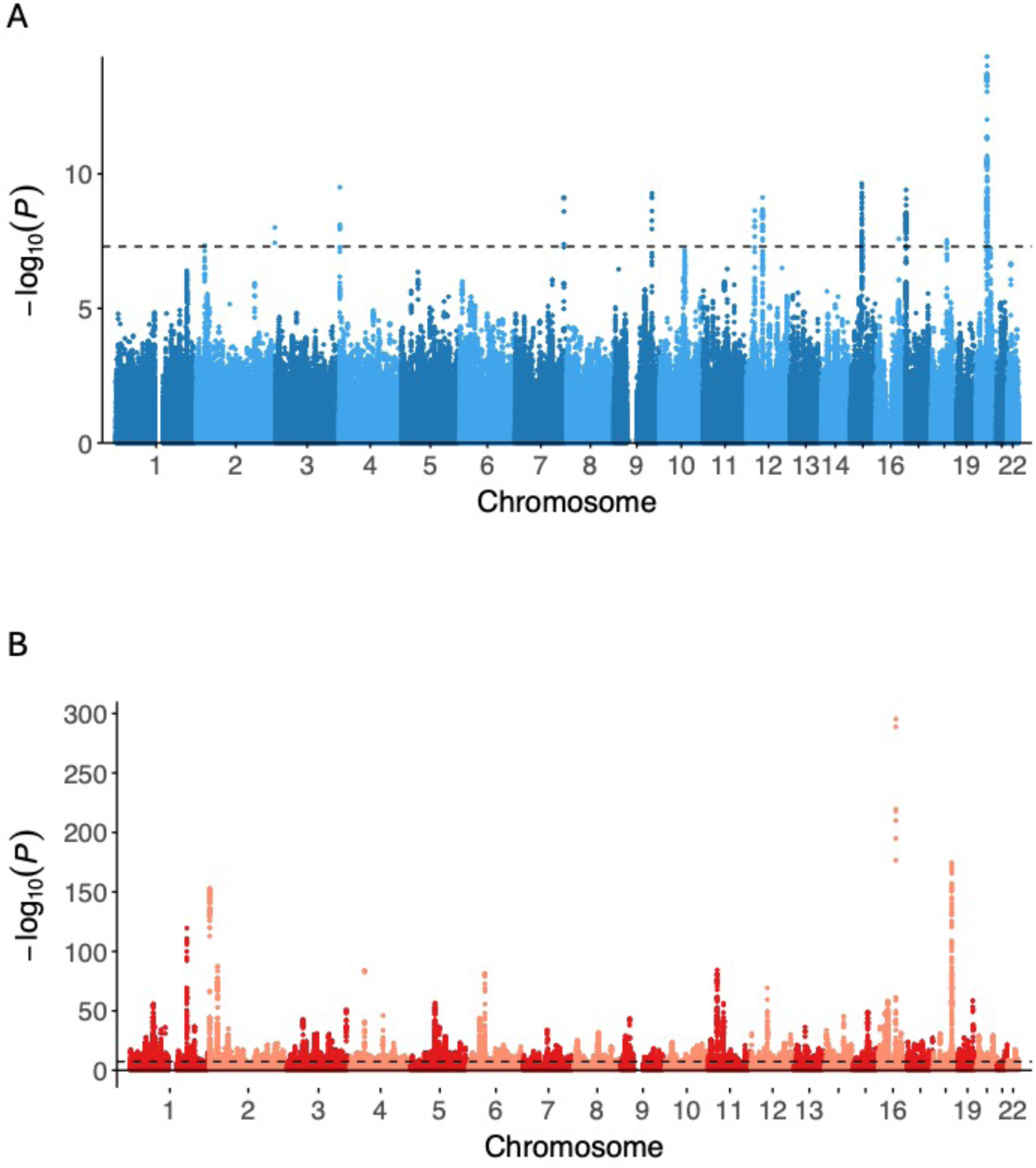
Manhattan plots of non-BMI-mediated and BMI-related GWAS for Total Knee Replacement (TKR). The non-BMI-mediated panels (A) depict associations not fully mediated by BMI while the BMI-related panels (B) depict associations attributable to BMI-related effects. Each point represents a tested variant. Chromosomes are ordered along the x-axis. Variants are plotted by genomic position (x-axis) and −log_10_*P* (y-axis). Genome-wide significance is indicated by the black horizontal line at *P* = 5 × 10^-8^.

**Supplementary Figure 11.**
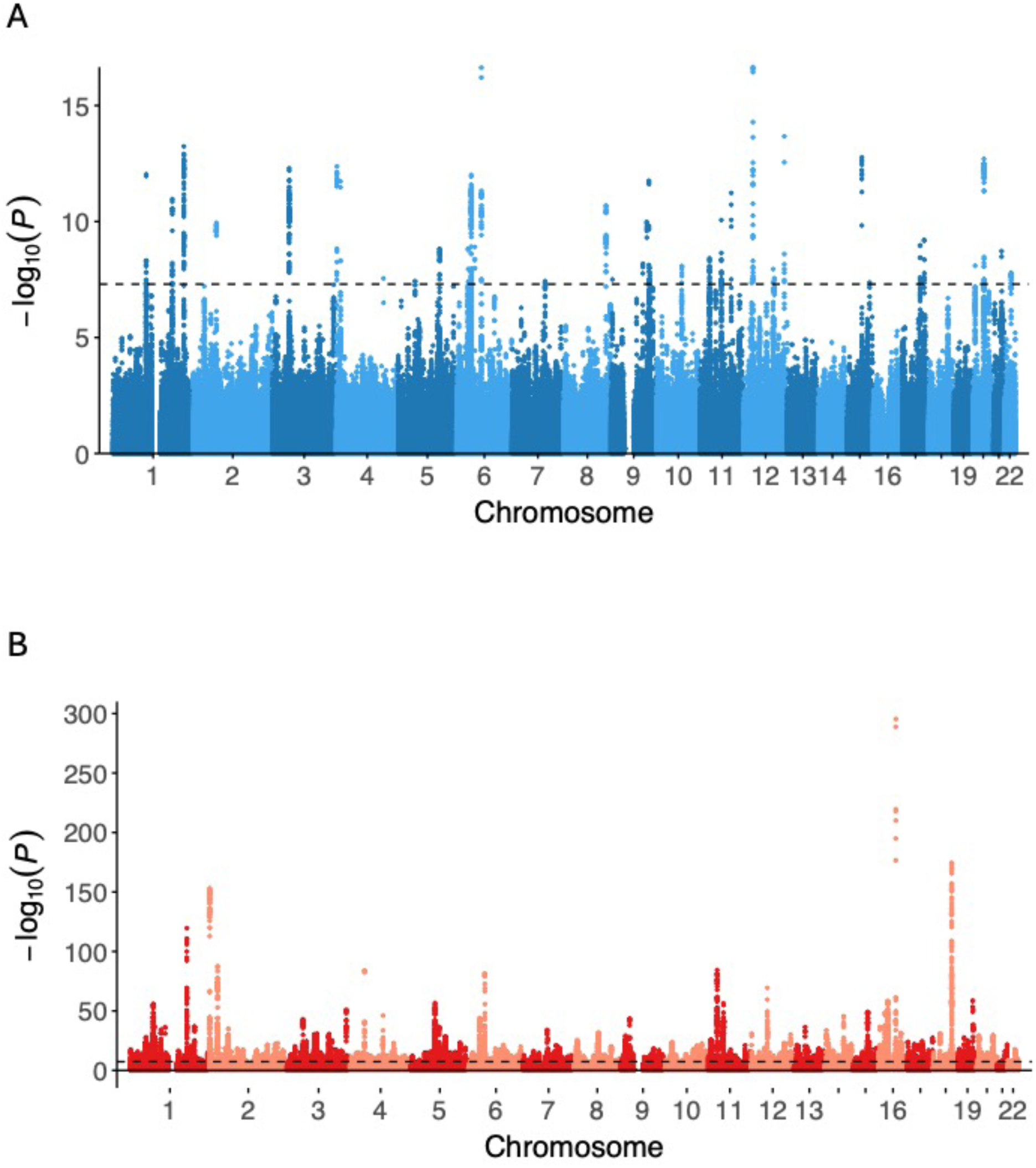
Manhattan plots of non-BMI-mediated and BMI-related GWAS for Total Joint Replacement (TJR). The non-BMI-mediated panels (A) depict associations not fully mediated by BMI while the BMI-related panels (B) depict associations attributable to BMI-related effects. Each point represents a tested variant. Chromosomes are ordered along the x-axis. Variants are plotted by genomic position (x-axis) and −log_10_*P* (y-axis). Genome-wide significance is indicated by the black horizontal line at *P* = 5 × 10^-8^.

**Supplementary Figure 12.**
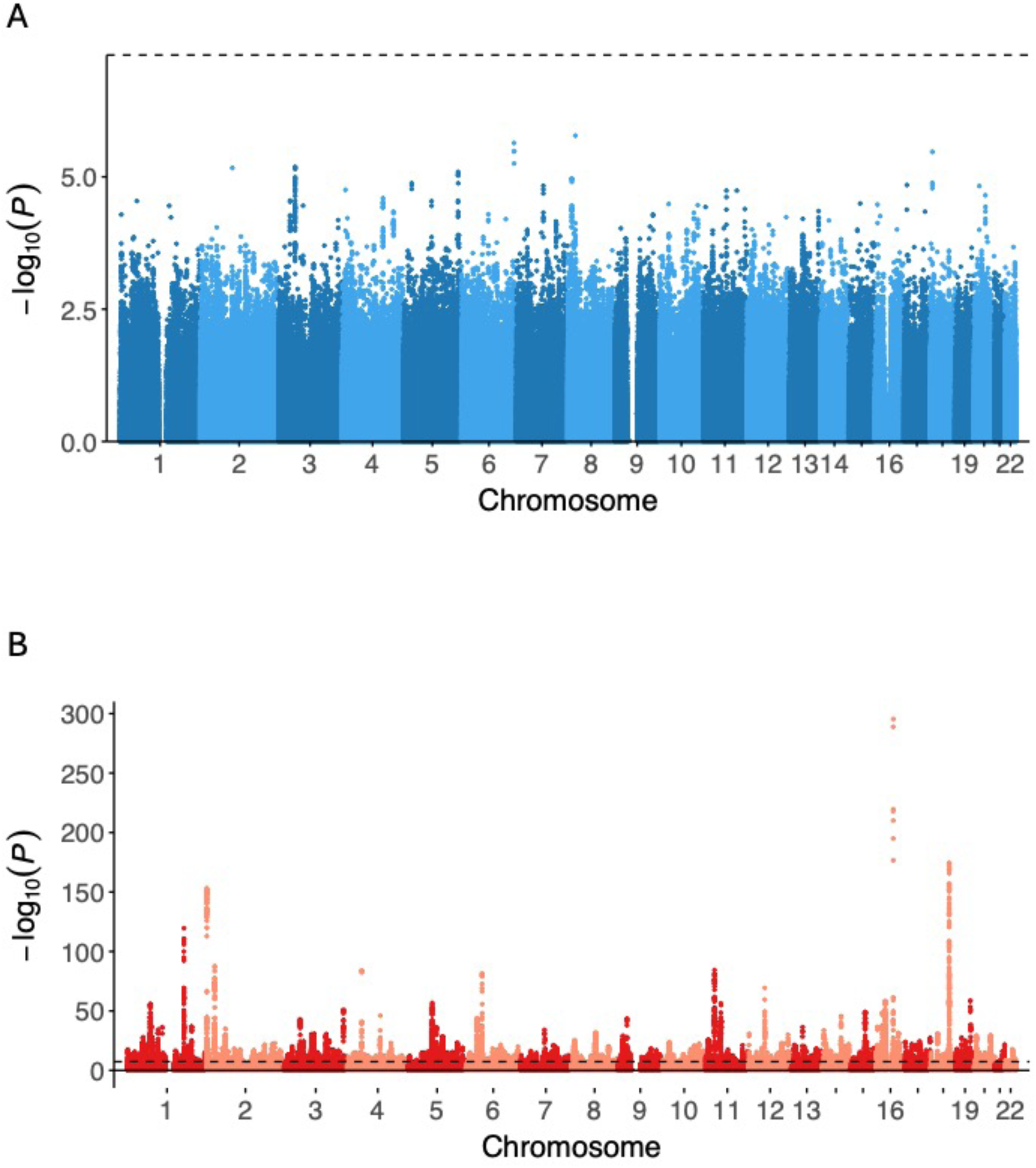
Manhattan plots of non-BMI-mediated and BMI-related GWAS for Early-onset OA. The non-BMI-mediated panels (A) depict associations not fully mediated by BMI while the BMI-related panels (B) depict associations attributable to BMI-related effects. Each point represents a tested variant. Chromosomes are ordered along the x-axis. Variants are plotted by genomic position (x-axis) and −log_10_*P* (y-axis). Genome-wide significance is indicated by the black horizontal line at *P* = 5 × 10^-8^.

**Supplementary Figure 13.**
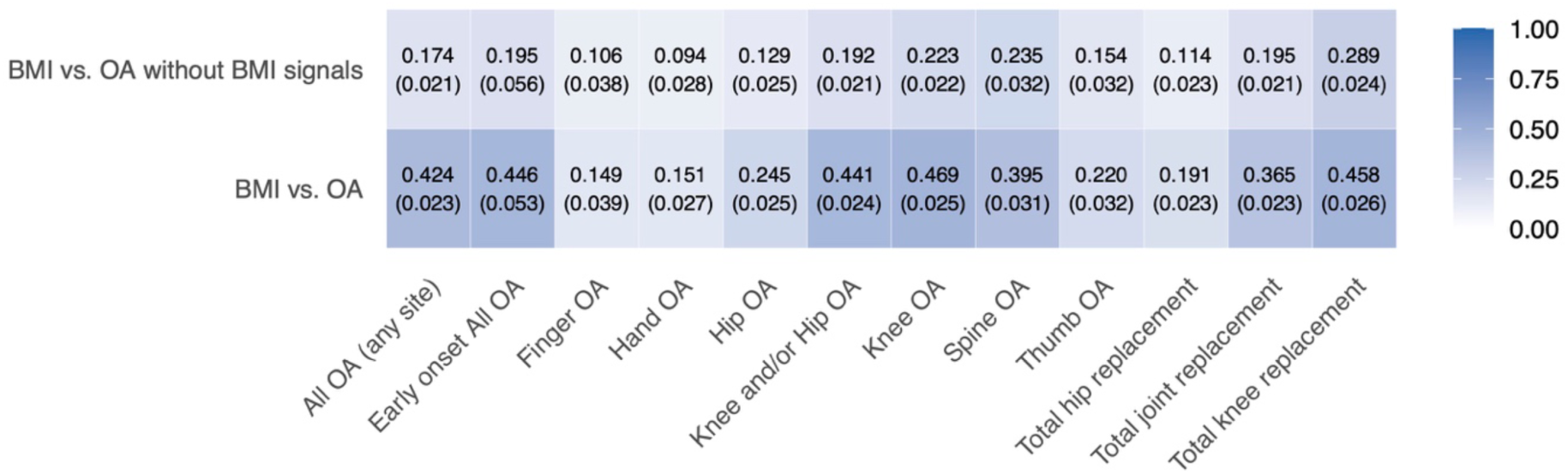
Heatmap of LDSC-estimated genetic correlations between BMI and osteoarthritis (OA) outcomes. The top row (“BMI vs. OA without BMI signals”) shows correlations between BMI and OA phenotypes after removing BMI-mediated genetic signals via GWAS-by-subtraction; the bottom row (“BMI vs. OA”) shows correlations with the original OA GWAS. Columns correspond to OA definitions: All OA (any site), Early-onset All OA, Finger OA, Hand OA, Hip OA, Knee and/or Hip OA, Knee OA, Spine OA, Thumb OA, Total hip replacement, Total joint replacement, and Total knee replacement. Each cell reports the correlation estimate with its standard error (in parentheses). Color scale: white = 0 and darker blue indicates stronger positive correlation (up to 1.0).

**Supplementary Figure 14.**
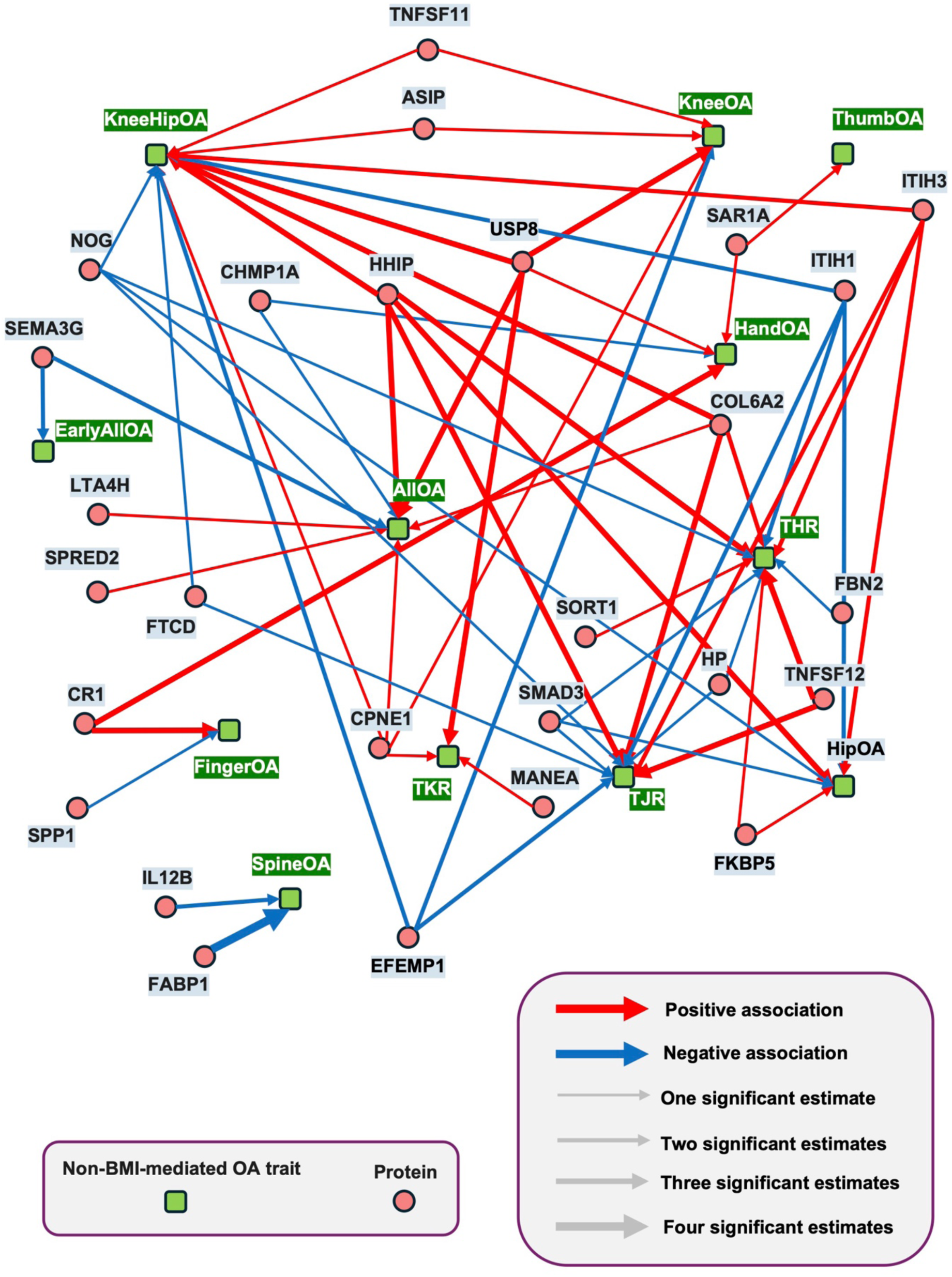
Network plot of putatively causal protein-phenotype associations in non-BMI-mediated osteoarthritis. This network diagram illustrates protein-OA trait pairs identified through Mendelian randomization and colocalization analyses. Green squares represent the non-BMI-mediated OA traits, while pink circles represent proteins. Edges indicate significant associations with MR and colocalization evidence, with red lines denoting positive associations and blue lines negative associations. The thickness of the lines reflects the number of significant causal estimates from studies ranging from one to four, as indicated in the legend. Traits include general and site-specific OA subtypes not mediated by BMI such as OA at any site (AllOA), Knee OA (KneeOA), Knee and/or Hip OA (KneeHipOA), Hip OA (HipOA), Hand OA (HandOA), Spine OA (SpineOA), Finger OA (FingerOA), Thumb OA (ThumbOA), Total Knee Replacement (TKR), Total Hip Replacement (THR), Total Joint Replacement (TJR), and Early-onset OA (EarlyAllOA).

**Supplementary Figure 15.**
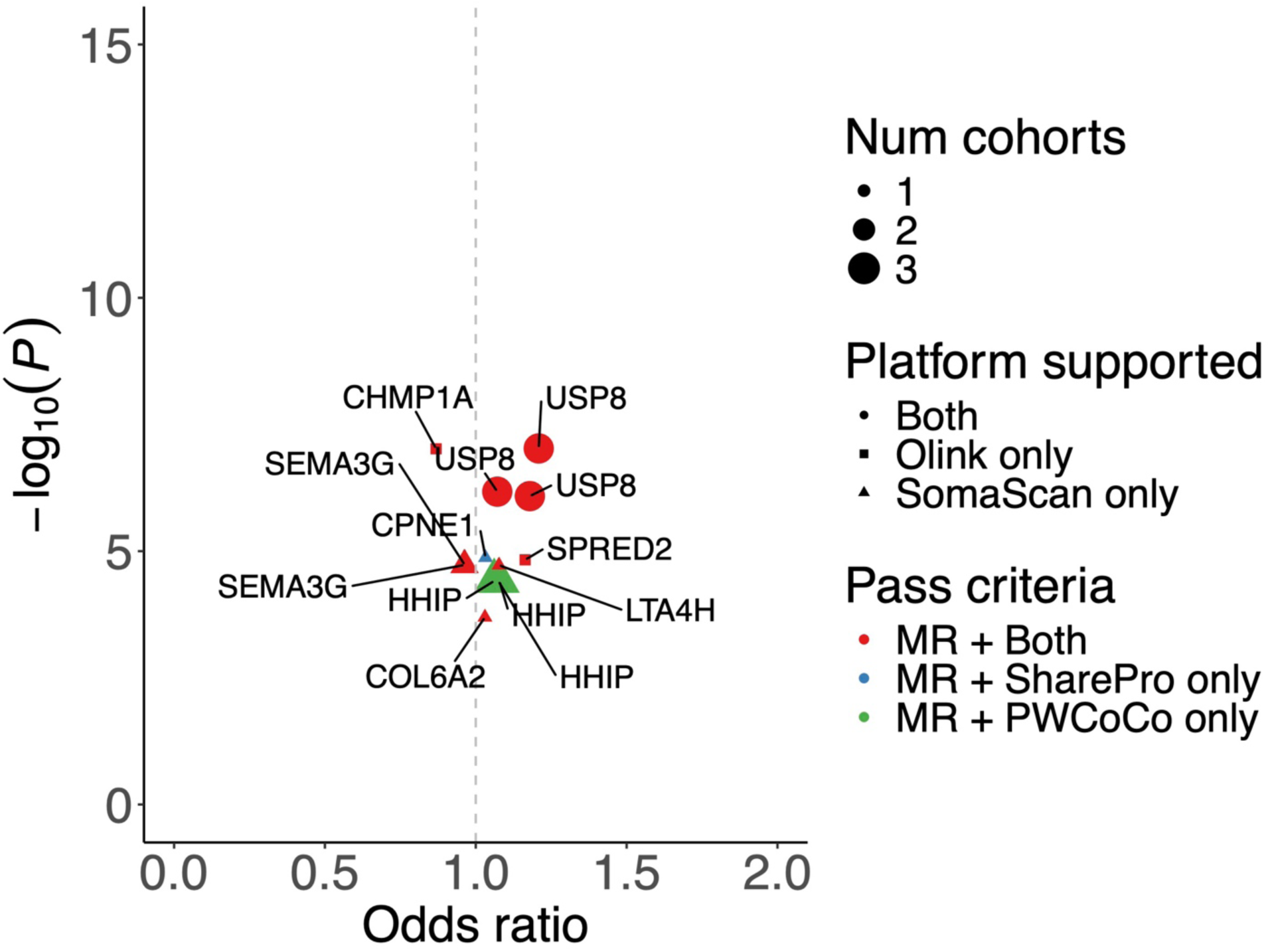
Volcano plots of putatively causal proteins for non-BMI-mediated OA at any site. In the following figures, each panel displays a volcano plot showing the odds ratio (OR) associated with a one standard deviation increase in circulating protein levels on the x-axis versus statistical significance (-log_10_(*P*)) on the y-axis of proteins identified through Mendelian randomization (MR) and colocalization analyses across non-BMI-mediated osteoarthritis traits. Proteins meeting significance thresholds under different criteria (MR + Both, MR + SharePro only, MR + PWCoCo only) are highlighted. MR + Both: proteins with significant MR associations, passing all sensitivity analyses, and colocalizing with a PP.H4 ≥ 0.8 in both PWCoCo and SharePro methods; MR + SharePro only: proteins with significant MR associations, passing all sensitivity analyses, and colocalizing with a PP.H4 ≥ 0.8 in SharePro only; MR + PWCoCo only: proteins with significant MR associations, passing all sensitivity analyses, and colocalizing with a PP.H4 ≥ 0.8 in PWCoCo only. Colors and shapes indicate the number of cohorts supporting the findings and the proteomic platforms (Olink, SomaScan, or both). Abbreviations: Osteoarthritis (OA), Total Hip Replacement (THR), Total Knee Replacement (TKR), Total Joint Replacement (TJR), and Early-onset OA (Early OA).

**Supplementary Figure 16.**
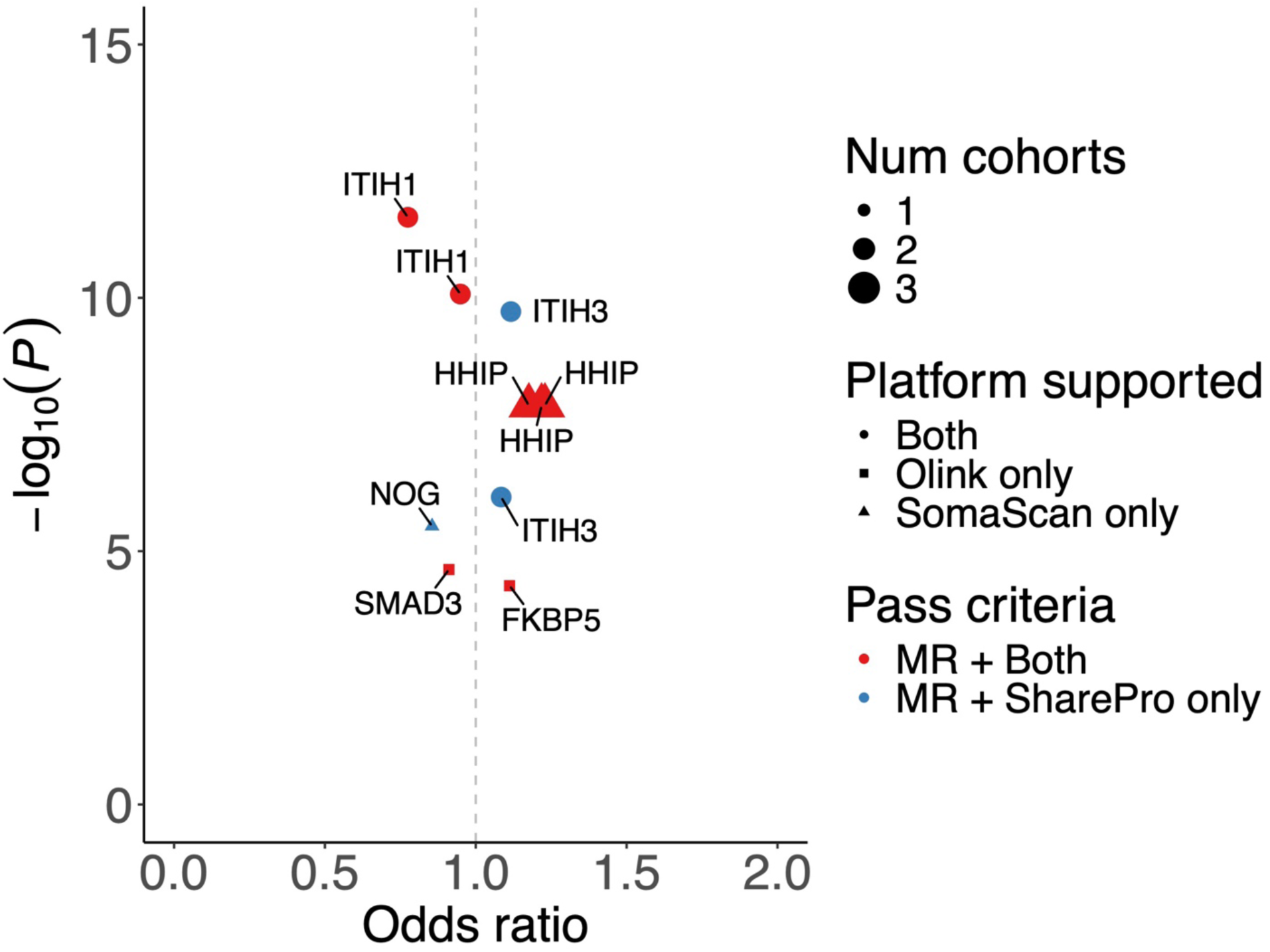
Volcano plots of putatively causal proteins for non-BMI-mediated Hip OA. In the following figures, each panel displays a volcano plot showing the odds ratio (OR) associated with a one standard deviation increase in circulating protein levels on the x-axis versus statistical significance (-log_10_(*P*)) on the y-axis of proteins identified through Mendelian randomization (MR) and colocalization analyses across non-BMI-mediated osteoarthritis traits. Proteins meeting significance thresholds under different criteria (MR + Both, MR + SharePro only, MR + PWCoCo only) are highlighted. MR + Both: proteins with significant MR associations, passing all sensitivity analyses, and colocalizing with a PP.H4 ≥ 0.8 in both PWCoCo and SharePro methods; MR + SharePro only: proteins with significant MR associations, passing all sensitivity analyses, and colocalizing with a PP.H4 ≥ 0.8 in SharePro only; MR + PWCoCo only: proteins with significant MR associations, passing all sensitivity analyses, and colocalizing with a PP.H4 ≥ 0.8 in PWCoCo only. Colors and shapes indicate the number of cohorts supporting the findings and the proteomic platforms (Olink, SomaScan, or both). Abbreviations: Osteoarthritis (OA), Total Hip Replacement (THR), Total Knee Replacement (TKR), Total Joint Replacement (TJR), and Early-onset OA (Early OA).

**Supplementary Figure 17.**
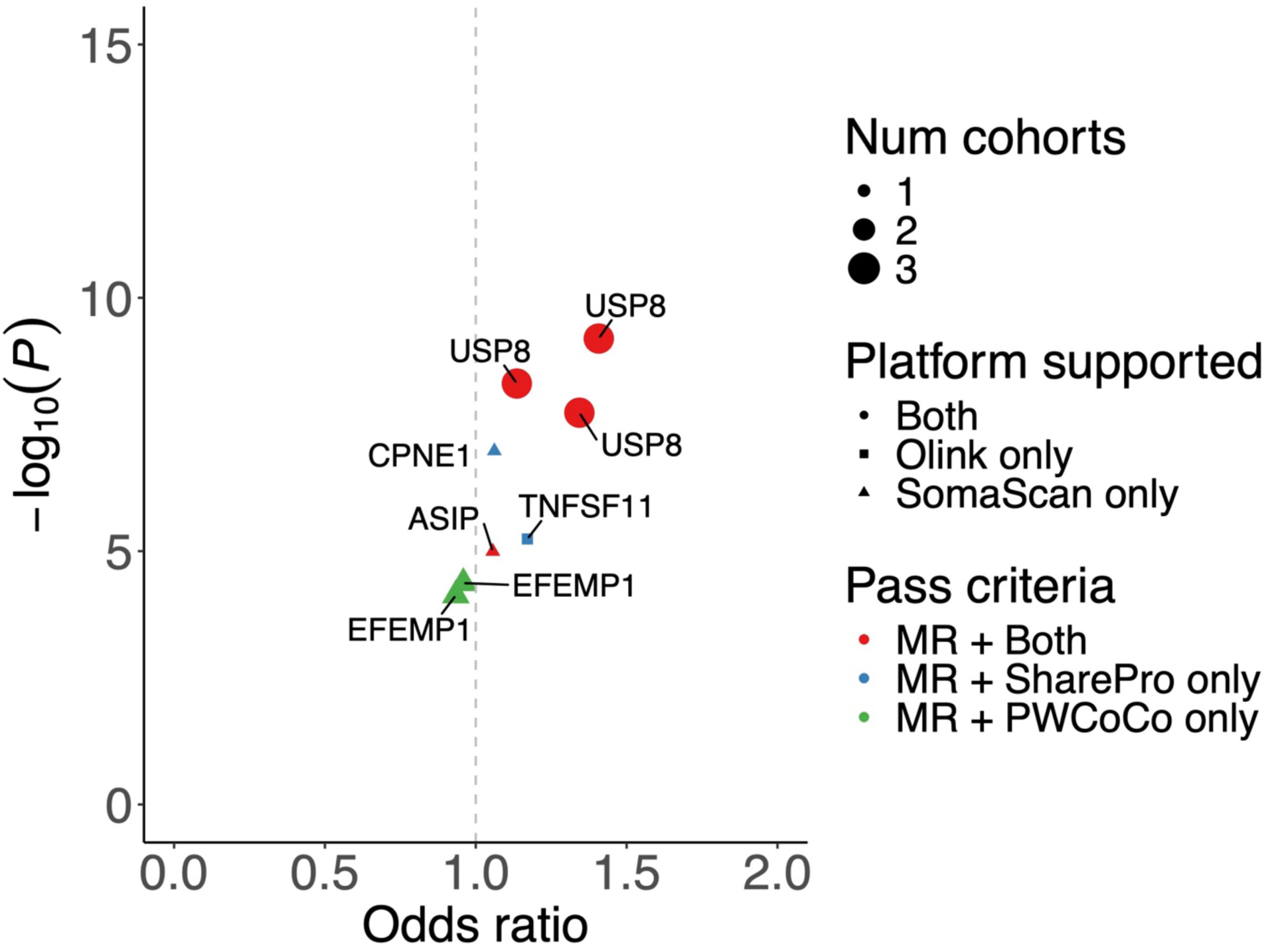
Volcano plots of putatively causal proteins for non-BMI-mediated Knee OA. In the following figures, each panel displays a volcano plot showing the odds ratio (OR) associated with a one standard deviation increase in circulating protein levels on the x-axis versus statistical significance (-log_10_(*P*)) on the y-axis of proteins identified through Mendelian randomization (MR) and colocalization analyses across non-BMI-mediated osteoarthritis traits. Proteins meeting significance thresholds under different criteria (MR + Both, MR + SharePro only, MR + PWCoCo only) are highlighted. MR + Both: proteins with significant MR associations, passing all sensitivity analyses, and colocalizing with a PP.H4 ≥ 0.8 in both PWCoCo and SharePro methods; MR + SharePro only: proteins with significant MR associations, passing all sensitivity analyses, and colocalizing with a PP.H4 ≥ 0.8 in SharePro only; MR + PWCoCo only: proteins with significant MR associations, passing all sensitivity analyses, and colocalizing with a PP.H4 ≥ 0.8 in PWCoCo only. Colors and shapes indicate the number of cohorts supporting the findings and the proteomic platforms (Olink, SomaScan, or both). Abbreviations: Osteoarthritis (OA), Total Hip Replacement (THR), Total Knee Replacement (TKR), Total Joint Replacement (TJR), and Early-onset OA (Early OA).

**Supplementary Figure 18.**
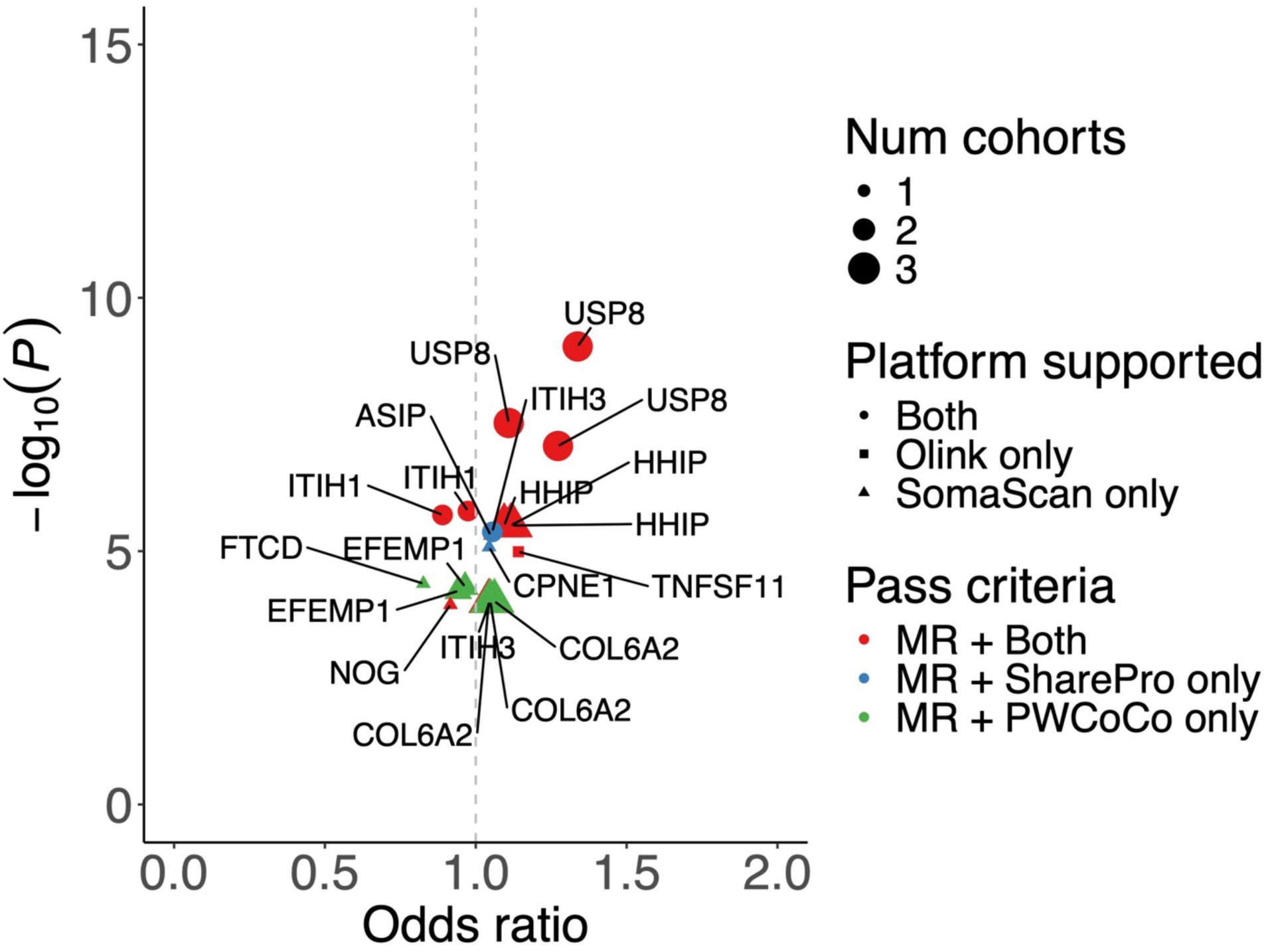
Volcano plots of putatively causal proteins for non-BMI-mediated Knee and/or Hip OA. In the following figures, each panel displays a volcano plot showing the odds ratio (OR) associated with a one standard deviation increase in circulating protein levels on the x-axis versus statistical significance (-log_10_(*P*)) on the y-axis of proteins identified through Mendelian randomization (MR) and colocalization analyses across non-BMI-mediated osteoarthritis traits. Proteins meeting significance thresholds under different criteria (MR + Both, MR + SharePro only, MR + PWCoCo only) are highlighted. MR + Both: proteins with significant MR associations, passing all sensitivity analyses, and colocalizing with a PP.H4 ≥ 0.8 in both PWCoCo and SharePro methods; MR + SharePro only: proteins with significant MR associations, passing all sensitivity analyses, and colocalizing with a PP.H4 ≥ 0.8 in SharePro only; MR + PWCoCo only: proteins with significant MR associations, passing all sensitivity analyses, and colocalizing with a PP.H4 ≥ 0.8 in PWCoCo only. Colors and shapes indicate the number of cohorts supporting the findings and the proteomic platforms (Olink, SomaScan, or both). Abbreviations: Osteoarthritis (OA), Total Hip Replacement (THR), Total Knee Replacement (TKR), Total Joint Replacement (TJR), and Early-onset OA (Early OA).

**Supplementary Figure 19.**
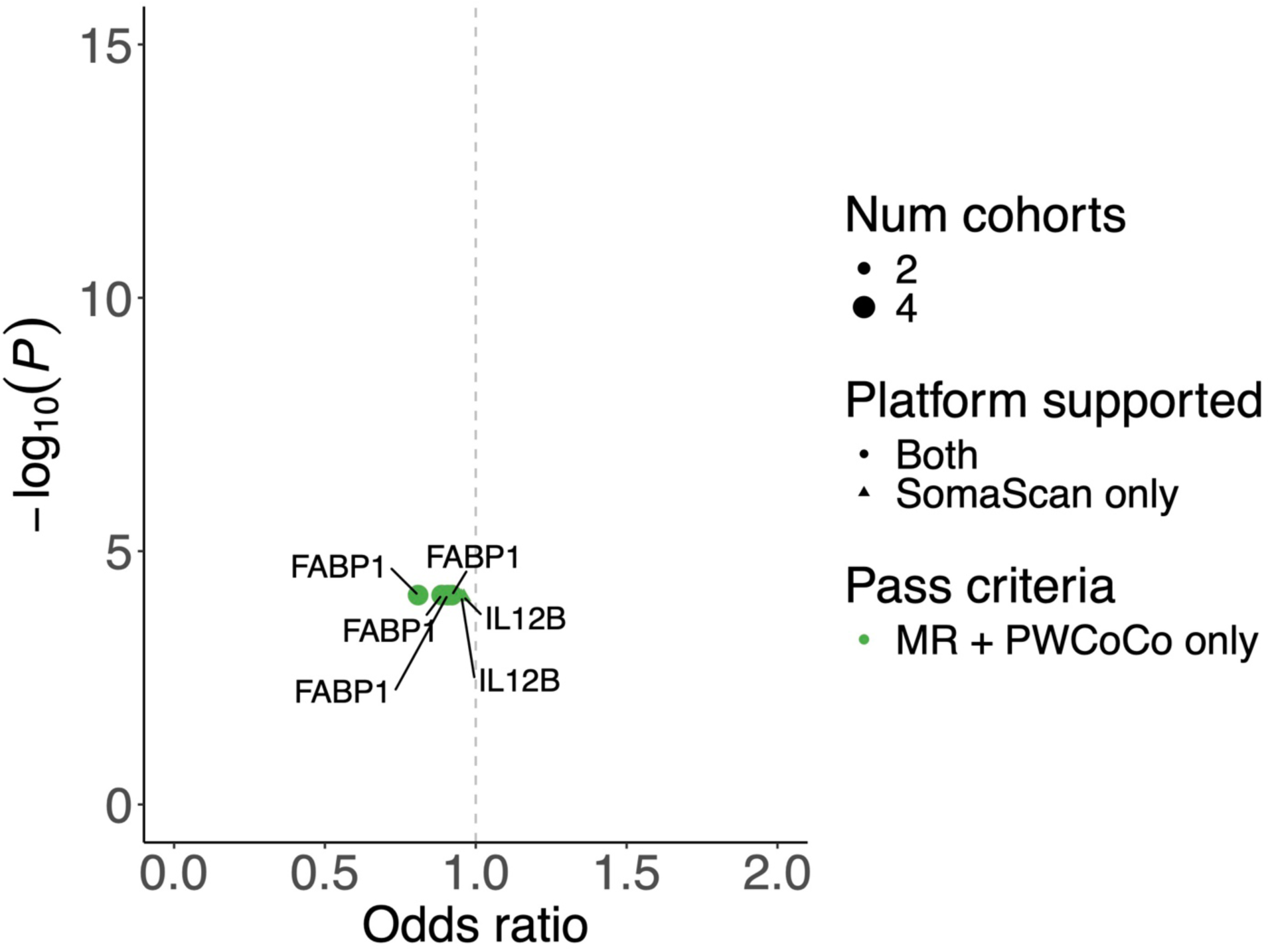
Volcano plots of putatively causal proteins for non-BMI-mediated Spine OA. In the following figures, each panel displays a volcano plot showing the odds ratio (OR) associated with a one standard deviation increase in circulating protein levels on the x-axis versus statistical significance (-log_10_(*P*)) on the y-axis of proteins identified through Mendelian randomization (MR) and colocalization analyses across non-BMI-mediated osteoarthritis traits. Proteins meeting significance thresholds under different criteria (MR + Both, MR + SharePro only, MR + PWCoCo only) are highlighted. MR + Both: proteins with significant MR associations, passing all sensitivity analyses, and colocalizing with a PP.H4 ≥ 0.8 in both PWCoCo and SharePro methods; MR + SharePro only: proteins with significant MR associations, passing all sensitivity analyses, and colocalizing with a PP.H4 ≥ 0.8 in SharePro only; MR + PWCoCo only: proteins with significant MR associations, passing all sensitivity analyses, and colocalizing with a PP.H4 ≥ 0.8 in PWCoCo only. Colors and shapes indicate the number of cohorts supporting the findings and the proteomic platforms (Olink, SomaScan, or both). Abbreviations: Osteoarthritis (OA), Total Hip Replacement (THR), Total Knee Replacement (TKR), Total Joint Replacement (TJR), and Early-onset OA (Early OA).

**Supplementary Figure 20.**
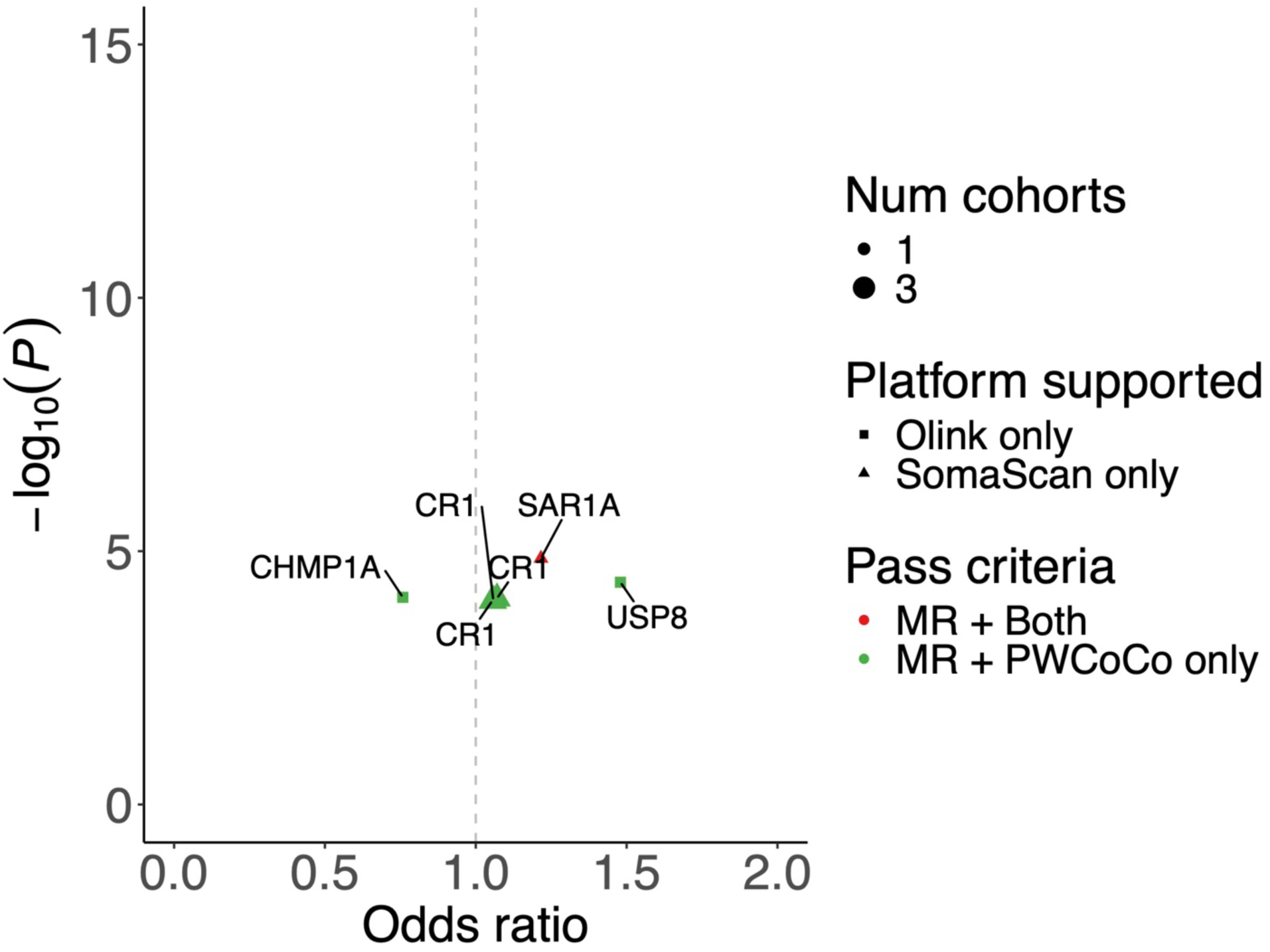
Volcano plots of putatively causal proteins for non-BMI-mediated Hand OA. In the following figures, each panel displays a volcano plot showing the odds ratio (OR) associated with a one standard deviation increase in circulating protein levels on the x-axis versus statistical significance (-log_10_(*P*)) on the y-axis of proteins identified through Mendelian randomization (MR) and colocalization analyses across non-BMI-mediated osteoarthritis traits. Proteins meeting significance thresholds under different criteria (MR + Both, MR + SharePro only, MR + PWCoCo only) are highlighted. MR + Both: proteins with significant MR associations, passing all sensitivity analyses, and colocalizing with a PP.H4 ≥ 0.8 in both PWCoCo and SharePro methods; MR + SharePro only: proteins with significant MR associations, passing all sensitivity analyses, and colocalizing with a PP.H4 ≥ 0.8 in SharePro only; MR + PWCoCo only: proteins with significant MR associations, passing all sensitivity analyses, and colocalizing with a PP.H4 ≥ 0.8 in PWCoCo only. Colors and shapes indicate the number of cohorts supporting the findings and the proteomic platforms (Olink, SomaScan, or both). Abbreviations: Osteoarthritis (OA), Total Hip Replacement (THR), Total Knee Replacement (TKR), Total Joint Replacement (TJR), and Early-onset OA (Early OA).

**Supplementary Figure 21.**
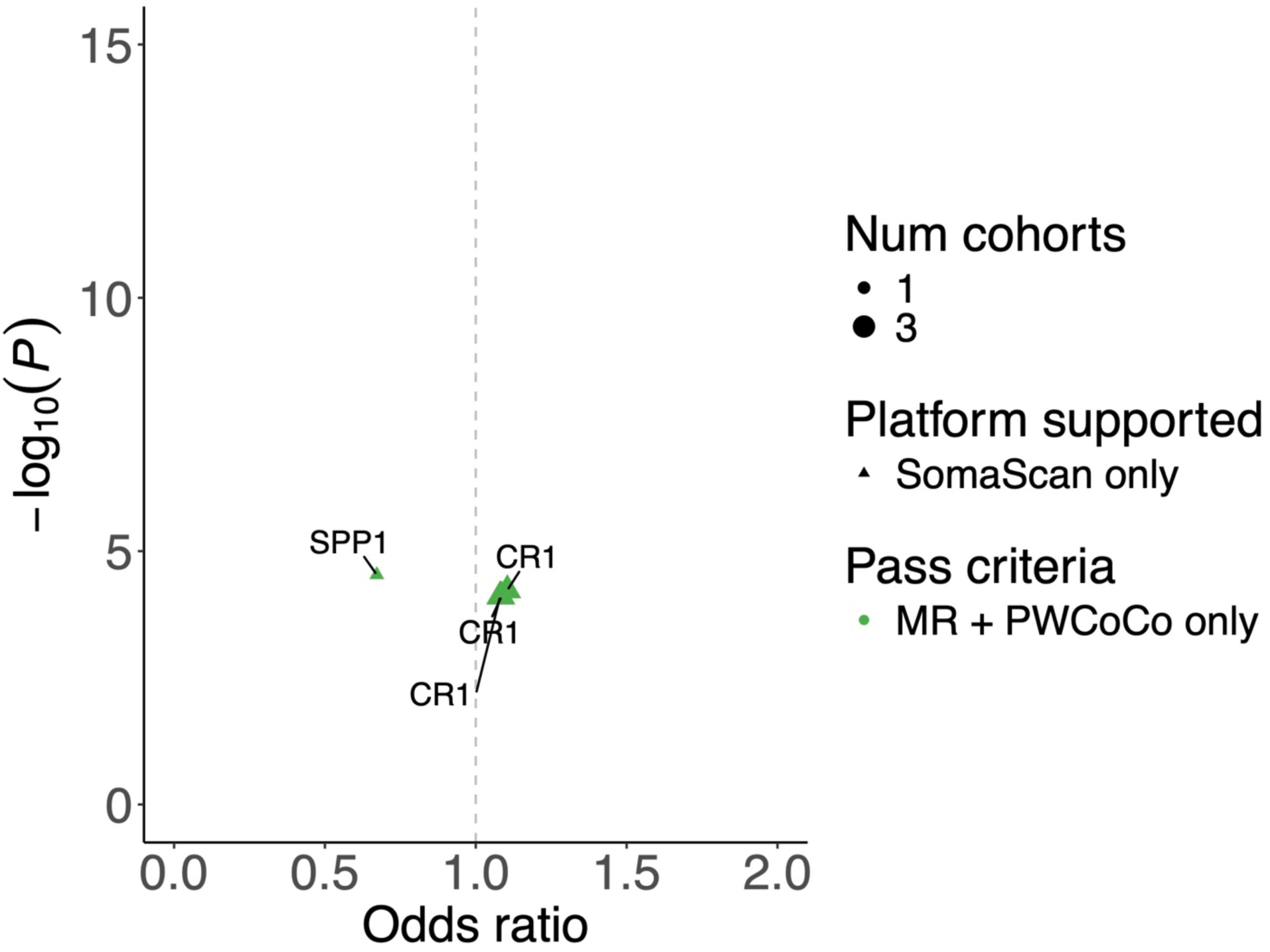
Volcano plots of putatively causal proteins for non-BMI-mediated Finger OA. In the following figures, each panel displays a volcano plot showing the odds ratio (OR) associated with a one standard deviation increase in circulating protein levels on the x-axis versus statistical significance (-log_10_(*P*)) on the y-axis of proteins identified through Mendelian randomization (MR) and colocalization analyses across non-BMI-mediated osteoarthritis traits. Proteins meeting significance thresholds under different criteria (MR + Both, MR + SharePro only, MR + PWCoCo only) are highlighted. MR + Both: proteins with significant MR associations, passing all sensitivity analyses, and colocalizing with a PP.H4 ≥ 0.8 in both PWCoCo and SharePro methods; MR + SharePro only: proteins with significant MR associations, passing all sensitivity analyses, and colocalizing with a PP.H4 ≥ 0.8 in SharePro only; MR + PWCoCo only: proteins with significant MR associations, passing all sensitivity analyses, and colocalizing with a PP.H4 ≥ 0.8 in PWCoCo only. Colors and shapes indicate the number of cohorts supporting the findings and the proteomic platforms (Olink, SomaScan, or both). Abbreviations: Osteoarthritis (OA), Total Hip Replacement (THR), Total Knee Replacement (TKR), Total Joint Replacement (TJR), and Early-onset OA (Early OA).

**Supplementary Figure 22.**
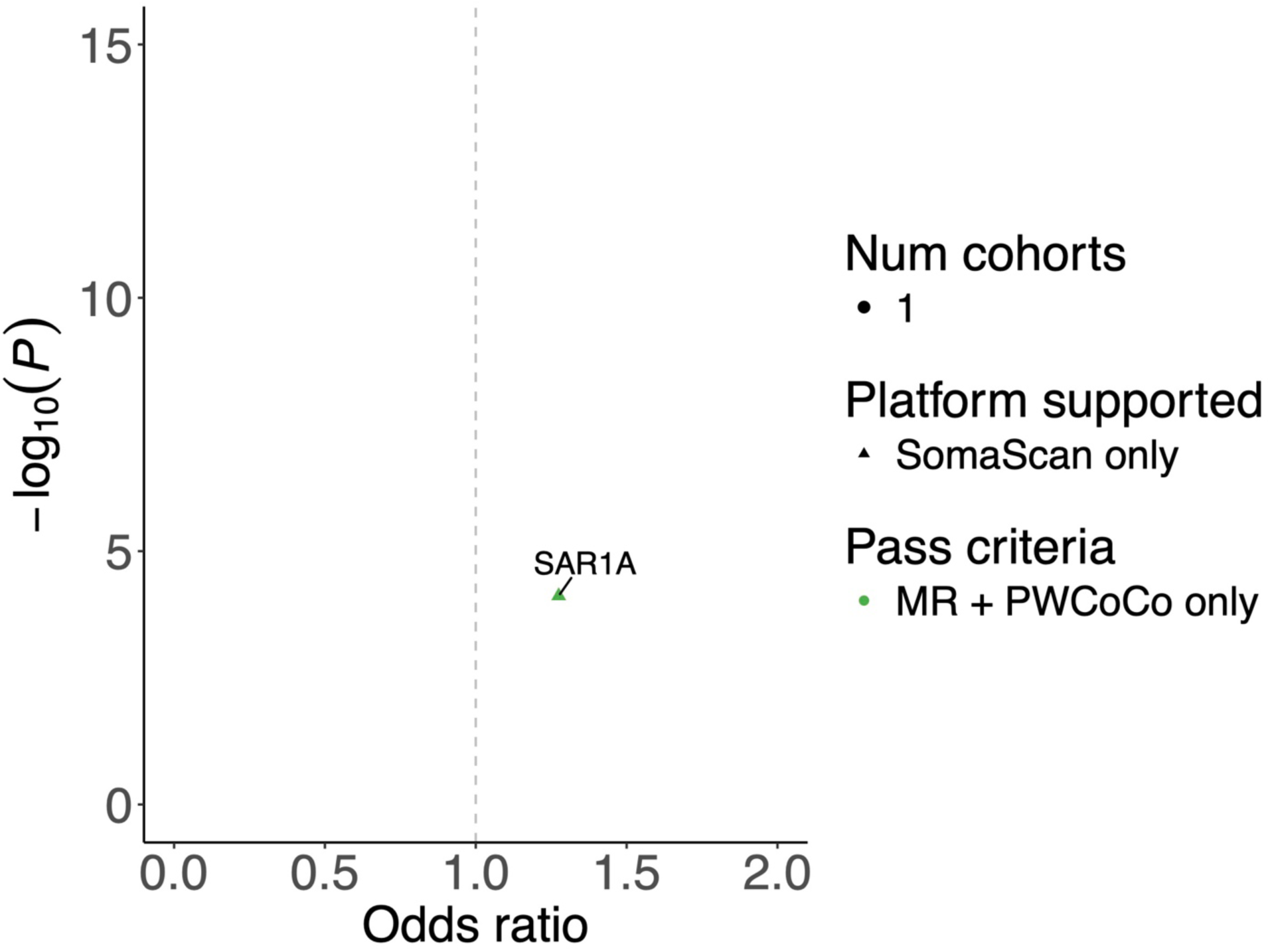
Volcano plots of putatively causal proteins for non-BMI-mediated Thumb OA. In the following figures, each panel displays a volcano plot showing the odds ratio (OR) associated with a one standard deviation increase in circulating protein levels on the x-axis versus statistical significance (-log_10_(*P*)) on the y-axis of proteins identified through Mendelian randomization (MR) and colocalization analyses across non-BMI-mediated osteoarthritis traits. Proteins meeting significance thresholds under different criteria (MR + Both, MR + SharePro only, MR + PWCoCo only) are highlighted. MR + Both: proteins with significant MR associations, passing all sensitivity analyses, and colocalizing with a PP.H4 ≥ 0.8 in both PWCoCo and SharePro methods; MR + SharePro only: proteins with significant MR associations, passing all sensitivity analyses, and colocalizing with a PP.H4 ≥ 0.8 in SharePro only; MR + PWCoCo only: proteins with significant MR associations, passing all sensitivity analyses, and colocalizing with a PP.H4 ≥ 0.8 in PWCoCo only. Colors and shapes indicate the number of cohorts supporting the findings and the proteomic platforms (Olink, SomaScan, or both). Abbreviations: Osteoarthritis (OA), Total Hip Replacement (THR), Total Knee Replacement (TKR), Total Joint Replacement (TJR), and Early-onset OA (Early OA).

**Supplementary Figure 23.**
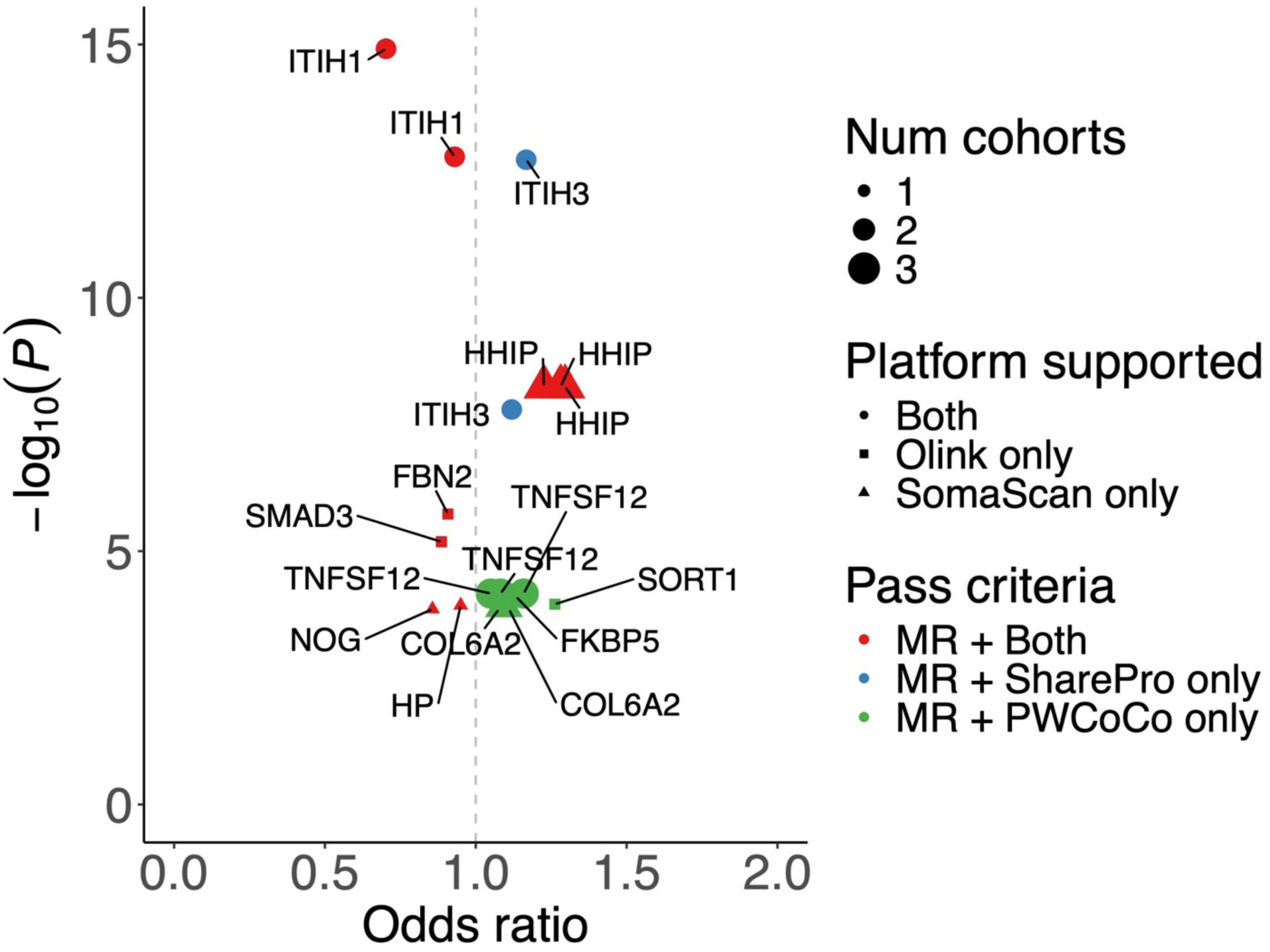
Volcano plots of putatively causal proteins for non-BMI-mediated Total Hip Replacement (THR). In the following figures, each panel displays a volcano plot showing the odds ratio (OR) associated with a one standard deviation increase in circulating protein levels on the x-axis versus statistical significance (-log_10_(*P*)) on the y-axis of proteins identified through Mendelian randomization (MR) and colocalization analyses across non-BMI-mediated osteoarthritis traits. Proteins meeting significance thresholds under different criteria (MR + Both, MR + SharePro only, MR + PWCoCo only) are highlighted. MR + Both: proteins with significant MR associations, passing all sensitivity analyses, and colocalizing with a PP.H4 ≥ 0.8 in both PWCoCo and SharePro methods; MR + SharePro only: proteins with significant MR associations, passing all sensitivity analyses, and colocalizing with a PP.H4 ≥ 0.8 in SharePro only; MR + PWCoCo only: proteins with significant MR associations, passing all sensitivity analyses, and colocalizing with a PP.H4 ≥ 0.8 in PWCoCo only. Colors and shapes indicate the number of cohorts supporting the findings and the proteomic platforms (Olink, SomaScan, or both). Abbreviations: Osteoarthritis (OA), Total Hip Replacement (THR), Total Knee Replacement (TKR), Total Joint Replacement (TJR), and Early-onset OA (Early OA).

**Supplementary Figure 24.**
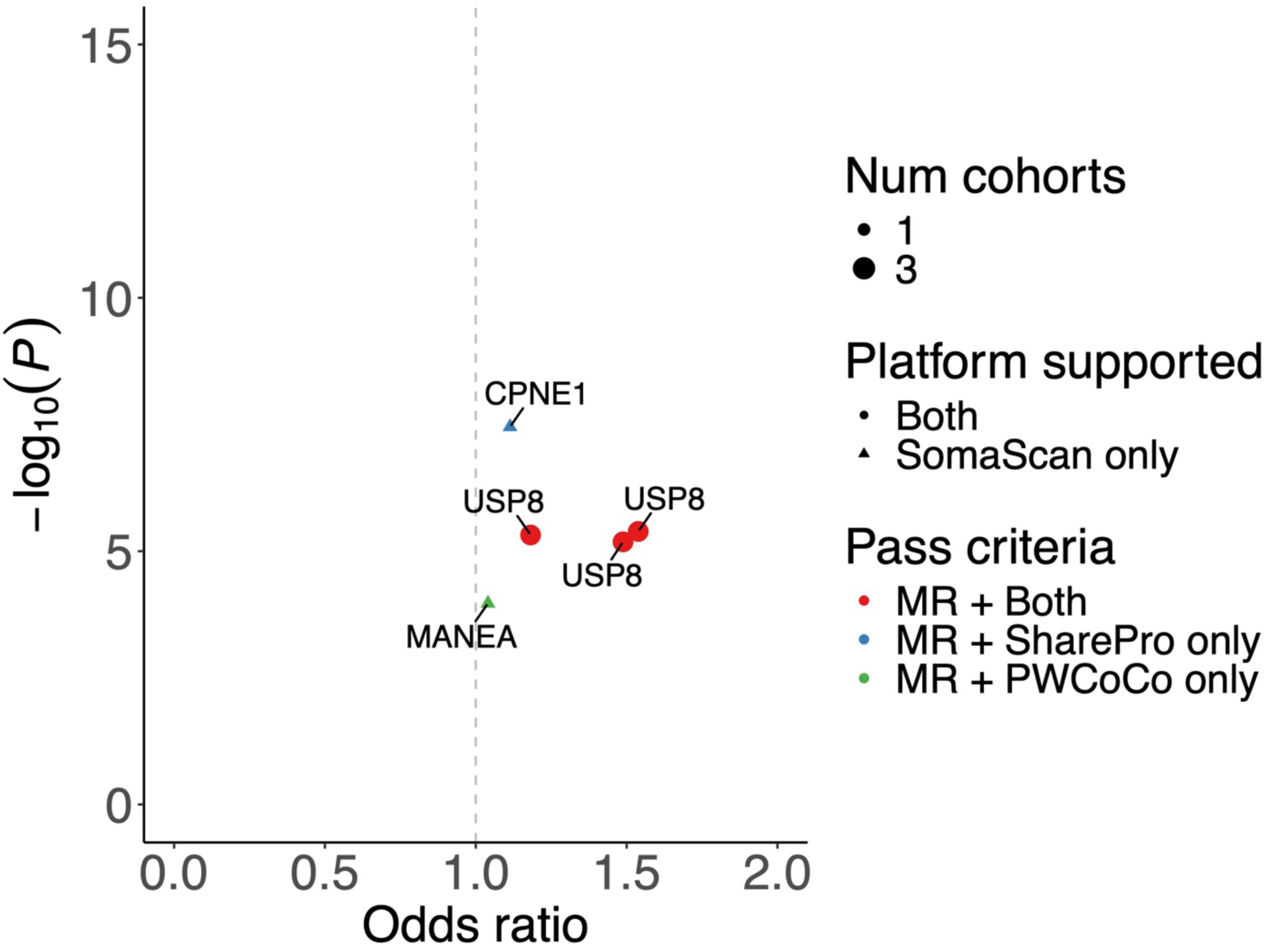
Volcano plots of putatively causal proteins for non-BMI-mediated Total Knee Replacement (TKR). In the following figures, each panel displays a volcano plot showing the odds ratio (OR) associated with a one standard deviation increase in circulating protein levels on the x-axis versus statistical significance (-log_10_(*P*)) on the y-axis of proteins identified through Mendelian randomization (MR) and colocalization analyses across non-BMI-mediated osteoarthritis traits. Proteins meeting significance thresholds under different criteria (MR + Both, MR + SharePro only, MR + PWCoCo only) are highlighted. MR + Both: proteins with significant MR associations, passing all sensitivity analyses, and colocalizing with a PP.H4 ≥ 0.8 in both PWCoCo and SharePro methods; MR + SharePro only: proteins with significant MR associations, passing all sensitivity analyses, and colocalizing with a PP.H4 ≥ 0.8 in SharePro only; MR + PWCoCo only: proteins with significant MR associations, passing all sensitivity analyses, and colocalizing with a PP.H4 ≥ 0.8 in PWCoCo only. Colors and shapes indicate the number of cohorts supporting the findings and the proteomic platforms (Olink, SomaScan, or both). Abbreviations: Osteoarthritis (OA), Total Hip Replacement (THR), Total Knee Replacement (TKR), Total Joint Replacement (TJR), and Early-onset OA (Early OA).

**Supplementary Figure 25.**
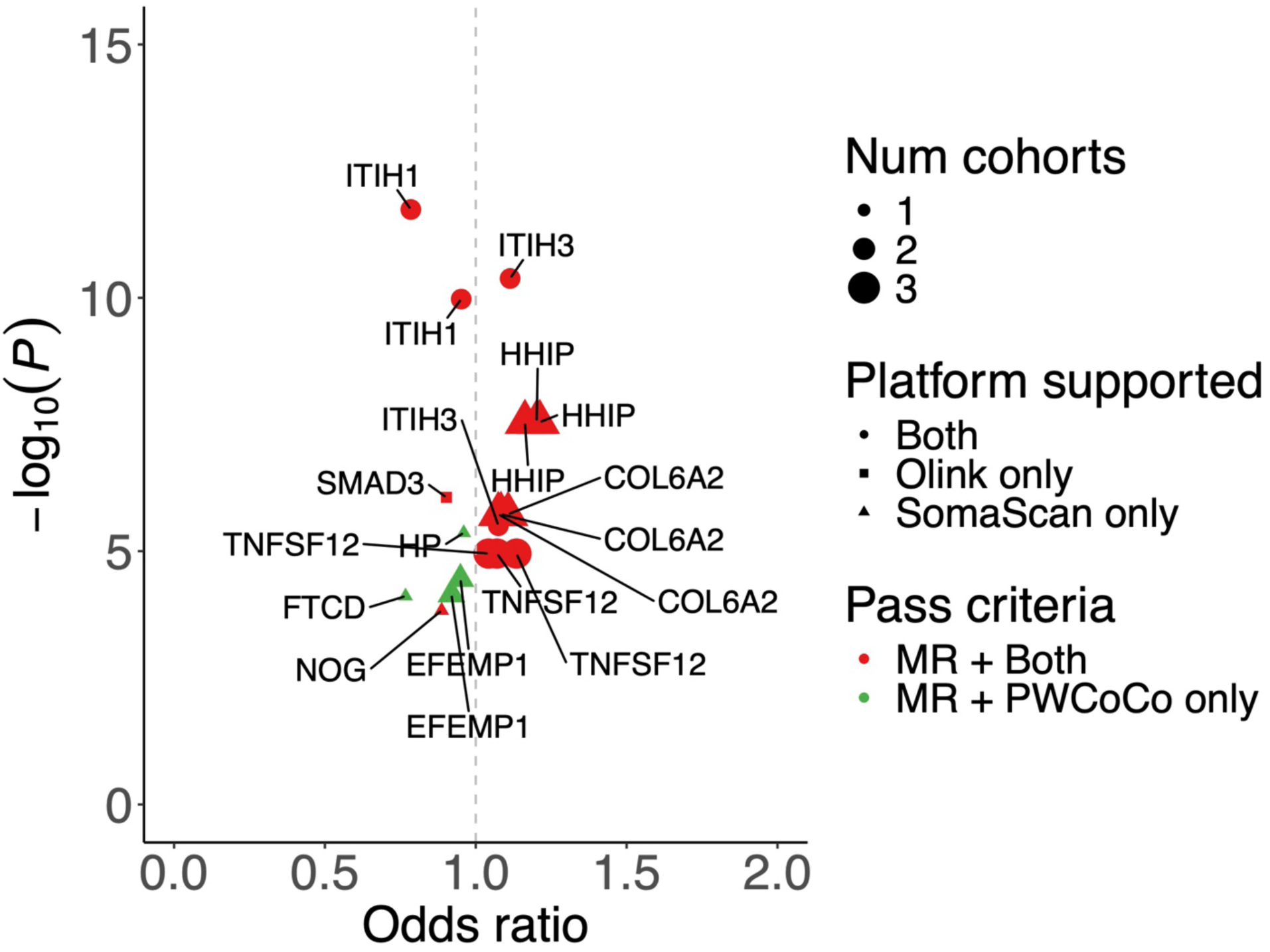
Volcano plots of putatively causal proteins for non-BMI-mediated Total Joint Replacement (TJR). In the following figures, each panel displays a volcano plot showing the odds ratio (OR) associated with a one standard deviation increase in circulating protein levels on the x-axis versus statistical significance (-log_10_(*P*)) on the y-axis of proteins identified through Mendelian randomization (MR) and colocalization analyses across non-BMI-mediated osteoarthritis traits. Proteins meeting significance thresholds under different criteria (MR + Both, MR + SharePro only, MR + PWCoCo only) are highlighted. MR + Both: proteins with significant MR associations, passing all sensitivity analyses, and colocalizing with a PP.H4 ≥ 0.8 in both PWCoCo and SharePro methods; MR + SharePro only: proteins with significant MR associations, passing all sensitivity analyses, and colocalizing with a PP.H4 ≥ 0.8 in SharePro only; MR + PWCoCo only: proteins with significant MR associations, passing all sensitivity analyses, and colocalizing with a PP.H4 ≥ 0.8 in PWCoCo only. Colors and shapes indicate the number of cohorts supporting the findings and the proteomic platforms (Olink, SomaScan, or both). Abbreviations: Osteoarthritis (OA), Total Hip Replacement (THR), Total Knee Replacement (TKR), Total Joint Replacement (TJR), and Early-onset OA (Early OA).

**Supplementary Figure 26.**
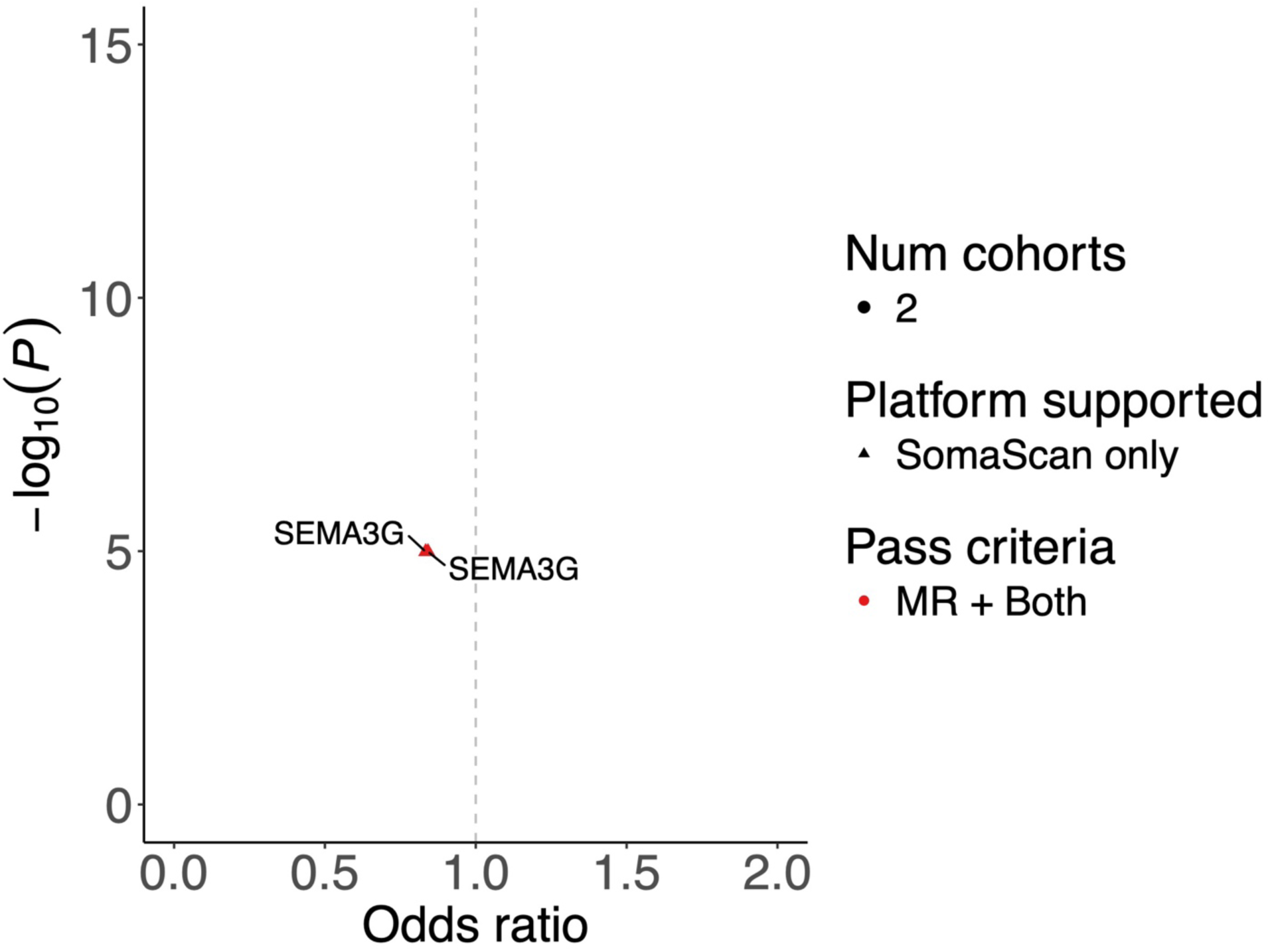
Volcano plots of putatively causal proteins for non-BMI-mediated Early-onset OA. In the following figures, each panel displays a volcano plot showing the odds ratio (OR) associated with a one standard deviation increase in circulating protein levels on the x-axis versus statistical significance (-log_10_(*P*)) on the y-axis of proteins identified through Mendelian randomization (MR) and colocalization analyses across non-BMI-mediated osteoarthritis traits. Proteins meeting significance thresholds under different criteria (MR + Both, MR + SharePro only, MR + PWCoCo only) are highlighted. MR + Both: proteins with significant MR associations, passing all sensitivity analyses, and colocalizing with a PP.H4 ≥ 0.8 in both PWCoCo and SharePro methods; MR + SharePro only: proteins with significant MR associations, passing all sensitivity analyses, and colocalizing with a PP.H4 ≥ 0.8 in SharePro only; MR + PWCoCo only: proteins with significant MR associations, passing all sensitivity analyses, and colocalizing with a PP.H4 ≥ 0.8 in PWCoCo only. Colors and shapes indicate the number of cohorts supporting the findings and the proteomic platforms (Olink, SomaScan, or both). Abbreviations: Osteoarthritis (OA), Total Hip Replacement (THR), Total Knee Replacement (TKR), Total Joint Replacement (TJR), and Early-onset OA (Early OA).

## Supplementary Note 1. Assessing robust evidence of target proteins implicated in non-BMI-mediated osteoarthritis

In order to prioritize potential targets implicated in non-BMI-mediated osteoarthritis (OA), we performed a series of analyses to evaluate the strength of evidence. The criterion we used is summarized in the following table:

| Criterion | Reasoning |
| --- | --- |
| 1. **Statistical robustness:** Mendelian randomization and strong colocalization evidence (PPH4 $\geq 0.8$ in both PWCoCo and SharePro) | Minimizes false positives driven by horizontal pleiotropy or confounding due to linkage disequilibrium |
| 2. **Replication** across<br>• **OA phenotypes** (12 phenotypes considered)<br>• **Cohorts** (4 proteomics cohorts considered)<br>• **Proteomics platform** (2 proteomics platforms considered) | Targets observed in more than one OA anatomic site, and replicated on two proteomic platforms are unlikely to be artifacts and more likely to be implicated in OA pathophysiology |

We found that 18 of 27 protein targets had robust evidence and satisfied Criteria 1. Of these targets, we classified them into three different priority levels (with lower Priority Level indicating greater replication evidence).

### Priority Level 1 targets

- Present in at least one OA trait, with evidence from at least 2 cohorts, and from both SomaScan and Olink platforms

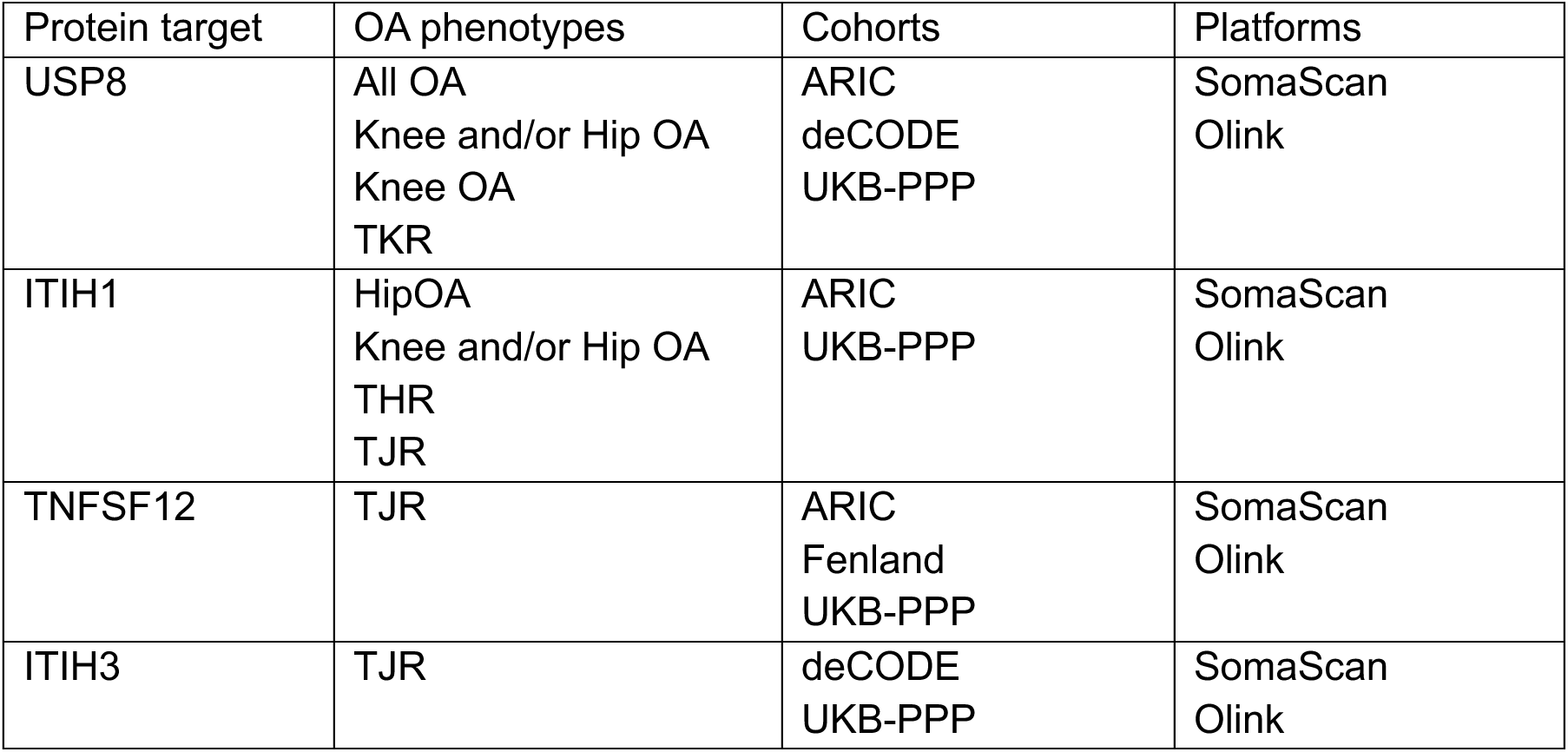

| Protein target | OA phenotypes | Cohorts | Platforms |
| --- | --- | --- | --- |
| USP8 | All OA<br>Knee and/or Hip OA<br>Knee OA<br>TKR | ARIC<br>deCODE<br>UKB-PPP | SomaScan<br>Olink |
| ITIH1 | HipOA<br>Knee and/or Hip OA<br>THR<br>TJR | ARIC<br>UKB-PPP | SomaScan<br>Olink |
| TNFSF12 | TJR | ARIC<br>Fenland<br>UKB-PPP | SomaScan<br>Olink |
| ITIH3 | TJR | deCODE<br>UKB-PPP | SomaScan<br>Olink |

### Priority Level 2 targets

- Present in at least one OA trait, with evidence from at least one cohort

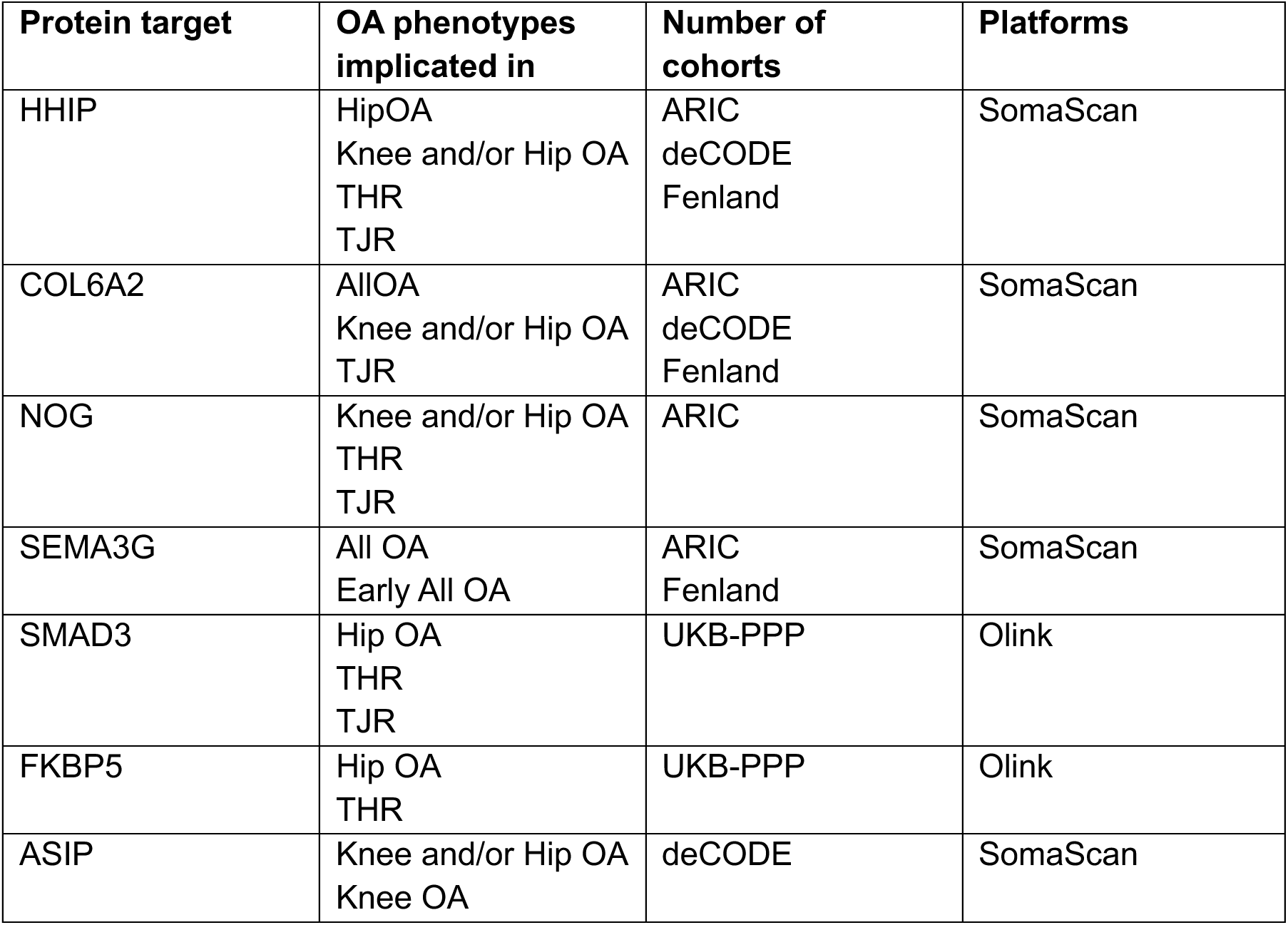

### Priority Level 3 targets

- Present in at least one OA trait, with evidence from one cohort

**Priority Level 3 targets:** - Present in at least one OA trait, with evidence from one cohort
| **Protein target** | **OA phenotypes implicated in** | **Number of cohorts** | **Platforms** |
| --- | --- | --- | --- |
| HP | THR | ARIC | SomaScan |
| LTA4H | All OA | ARIC | SomaScan |
| SAR1A | Hand OA | deCODE | SomaScan |
| CHMP1A | All OA | UKB-PPP | Olink |
| SPRED2 | All OA | UKB-PPP | Olink |
| FBN2 | THR | UKB-PPP | Olink |
| TNFSF11 | Knee and/or Hip OA | UKB-PPP | Olink |

## Supplementary Note 2. Classification of target proteins around four axes

We provide a more detailed description of each protein target represented in Figure 4A below.

1. **Inflammation and Immune Modulation** These proteins are involved in immune signaling, cytokine production, and inflammation regulation—central to osteoarthritis (OA) pathogenesis.

- **FKBP5** – Modulates glucocorticoid receptor sensitivity; involved in stress and inflammation responses^1^.
- **LTA4H** – Leukotriene pathway; regulates pro-inflammatory leukotriene B4^2^.
- **IL12B** – Encodes a subunit of interleukin (IL)-12 and IL-23; bridges innate and adaptive immunity^3^.
- **TNFSF12 (TWEAK)** – Implicated in tissue remodeling and inflammation^4^.
- **CR1** – Complement receptor; involved in immune complex clearance^5^.
- **HP (Haptoglobin)** – Acute-phase protein; involved in scavenging free hemoglobin, oxidative stress^6^.
- **SPRED2** – Negative regulator of MAPK signaling, with roles in immune regulation^7^.
- **ASIP** – Antagonist of melanocortin receptors (especially MC1R and MC4R)^8^.
2. **Extracellular Matrix (ECM), Cartilage, and Bone Remodeling** These proteins influence the structural integrity of connective tissue, cartilage, and bone.

- **TNFSF11 (RANKL)** – central mediator that drives osteoclast differentiation and activates bone resorption^9^.
- **SPP1 (Osteopontin)** – Extracellular matrix (ECM) protein with roles in bone remodeling and inflammation^10^.
- **SEMA3G** – Involved in axon guidance^11^; may play a role in cartilage innervation and remodeling^12^.
- **ITIH1**, **ITIH3** – Part of the inter-alpha-trypsin inhibitor family; stabilize ECM, especially in cartilage^13^.
- **SMAD3** – a key intracellular signaling mediator in the transforming growth factor-beta (TGF-β) pathway, which plays a central role in cartilage homeostasis, ECM remodeling, and chondrocyte differentiation^14^. Dysregulation of SMAD3 signaling has been implicated in the pathogenesis of OA, particularly through its influence on matrix synthesis and degradation^15^.
- **EFEMP1 (Fibulin-3)** – ECM glycoprotein; important in matrix integrity and angiogenesis^16^.
- **FBN2** – Fibrillin; structural protein in ECM^17^.
- **COL6A2** – Collagen; structural protein in cartilage^18^.
- **HHIP** – Regulates Hedgehog signaling; involved in chondrocyte and cartilage development^19^.
- **NOG (Noggin)** – Bone morphogenetic protein inhibitor; regulates cartilage and bone development^20^.
3. **Metabolism and Cellular Stress** These are proteins that mediate metabolic processes, oxidative stress, and cellular response mechanisms.

- **MANEA** – Involved in glycoprotein processing in the endoplasmic reticulum (ER); protein quality control^21^.
- **USP8** – Deubiquitinase; regulates protein degradation and receptor recycling^22^.
- **CPNE1 (Copine-1)** – Calcium-dependent membrane-binding protein; signaling and stress responses^23^.
4. **Vesicular Transport, Protein Trafficking, and Autophagy** These proteins are linked to intracellular trafficking and autophagy which are important in cartilage cell survival and turnover.

- **SAR1A** – Small GTPase; regulates ER-to-Golgi vesicle transport^24^.
- **CHMP1A** – Involved in endosomal sorting (endosomal sorting complexes required for transport also known as endosomal sorting complexes required for transport complex); autophagy and cellular waste disposal^25^.
- **SORT1** – Involved in protein sorting and trafficking^26^; may influence cartilage maintenance^27^.
- **FABP1** – Fatty acid binding protein; involved in lipid metabolism^28^.
- **FTCD** – Involved in histidine and folate metabolism^29^.

### Methods

A set of 27 protein targets was identified as being associated with osteoarthritis through pathways not fully mediated by BMI. To explore the potential biological mechanisms underlying these associations, genes were categorized based on their most likely roles in relevant pathophysiological pathways. Categorization was performed using a combination of literature review and functional annotation with the assistance of ChatGPT5 Thinking (OpenAI, September 2025), to group the protein-coding genes into four primary biological processes: (1) Inflammation & Immune Modulation, (2) Extracellular Matrix (ECM), Cartilage, and Bone Remodeling, (3) Vesicular Transport, Protein Trafficking, and Autophagy, and (4) Metabolism and Cellular Stress. A schematic figure was created to visually represent these categories, using illustrative icons and anatomical overlays to indicate areas typically affected in osteoarthritis. Gene names were listed under each category to reflect their putative function. BioRender was used to assemble and style the visual elements.

## Supplementary Note 3: STROBE-MR checklist of recommended items to address in reports of Mendelian randomization studies^1^ ^2^

Note: Page number will be added at the proof-reading stage.

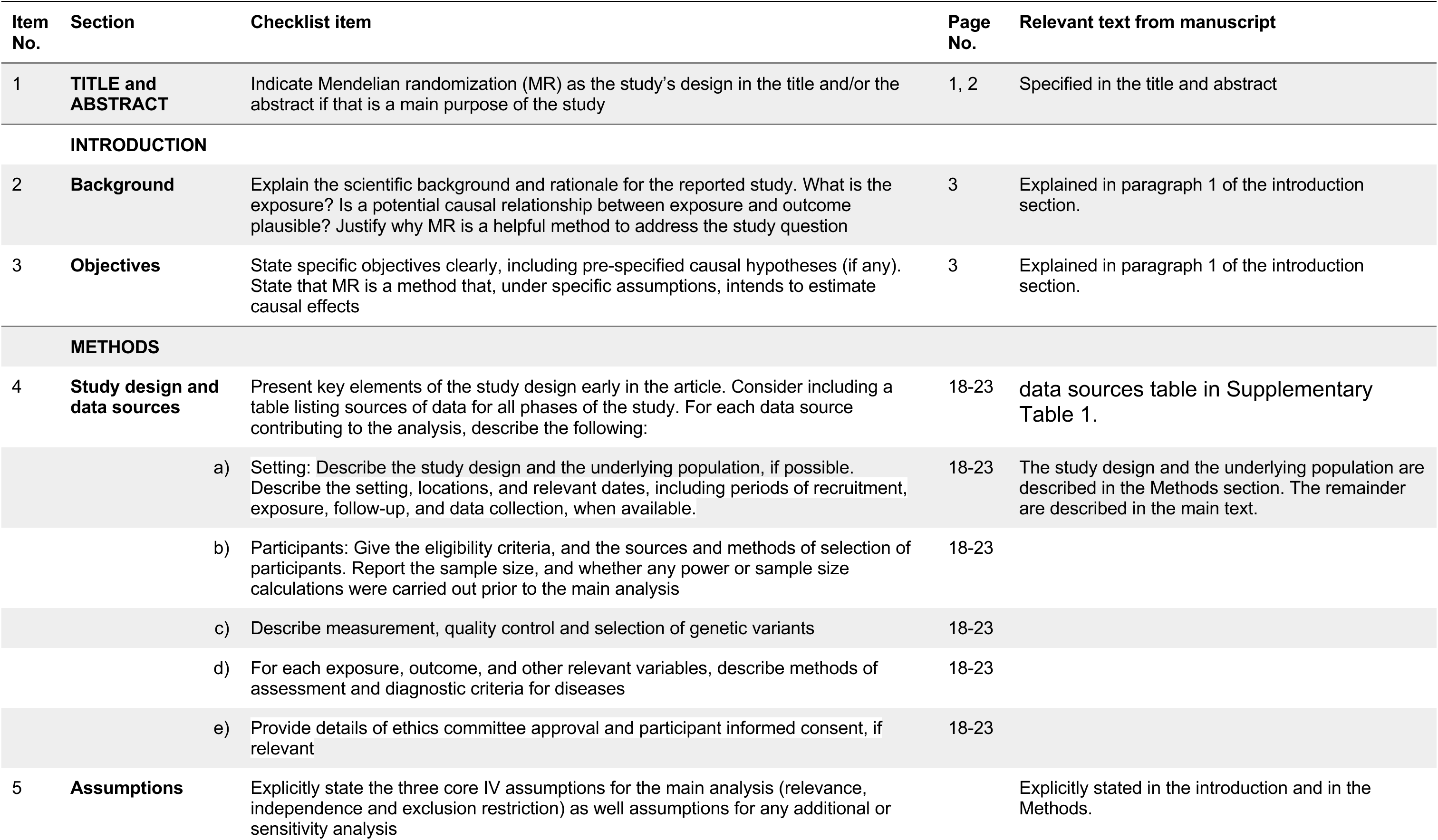
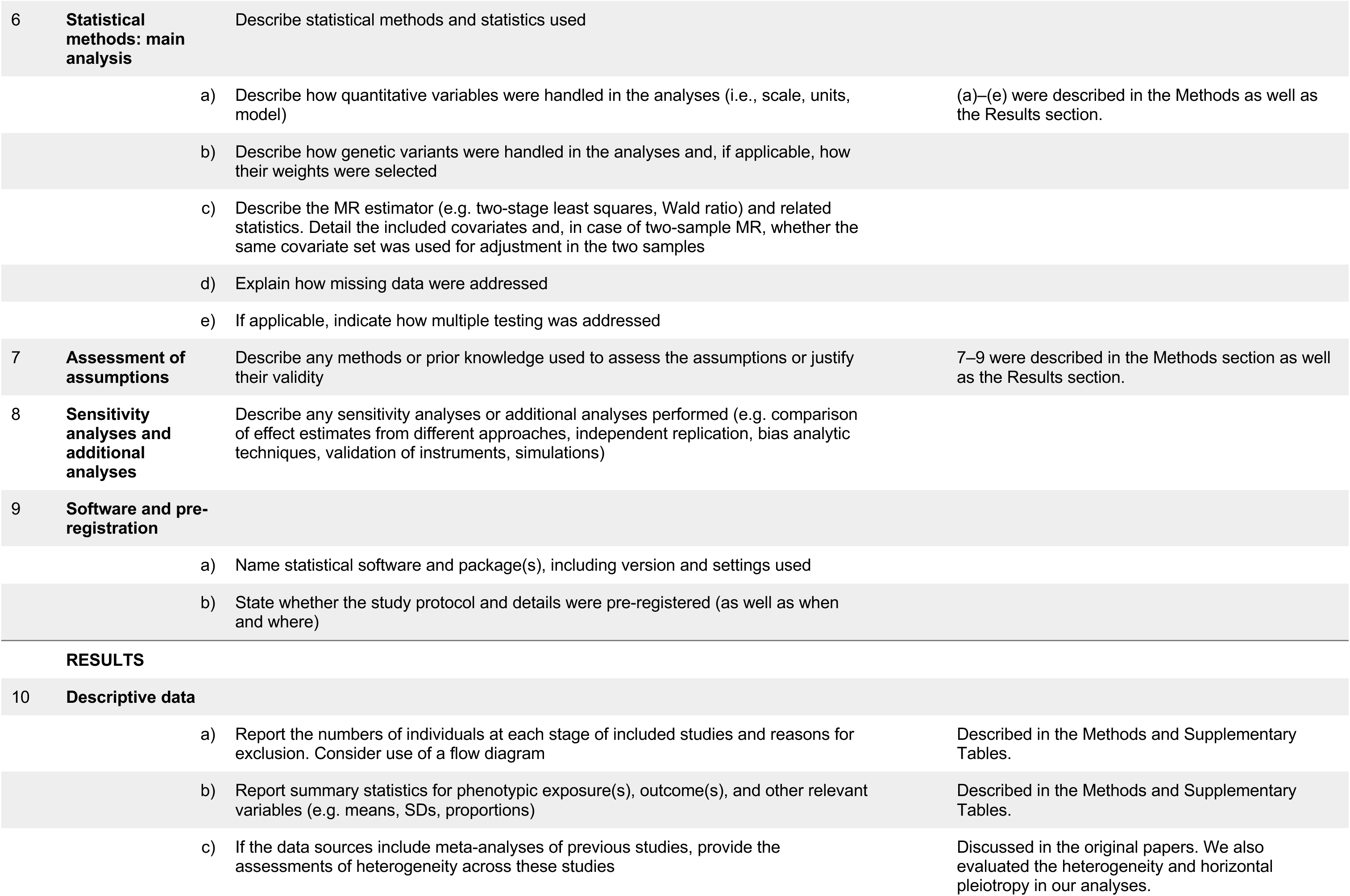
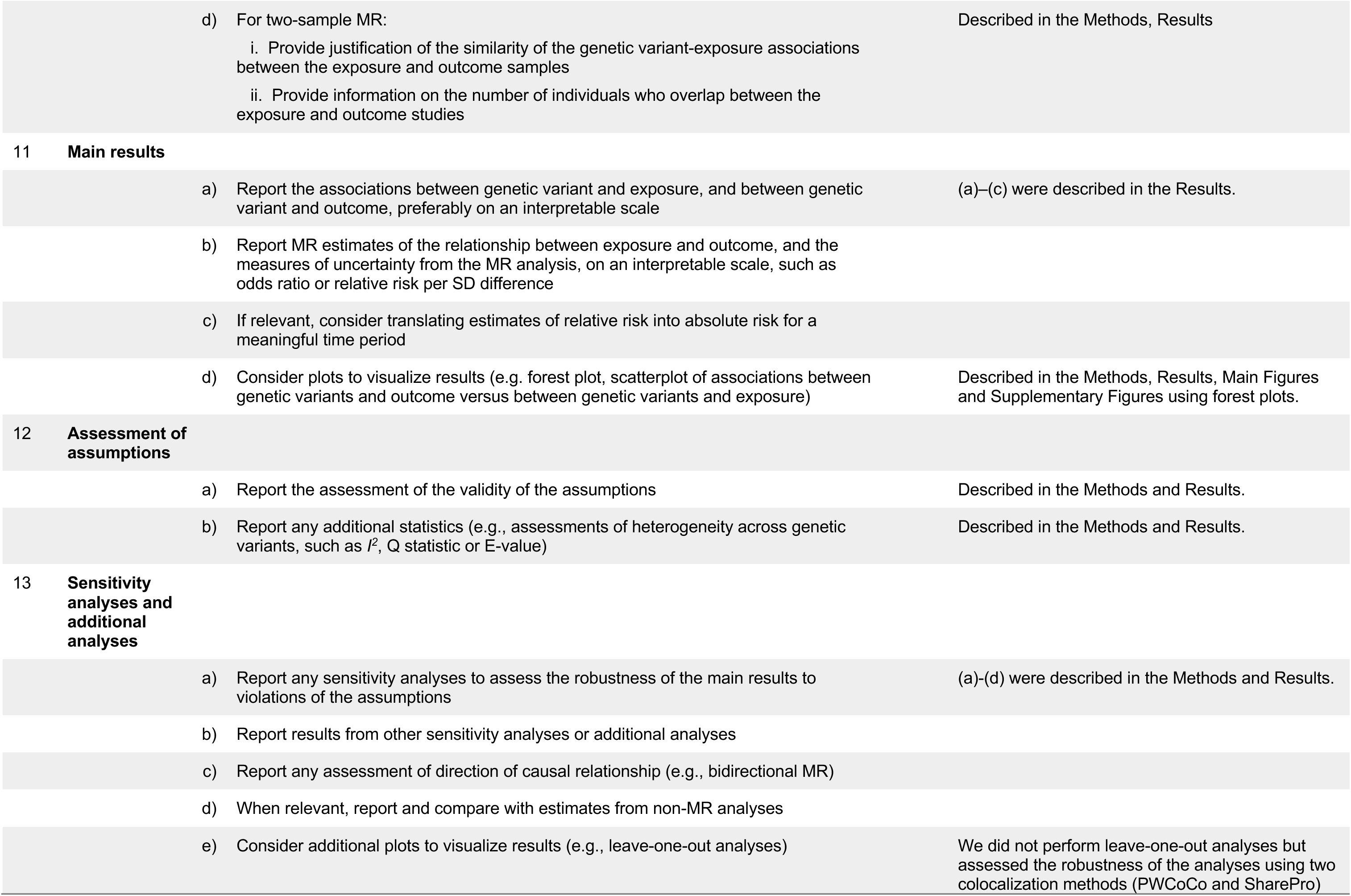
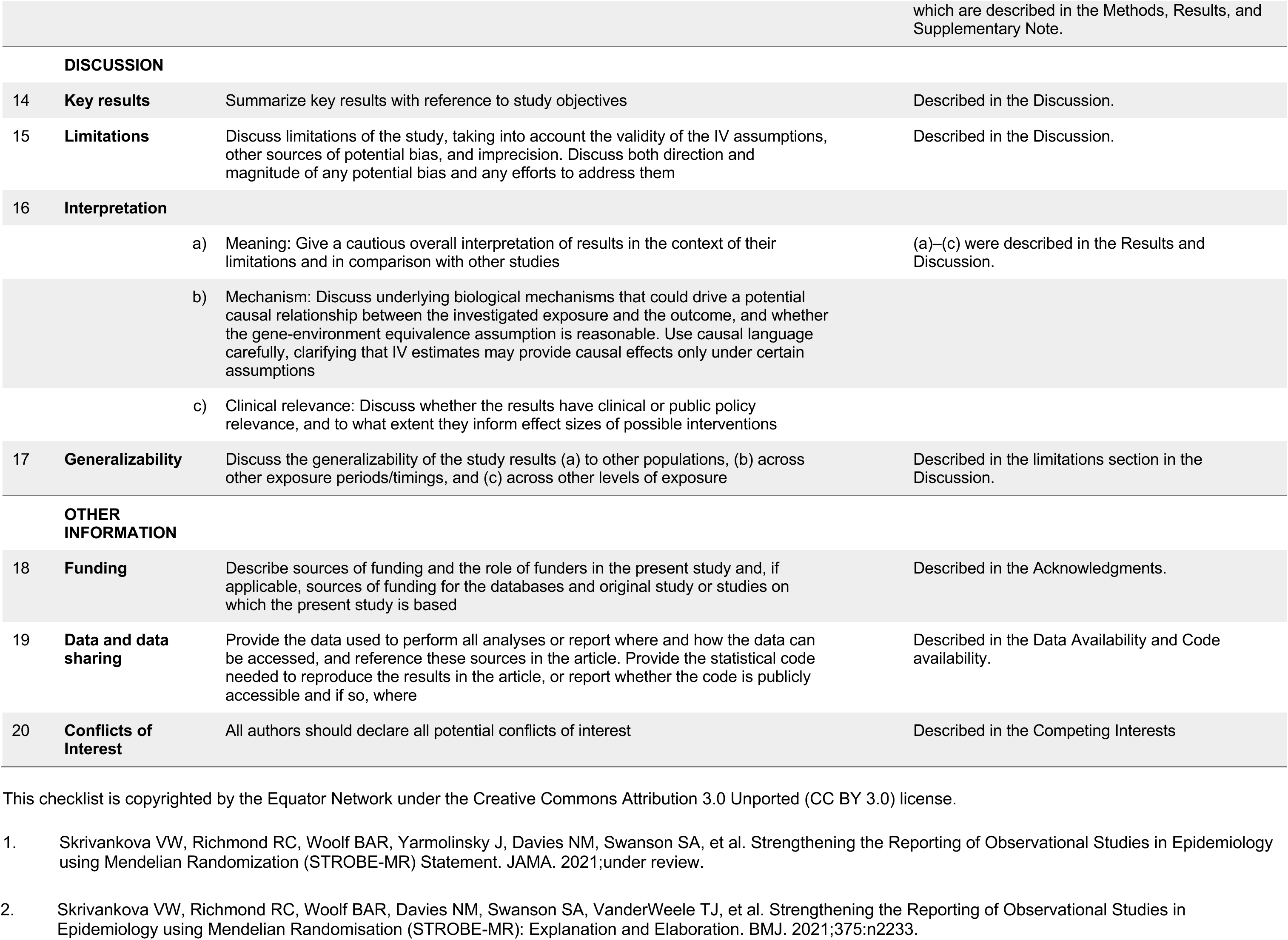

| Item No. | Section | Checklist item | Page No. | Relevant text from manuscript |
| --- | --- | --- | --- | --- |
| 1 | **TITLE and ABSTRACT** | Indicate Mendelian randomization (MR) as the study's design in the title and/or the abstract if that is a main purpose of the study | 1, 2 | Specified in the title and abstract |
| **INTRODUCTION** |  |  |  |  |
| 2 | **Background** | Explain the scientific background and rationale for the reported study. What is the exposure? Is a potential causal relationship between exposure and outcome plausible? Justify why MR is a helpful method to address the study question | 3 | Explained in paragraph 1 of the introduction section. |
| 3 | **Objectives** | State specific objectives clearly, including pre-specified causal hypotheses (if any). State that MR is a method that, under specific assumptions, intends to estimate causal effects | 3 | Explained in paragraph 1 of the introduction section. |
| **METHODS** |  |  |  |  |
| 4 | **Study design and data sources** | Present key elements of the study design early in the article. Consider including a table listing sources of data for all phases of the study. For each data source contributing to the analysis, describe the following: | 18-23 | data sources table in Supplementary Table 1. |
|  | a) | Setting: Describe the study design and the underlying population, if possible. Describe the setting, locations, and relevant dates, including periods of recruitment, exposure, follow-up, and data collection, when available. | 18-23 | The study design and the underlying population are described in the Methods section. The remainder are described in the main text. |
|  | b) | Participants: Give the eligibility criteria, and the sources and methods of selection of participants. Report the sample size, and whether any power or sample size calculations were carried out prior to the main analysis | 18-23 |  |
|  | c) | Describe measurement, quality control and selection of genetic variants | 18-23 |  |
|  | d) | For each exposure, outcome, and other relevant variables, describe methods of assessment and diagnostic criteria for diseases | 18-23 |  |
|  | e) | Provide details of ethics committee approval and participant informed consent, if relevant | 18-23 |  |
| 5 | **Assumptions** | Explicitly state the three core IV assumptions for the main analysis (relevance, independence and exclusion restriction) as well assumptions for any additional or sensitivity analysis |  | Explicitly stated in the introduction and in the Methods. |

|  |  |  |  |
| --- | --- | --- | --- |
| 6 | **Statistical methods: main analysis** | Describe statistical methods and statistics used |  |
|  | a) | Describe how quantitative variables were handled in the analyses (i.e., scale, units, model) | (a)–(e) were described in the Methods as well as the Results section. |
|  | b) | Describe how genetic variants were handled in the analyses and, if applicable, how their weights were selected |  |
|  | c) | Describe the MR estimator (e.g. two-stage least squares, Wald ratio) and related statistics. Detail the included covariates and, in case of two-sample MR, whether the same covariate set was used for adjustment in the two samples |  |
|  | d) | Explain how missing data were addressed |  |
|  | e) | If applicable, indicate how multiple testing was addressed |  |
| 7 | **Assessment of assumptions** | Describe any methods or prior knowledge used to assess the assumptions or justify their validity | 7–9 were described in the Methods section as well as the Results section. |
| 8 | **Sensitivity analyses and additional analyses** | Describe any sensitivity analyses or additional analyses performed (e.g. comparison of effect estimates from different approaches, independent replication, bias analytic techniques, validation of instruments, simulations) |  |
| 9 | **Software and pre-registration** |  |  |
|  | a) | Name statistical software and package(s), including version and settings used |  |
|  | b) | State whether the study protocol and details were pre-registered (as well as when and where) |  |

|  |  |  |  |
| --- | --- | --- | --- |
| 10 | **Descriptive data** |  |  |
|  | a) | Report the numbers of individuals at each stage of included studies and reasons for exclusion. Consider use of a flow diagram | Described in the Methods and Supplementary Tables. |
|  | b) | Report summary statistics for phenotypic exposure(s), outcome(s), and other relevant variables (e.g. means, SDs, proportions) | Described in the Methods and Supplementary Tables. |
|  | c) | If the data sources include meta-analyses of previous studies, provide the assessments of heterogeneity across these studies | Discussed in the original papers. We also evaluated the heterogeneity and horizontal pleiotropy in our analyses. |

|  |  |  |
| --- | --- | --- |
|  | d) For two-sample MR:<br>i. Provide justification of the similarity of the genetic variant-exposure associations between the exposure and outcome samples<br>ii. Provide information on the number of individuals who overlap between the exposure and outcome studies | Described in the Methods, Results |
| **11** | **Main results** |  |
|  | a) Report the associations between genetic variant and exposure, and between genetic variant and outcome, preferably on an interpretable scale | (a)–(c) were described in the Results. |
|  | b) Report MR estimates of the relationship between exposure and outcome, and the measures of uncertainty from the MR analysis, on an interpretable scale, such as odds ratio or relative risk per SD difference |  |
|  | c) If relevant, consider translating estimates of relative risk into absolute risk for a meaningful time period |  |
|  | d) Consider plots to visualize results (e.g. forest plot, scatterplot of associations between genetic variants and outcome versus between genetic variants and exposure) | Described in the Methods, Results, Main Figures and Supplementary Figures using forest plots. |
| **12** | **Assessment of assumptions** |  |
|  | a) Report the assessment of the validity of the assumptions | Described in the Methods and Results. |
| | b) Report any additional statistics (e.g., assessments of heterogeneity across genetic variants, such as $I^2$, Q statistic or E-value) | Described in the Methods and Results. |
| **13** | **Sensitivity analyses and additional analyses** |  |
|  | a) Report any sensitivity analyses to assess the robustness of the main results to violations of the assumptions | (a)–(d) were described in the Methods and Results. |
|  | b) Report results from other sensitivity analyses or additional analyses |  |
|  | c) Report any assessment of direction of causal relationship (e.g., bidirectional MR) |  |
|  | d) When relevant, report and compare with estimates from non-MR analyses |  |
|  | e) Consider additional plots to visualize results (e.g., leave-one-out analyses) | We did not perform leave-one-out analyses but assessed the robustness of the analyses using two colocalization methods (PWCoCo and SharePro) |

|  |  |  |  |
| --- | --- | --- | --- |
|  |  |  | which are described in the Methods, Results, and Supplementary Note. |
| **DISCUSSION** |  |  |  |
| 14 | **Key results** | Summarize key results with reference to study objectives | Described in the Discussion. |
| 15 | **Limitations** | Discuss limitations of the study, taking into account the validity of the IV assumptions, other sources of potential bias, and imprecision. Discuss both direction and magnitude of any potential bias and any efforts to address them | Described in the Discussion. |
| 16 | **Interpretation** | a) Meaning: Give a cautious overall interpretation of results in the context of their limitations and in comparison with other studies<br>b) Mechanism: Discuss underlying biological mechanisms that could drive a potential causal relationship between the investigated exposure and the outcome, and whether the gene-environment equivalence assumption is reasonable. Use causal language carefully, clarifying that IV estimates may provide causal effects only under certain assumptions<br>c) Clinical relevance: Discuss whether the results have clinical or public policy relevance, and to what extent they inform effect sizes of possible interventions | (a)–(c) were described in the Results and Discussion. |
| 17 | **Generalizability** | Discuss the generalizability of the study results (a) to other populations, (b) across other exposure periods/timings, and (c) across other levels of exposure | Described in the limitations section in the Discussion. |
| **OTHER INFORMATION** |  |  |  |
| 18 | **Funding** | Describe sources of funding and the role of funders in the present study and, if applicable, sources of funding for the databases and original study or studies on which the present study is based | Described in the Acknowledgments. |
| 19 | **Data and data sharing** | Provide the data used to perform all analyses or report where and how the data can be accessed, and reference these sources in the article. Provide the statistical code needed to reproduce the results in the article, or report whether the code is publicly accessible and if so, where | Described in the Data Availability and Code availability. |
| 20 | **Conflicts of Interest** | All authors should declare all potential conflicts of interest | Described in the Competing Interests |

This checklist is copyrighted by the Equator Network under the Creative Commons Attribution 3.0 Unported (CC BY 3.0) license.
1. Skrivankova VW, Richmond RC, Woolf BAR, Yarmolinsky J, Davies NM, Swanson SA, et al. Strengthening the Reporting of Observational Studies in Epidemiology using Mendelian Randomization (STROBE-MR) Statement. JAMA. 2021;under review.
2. Skrivankova VW, Richmond RC, Woolf BAR, Davies NM, Swanson SA, VanderWeele TJ, et al. Strengthening the Reporting of Observational Studies in Epidemiology using Mendelian Randomisation (STROBE-MR): Explanation and Elaboration. *BMJ*. 2021;375:n2233.

## Notes

### Competing Interest Statement

The authors have declared no competing interest.

### Author Declarations

The study used previously published GWAS summary statistics (OA from the Genetics of Osteoarthritis Consortium; BMI from GIANT/UK Biobank) and proteomics pQTL resources from established cohorts (ARIC, deCODE, Fenland, UKB-PPP); it did not involve new human subject recruitment, IRB review, or gated/request-only datasets.

